# Robust Longitudinal Dementia Prediction under Systemic Missingness via Hierarchical Fusion and Test-Time Adaptation

**DOI:** 10.64898/2026.07.02.26357089

**Authors:** Chen Zhang, Hetu Li, Fang Tian, Mansour L. Sina, Csaba Orban, Christopher Chen, Juan Helen Zhou, B.T. Thomas Yeo, the Alzheimer’s Disease Neuroimaging Initiative, Australian Imaging Biomarkers and Lifestyle Study of Ageing

## Abstract

Longitudinal dementia progression prediction is essential for clinical decision-making. However, models often degrade on external cohorts due to systemic missingness — where certain biomarkers available during training are completely absent at test time — compounded by distribution shifts and patient-specific variability. Here, we propose Progression-aware Feature Fusion with Test-Time Adaptation (ProFuse-TTA), a two-stage hierarchical Transformer for longitudinal dementia prediction. Stage 1 learns per-biomarker temporal representations from irregular observations without imputation. Stage 2 fuses them via cross-feature attention, with simulated modality dropout during training for robustness to systemic missingness. At inference, a lightweight test-time adaptation module performs per-individual calibration. We trained on ADNI and evaluated on three external cohorts comprising 2,316 participants and 13,205 timepoints, with controlled modality ablation experiments isolating the effect of systemic missingness. We compared against six baselines, four from a recent benchmark study and two new baselines including one built on a tabular foundation model. ProFuse-TTA achieved the best cross-dataset performance in 8 of 9 settings across clinical diagnosis, MMSE, and hippocampal volume prediction, and ranked first in 14 of 15 ablation scenarios. The model maintained superior performance across varying input lengths and prediction horizons up to 6 years. Pretrained ADNI models are available at XXX.

## 1 Introduction

Alzheimer’s disease (AD) dementia is a neurodegenerative disorder characterized by neuropathological changes beginning years before dementia onset (Villemagne et al., 2013; Jack et al., 2018; Hampel et al., 2021). Although no cure exists, disease-modifying therapies targeting early-stage pathology have shown promise in slowing cognitive decline (Cummings et al., 2019; Van Dyck et al., 2023), making the identification of at-risk individuals an important goal (Dubois et al., 2016; Scheltens et al., 2021; Aisen et al., 2022). Accurate forecast of dementia progression can enable earlier intervention and more efficient clinical trial recruitment (de Vugt & Verhey, 2013; Rasmussen & Langerman, 2019; Burns et al., 2021; Oxtoby et al., 2022).

Most AD prediction studies adopt setups that do not reflect the complexity of real-world clinical setting. Many studies assume a single observed timepoint (Hett et al., 2021; Hebling Vieira et al., 2022; C. Wang et al., 2022), restrict analyses to participants with complete multimodal data (Golovanevsky et al., 2022; C. Wang et al., 2022; Reas et al., 2023), and/or predict a single outcome within a fixed window, e.g., whether a patient with mild cognitive impairment (MCI) converts to dementia within 3 years (Basaia et al., 2019; El-Sappagh et al., 2021; Ocasio & Duong, 2021). In practice, patients have varying numbers of visits, not every modality is measured at every visit, and clinical decisions may require prediction at any future horizon. The TADPOLE challenge (Marinescu et al., 2019, 2021) introduced a prediction setup that better reflects these conditions: (1) variable number of input timepoints per participant, (2) missing data across modalities and visits, and (3) open-ended forecast horizon.

Beyond the prediction setup, models are often trained and evaluated within the same dataset (Karaman & Sabuncu, 2024; Moghaddami et al., 2025). Single-cohort evaluation can overestimate generalizability (Chekroud et al., 2024), since external cohorts often differ in population characteristics, acquisitions, and data processing (Jovicich et al., 2006; Hawco et al., 2018; Bigler et al., 2020; Kang et al., 2020). We previously evaluated five algorithms based on the TADPOLE setup, where models were trained on the Alzheimer’s Disease Neuroimaging Initiative (ADNI) dataset and evaluated on three external cohorts (Zhang et al., 2025). We found that best cross-cohort performance was enabled by the longitudinal-to-cross-sectional (L2C) transformation, which was used by the winner of the TADPOLE challenge (Marinescu et al., 2021).

The L2C transformation converts variable-length longitudinal histories into fixed-length feature vectors. Although participants initially have different numbers of input features due to varying visit counts and missing data, the L2C transformation ensures that each participant has the same number of features. This unlocks two benefits. First, the same-length L2C features can be fed to powerful algorithms (e.g., XGBoost) that do not natively handle variable-length inputs. Second, the L2C transformation captures temporal statistics (e.g., rate of change, historical maximum / minimum) that can be computed despite sporadic missing data (Section 2.6).

However, our previous study only used biomarkers common across cohorts (Zhang et al., 2025), which is unrealistic. Sporadic missingness arises from incomplete observations due to missed appointments, inability to complete assessments, or acquisition issues (Hardy et al., 2009; Ibrahim et al., 2012; Esteban et al., 2019; Chowdhry et al., 2021). L2C is robust to sporadic missingness, but is not robust to systemic missingness, where certain biomarkers available during training are completely absent from a target dataset. Systemic missingness is driven by study design, evolving biomarker availability, and resource constraints (Aisen et al., 2010; Jack et al., 2018; Hansson, 2021; Schöll et al., 2024). For example, a model trained on ADNI with MRI and cognitive features may need to be deployed in a clinic where only cognitive features are available. Re-training a separate model for each clinic is not scalable. Thus, systemic missingness remains an important but underexplored challenge in dementia progression prediction.

Several classical approaches handle sporadic missing data that could be adapted for systemic missingness. The simplest approach is to impute missing values using methods such as Multiple Imputation by Chained Equations (MICE; White et al., 2011; Aschwanden et al., 2020; El-Sappagh et al., 2021) before feeding them to a downstream predictor. A second approach integrates imputation and prediction in a “model-filling” framework, where a dynamical state model updates a latent disease state from observed data and uses it to fill missing values at the next timepoint. This was introduced by MinimalRNN (Nguyen et al., 2020), which ranked second on the TADPOLE leaderboard, and has been adopted by subsequent studies (Jung et al., 2021; W. Liang et al., 2021; Xu et al., 2022; Cheng et al., 2024). Finally, tree-based models like XGBoost (T. Chen & Guestrin, 2016) naturally learn split directions for missing values. In the current study, we evaluate L2C combined with XGBoost (L2C-XGB) or MICE (L2C-MICE), as well as MinimalRNN, as baselines.

The transformer architecture (Vaswani et al., 2017) is well-suited to missing values because self-attention can integrate available inputs while down-weighting missing ones at inference. Recent work has applied transformers directly to longitudinal AD progression (Karaman & Sabuncu, 2024; Moghaddami et al., 2025), with each visit serving as a token and all biomarkers stacked within the token vector. While these designs handle sporadic missingness via imputation or missingness masks, the visit-as-token formulation does not protect against systemic missingness. In multimodal settings, robustness to systemic missingness can be improved by training with incomplete subsets of inputs (L. Liu et al., 2023; Tang et al., 2024; Xue et al., 2024; Yu et al., 2024). However, multimodal transformers often allocate token dimension to modalities rather than timepoints, so they have been mostly applied to cross-sectional prediction. The Tabular Prior-Fitted Network (TabPFN), a transformer-based tabular foundation model pretrained on millions of synthetic datasets with injected missingness, outperformed strong tabular baselines across 57 datasets (Hollmann et al., 2025), but is also cross-sectional. A workaround is to combine multi-modal transformers or TabPFN with L2C. Here we include L2C-TabPFN as a baseline in the current study. However, L2C is a hand-engineered feature extraction step, so temporal representations are not learned with the prediction task.

To handle systemic missingness while learning temporal representations, we propose Progression-aware Feature Fusion with Test-Time Adaptation (ProFuse-TTA), a two-stage hierarchical transformer. In stage 1, feature-wise temporal encoders learn per-biomarker temporal representations from irregular longitudinal observations, conceptually similar to L2C but learned end-to-end. In stage 2, these representations are fused through cross-feature attention, with simulated modality dropout during training to provide robustness to systemic missingness. Since training-time robustness alone cannot eliminate residual cross-cohort differences at inference, a test-time adaptation (TTA) module performs lightweight per-individual calibration.

We train ProFuse-TTA and all baselines on ADNI and evaluate them on three external cohorts comprising 2316 participants with 13,205 time points from the United States, Australia, and Singapore.

## 2 Methods

### 2.1 Problem overview

The problem setup followed our previous study (Zhang et al., 2025), which built upon the TADPOLE challenge formulation (Marinescu et al., 2019, 2021). Given multimodal biomarkers and diagnostic history (Table 1) at one or more timepoints of an individual, we aimed to predict the cognitive state, hippocampus volume normalized by intracranial volume (ICV), and clinical diagnosis of the individual for every subsequent month beyond the last observed timepoint up to 120 months into the future. While the original TADPOLE challenge used the Alzheimer’s Disease Assessment Scale Cognitive Subdomain (ADAS-Cog13), we switched to predicting the Mini-Mental State Examination (MMSE) score, as it was more consistently available across all datasets in this study.

We used four longitudinal datasets: the Alzheimer’s Disease Neuroimaging Initiative (ADNI) dataset, the Australian Imaging Biomarkers and Lifestyle Study of Ageing (AIBL) dataset, the Memory, Ageing and Cognition Centre (MACC) Harmonization Cohort, and the Open Access Series of Imaging Studies (OASIS) dataset. These datasets contained overlapping subsets of modalities, including T1-weighted structural MRI, positron emission tomography (PET), cerebrospinal fluid (CSF) biomarkers, cognitive assessments, clinical diagnoses, and demographic information, with ADNI providing the most comprehensive set. The diagnostic categories corresponded to cognitively normal (CN), mild cognitive impairment (MCI), and dementia of various etiologies (DEM). For clarity, in Table 1, we distinguish between dynamic features that are measured longitudinally over time and static features defined at baseline. Data collection was approved by the Institutional Review Board (IRB) at each corresponding institution. The analysis in the current study is approved by the National University of Singapore IRB.

Compared to our prior study (Zhang et al., 2025), where both training and external test datasets shared a common subset of biomarkers, a key goal of the present study is to evaluate model robustness to “systemic missingness”. We define a “modality” as a broader data category such as structural MRI, PET, CSF, cognitive assessments, or clinical diagnosis, each of which may contain multiple “biomarkers” (e.g., hippocampal volume from MRI, FDG SUVR from PET, MMSE from cognitive assessments). We define “systemic missingness” as the complete absence of a biomarker in a test dataset that was present during training.

In this study, we incorporated additional PET, CSF, and cognitive biomarkers into the ADNI training dataset, while recognizing that such measures were inconsistently available in the external test cohorts (AIBL, MACC, OASIS). To create a challenging and clinically relevant evaluation, we standardized the test sets to reflect a common clinical reality: widespread access to cognitive assessments and structural MRI, but limited availability of more costly or invasive measures (Hornberger et al., 2017; Roth et al., 2023; Schöll et al., 2024). Accordingly, while retaining all cognitive scores, we excluded all PET and CSF biomarkers from the three external test sets. This produced nearly identical feature sets across test cohorts, with the only exception being the Montreal Cognitive Assessment (MoCA) score, available in MACC and OASIS but not in AIBL (Table 1). This design directly tests each model’s ability to generalize from a rich, multimodal training environment (ADNI) to a more resource-constrained but common clinical setting.

**Table 1.** Feature availability across training and testing datasets.

| Category | Modality / Feature |  | Training Dataset | Test Datasets |
| --- | --- | --- | --- | --- |
| **Static features** | Sex (male, female), Genetics (number of APOE-ε4 allele), Marital status (married, not married), Education level (number of years of education) | | ✓ | ✓ |
| **Dynamic features** | MRI features | Hippocampus, Entorhinal, Fusiform, Middle Temporal, Ventricle, Whole Brain, ICV | ✓ | ✓ |
|  | Demographics | Age | ✓ | ✓ |
|  | Cognitive features | MMSE (0-30) | ✓ | ✓ |
|  |  | CDRSB (0-18) | ✓ | ✓ |
|  |  | MoCA (0-30) | ✓ | ✓<sup>#</sup> |
|  |  | ADAS-Cog11 (0-70) | ✓ |  |
|  |  | ADAS-Cog13 (0-85) | ✓ |  |
|  |  | RAVLT_immediate (0-75) | ✓ |  |
|  |  | RAVLT_learning (0-15) | ✓ |  |
|  |  | RAVLT_forgetting (0-15) | ✓ |  |
|  |  | RAVLT_perc_forgetting (0-100) | ✓ |  |
|  |  | FAQ (0-30) | ✓ |  |
|  | Diagnostic features | Clinical diagnosis (CN, MCI, DEM) | ✓ | ✓ |
|  | PET features | AV45 (Florbetapir) SUVR, FDG (Fluorodeoxyglucose) SUVR | ✓ |  |
| | CSF features | ABETA (Aβ42) concentration, TAU (total-Tau) concentration, PTAU (phosphorylated-Tau) concentration | ✓ | |

A checkmark (✓) indicates the feature was included in the corresponding dataset for this study. Abbreviations: ICV: intracranial volume. MMSE: Mini Mental State Examination. CDRSB: Clinical Dementia Rating – Sum of Boxes. MoCA: Montreal Cognitive Assessment. ADAS-Cog11: Alzheimer’s Disease Assessment Scale Cognitive Subdomain (11-item). ADAS-Cog13: Alzheimer’s Disease Assessment Scale Cognitive Subdomain (13-item). RAVLT: Rey Auditory Verbal Learning Test (immediate: the sum of scores from 5 first trials (Trials 1 to 5); learning: the score of Trial 5 minus the score of Trial 1; forgetting: the score of Trial 5 minus score of the delayed recall; perc_forgetting: RAVLT Forgetting divided by the score of Trial 5). FAQ: Functional Activities Questionnaire. CN: cognitively normal. MCI: mild cognitive impairment. DEM: Dementia with various etiologies. SUVR: Standardized Uptake Value Ratio. ^#^MoCA was available and used for MACC and OASIS, but not for AIBL. The L2C variants and ProFuse-TTA make use of all 28 features in Table 1. MinimalRNN does not support covariates and therefore omits the static features as well as age (see Section 2.4 for details). AD-Map handles neither covariates nor categorical inputs, so it additionally omits clinical diagnosis (see Section 2.5 for details).

### 2.2 Datasets, preprocessing and participant selection

#### 2.2.1 Datasets

Data used in the preparation of this article were obtained from the Alzheimer’s Disease Neuroimaging Initiative (ADNI) database (adni.loni.usc.edu). The ADNI was launched in 2003 as a public-private partnership, led by Principal Investigator Michael W. Weiner, MD. The primary goal of ADNI has been to test whether serial magnetic resonance imaging (MRI), positron emission tomography (PET), other biological markers, and clinical and neuropsychological assessment can be combined to measure the progression of mild cognitive impairment (MCI) and early Alzheimer’s disease (AD).

ADNI is a large-scale, multisite research program based in the United States (Jack et al., 2008; Petersen et al., 2010). Three phases have been completed to date: ADNI1 (2004-2009), ADNI-GO/2 (2010-2016), and ADNI3 (2017-2023). Each phase recruited new participants while also retaining individuals from earlier phases. MRI hardware also evolved over the course of the study: ADNI1 relied predominantly on 1.5T scanners, whereas later phases moved to 3T systems (see Table S1 for details). T1-weighted MRI scans, together with the ADNIMERGE spreadsheet (the ADNI team, 2023) — which compiles demographics, clinical diagnoses, cognitive assessments, PET, and CSF measurements — were retrieved from the USC Laboratory of Neuro Imaging’s Image and Data Archive (IDA).

The AIBL study (Fowler et al., 2021) is a large prospective Australian cohort whose study design and biomarker protocols closely mirror those of ADNI. T1-weighted scans were collected on a combination of 1.5T and 3T platforms (Avanto, Tim Trio, and Verio; see Table S2 for details), and both the imaging and accompanying phenotypic data were retrieved from the USC Laboratory of Neuro Imaging’s Image and Data Archive (IDA).

The MACC Harmonization cohort (Lim et al., 2025) represents a memory clinic population in Singapore. In this dataset, T1-weighted MRI scans were collected exclusively using 3T scanners (Tim Trio and Prisma; see Table S3 for details). The cohort includes individuals diagnosed with vascular dementia and/or Alzheimer’s disease dementia; for the purposes of this study, these were combined into a single category termed Vascular/Alzheimer’s disease dementia (DEM). The mixed pathology enables assessment of model generalizability beyond Alzheimer’s disease dementia alone.

The OASIS dataset (LaMontagne et al., 2019) is an open-access multimodal resource intended to support research on healthy ageing and cognitive decline. It comprises four releases (OASIS-1, OASIS-2, OASIS-3, and OASIS-4). We used OASIS-3, the principal large-scale release, which subsumes participants from the smaller OASIS-1 (cross-sectional) and OASIS-2 (longitudinal) studies and is independent of the clinical OASIS-4 cohort. Following the same convention as MACC, we grouped both AD and non-AD dementia cases into a single DEM category. OASIS MRI was acquired across a mix of 1.5T and 3T scanners (see Table S4 for details), and both imaging and phenotypic data were obtained from XNAT Central (Herrick et al., 2016).

#### 2.2.2 Preprocessing

T1-weighted MRI scans were first deobliqued and reoriented to RPI orientation. Volumetric measurements of regions of interest (ROIs) were then obtained by running each scan through the FreeSurfer 6.0 recon-all pipeline (Fischl et al., 2002; Desikan et al., 2006). From this output we retained six AD-relevant ROI volumes — hippocampus, entorhinal cortex, fusiform gyrus, middle temporal gyrus, ventricles, and whole brain (see Table S5 for details) — together with intracranial volume (ICV). The six ROI volumes were then divided by ICV to correct for head size, and ICV itself was retained as an additional feature (Table 1).

The resulting brain ROI features were merged with the downloaded phenotypic data — demographics, clinical diagnoses, and cognitive, PET, and CSF measurements. Because MRI visits and non-MRI assessments did not always occur on the same day, we adopted the ADNIMERGE convention of treating an MRI visit and a non-MRI assessment as a single timepoint whenever the two visits were separated by no more than six months. The merged timepoint inherited the date from the non-MRI visit.

We systematically removed empty or duplicate entries, along with those displaying outliers or errors. Certain datasets used inconsistent coding for missing values, such as NaN or special integers (e.g., −1, −4, 999). In the ADNI dataset, CSF TAU measurements occasionally (∼0.38%) appeared as out-of-range strings (e.g., “<80”, “>1300”), which were likewise treated as missing values during data cleaning. To ensure consistency, we replaced all such special entries with NaN. Additionally, rare negative values (∼2.5%) in derived RAVLT metrics in ADNI (e.g., learning, forgetting, percent forgetting) were set to zero to create a consistent non-negative data distribution for modeling. This process resulted in a clean longitudinal data table where each row represents a single participant visit, containing MRI features and/or other clinical data.

#### 2.2.3 Participant selection and characteristics

Because our goal was to forecast dementia trajectories, we restricted each of the four cohorts to participants who had at least two visits with non-empty dynamic features (Table 1). The dynamic features were not required to be the same across visits. For example, a participant whose first visit contained only MRI measurements and whose second visit contained only cognitive scores still met this criterion.

After applying this filter, the final ADNI cohort contained 2,114 participants spanning 15,801 timepoints (of which 9,671 included MRI features). The corresponding figures were 403 participants and 1,222 timepoints (942 with MRI) for AIBL, 653 participants and 3,070 timepoints (1,456 with MRI) for MACC, and 1,260 participants and 8,913 timepoints (2,519 with MRI) for OASIS.

The four cohorts differ markedly in demographics, severity, and visit density (Table 2; see Figures S1 and S2 for full distributions). Relative to ADNI, AIBL participants were on average younger, had higher MMSE scores, and skewed more heavily toward CN. MACC participants had lower MMSE scores, as well as a higher proportion of female participants and individuals with a DEM diagnosis. OASIS participants, like those in AIBL, were younger and more cognitively intact than the ADNI sample on average, with a higher proportion of female and CN participants.

**Table 2.** Participant characteristics in the four datasets. Each external cohort (AIBL, MACC, OASIS) was compared against ADNI on every row of the table: numeric variables such as age and MMSE were tested with a permutation test, and categorical variables such as sex and diagnosis with a chi-square test. p-values surviving false discovery rate correction (FDR q < 0.05) are shown in bold. Baseline diagnosis percentages do not sum to 100% in OASIS because a subset of OASIS participants lack a recorded clinical diagnosis at baseline.

|  | ADNI<br>(N = 2114) | AIBL<br>(N = 403) |  | MACC<br>(N = 653) |  | OASIS<br>(N = 1260) |  |
| --- | --- | --- | --- | --- | --- | --- | --- |
| Measures | mean ± std | mean ± std | p | mean ± std | p | mean ± std | p |
| Baseline age (y) | 73.3 ± 7.2 | 72.4 ± 6.7 | **3.0e-2** | 72.7 ± 8.0 | 1.1e-1 | 69.0 ± 9.0 | **1.0e-4** |
| Baseline MMSE | 27.4 ± 2.6 | 28.0 ± 2.8 | **1.0e-4** | 21.6 ± 6.1 | **1.0e-4** | 28.3 ± 2.5 | **1.0e-4** |
| Sex (M/F) | 1135 / 979<br>(54% / 46%) | 194 / 209<br>(48% / 52%) | 4.6e-2 | 289 / 364<br>(44% / 56%) | **3.0e-5** | 560 / 700<br>(44% / 56%) | **2.5e-7** |
| Baseline diagnosis<br>(CN/MCI/DEM) | 748 / 995 / 371<br>(35%/47%/18%) | 320 / 48 / 35<br>(79%/12%/9%) | **8.5e-60** | 132 / 272 / 249<br>(20%/42%/38%) | **9.7e-30** | 741 / 27 / 187<br>(59%/2%/15%) | **1.9e-137** |
| Baseline DEM<br>(AD/Other dementias) | - | - | - | 195 / 54<br>(78% / 22%) | - | 53 / 134<br>(28% / 72%) | - |
| CN demographics |  |  |  |  |  |  |  |
| Baseline Age (y) | 73.0 ± 6.2 | 72.1 ± 6.4 | **2.3e-2** | 68.5 ± 7.6 | **1.0e-4** | 68.3 ± 8.4 | **1.0e-4** |
| Sex (M/F) | 333 / 415<br>(45% / 55%) | 150 / 170<br>(47% / 53%) | 5.2e-1 | 59 / 73<br>(45% / 55%) | 9.5e-1 | 319 / 422<br>(43% / 57%) | 6.0e-1 |
| MCI demographics |  |  |  |  |  |  |  |
| Baseline age (y) | 73.0 ± 7.5 | 75.2 ± 6.5 | 4.2e-2 | 72.9 ± 7.6 | 9.2e-1 | 71.8 ± 6.2 | 4.3e-1 |
| Sex (M/F) | 592 / 403<br>(59% / 41%) | 28 / 20<br>(58% / 42%) | 9.9e-1 | 130 / 142<br>(48% / 52%) | **7.1e-4** | 15 / 12<br>(56% / 44%) | 8.3e-1 |
| DEM demographics |  |  |  |  |  |  |  |
| Baseline age (y) | 74.6 ± 7.8 | 71.8 ± 8.8 | 5.2e-2 | 74.8 ± 7.7 | 7.6e-1 | 74.2 ± 7.3 | 6.1e-1 |
| Sex (M/F) | 210 / 161<br>(57% / 43%) | 16 / 19<br>(46% / 54%) | 2.9e-1 | 100 / 149<br>(40% / 60%) | **8.4e-5** | 96 / 91<br>(51% / 49%) | 2.8e-1 |
| Number of visits | 7.5 ± 4.5 | 3.0 ± 1.3 | **1.0e-4** | 4.7 ± 1.4 | **1.0e-4** | 7.1 ± 4.7 | **1.4e-2** |
| Follow-up duration (y) | 4.5 ± 3.4 | 3.3 ± 2.0 | **1.0e-4** | 3.9 ± 1.5 | **1.0e-4** | 7.8 ± 5.6 | **1.0e-4** |
| Percentage of timepoints with missing data for recurring features |  |  |  |  |  |  |  |
| CDRSB | 28.6% | 0.7% | **2.6e-100** | 1.5% | **2.0e-225** | 4.9% | **0.0** |
| MMSE | 30.1% | 0.4% | **7.0e-110** | 1.5% | **1.0e-242** | 16.4% | **1.7e-125** |
| MRI features | 38.8% | 22.9% | **3.5e-28** | 52.6% | **1.2e-45** | 71.7% | **0.0** |
| Diagnosis | 30.2% | 0.4% | **2.9e-110** | 1.2% | **7.3e-249** | 19.1% | **6.5e-82** |

Visit density also varied substantially. In AIBL and MACC most participants have fewer than seven visits within roughly a seven-year window, whereas a subset of ADNI and OASIS participants accumulate 20–30 visits over more than 15 years of follow-up. The fraction of timepoints with missing data likewise differed substantially across the four cohorts (Table 2).

### 2.3 Training, validation and test procedure

We benchmarked the proposed ProFuse-TTA model against six baseline methods: MinimalRNN, AD-Map, and four L2C-based variants (L2C-XGB, L2C-FNN, L2C-MICE, and L2C-PFN). Models were trained on ADNI and evaluated under two regimes — within-dataset via 20-fold cross-validation on ADNI itself, and cross-dataset on the three external cohorts (AIBL, MACC, and OASIS). For the within-ADNI evaluation, ADNI participants were assigned to 20 non-overlapping partitions. Since partitioning was performed at the participant level, all visits from a given participant remained in the same partition; this prevented any leakage of an individual’s data across the train/validation/test boundary.

To train a given model, 18 partitions were used for training, while 1 partition was used as a validation set for hyperparameter tuning. The remaining partition was used as test set to evaluate the within-ADNI performance of the trained model. This procedure was repeated 20 times with a different partition being the test set (e.g., partition 3) and the neighboring partition being the validation set (e.g., partition 4). Therefore, we ended up with 20 sets of trained models together with 20 sets of within-ADNI evaluation results. The 20 sets of trained models were applied without modification to AIBL, MACC, and OASIS for cross-cohort evaluation (Figure 1).

**Figure 1.**
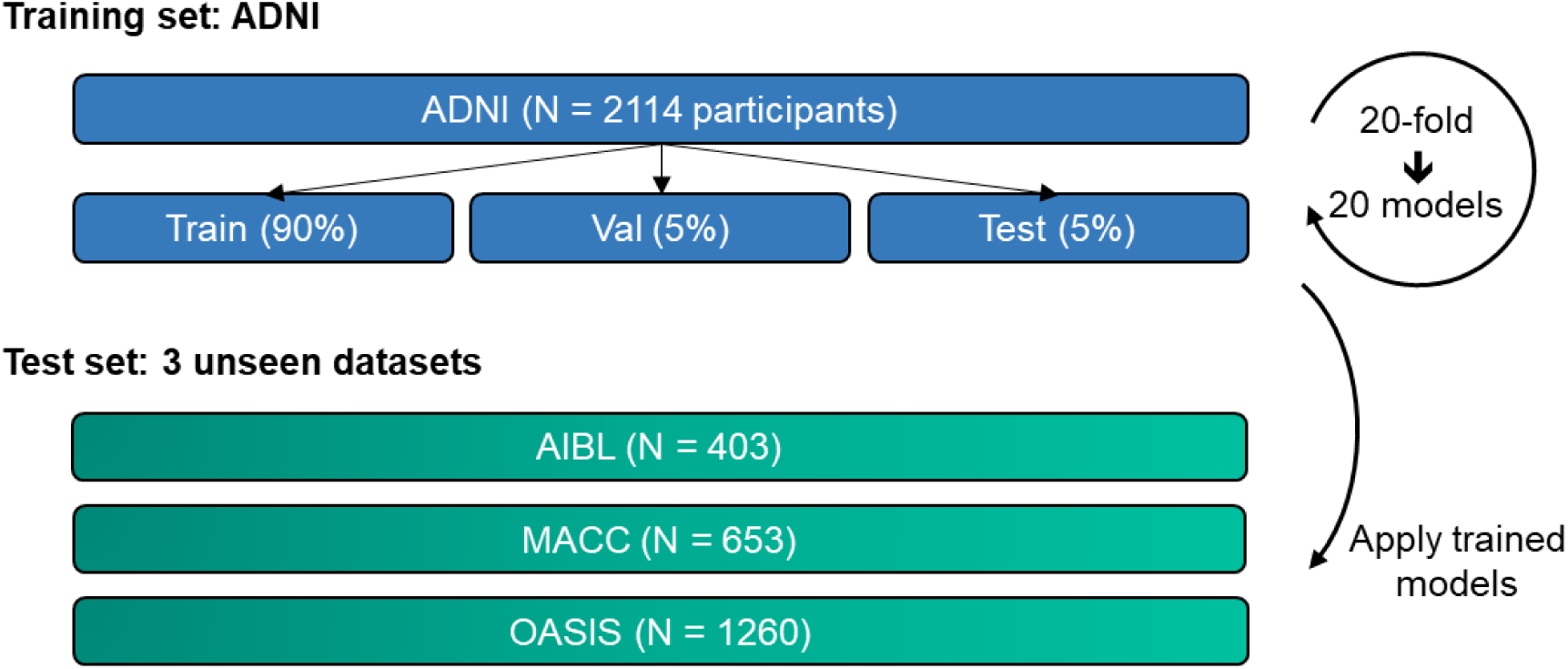
Training and testing procedure. All models were trained on the ADNI dataset and subsequently applied to three unseen test datasets to assess generalizability. Within ADNI, participants were split 18:1:1 across training, validation, and test partitions, and this assignment was rotated 20 times so that every participant appeared in the test set exactly once across folds. The 20 trained models obtained from these folds were then applied without modification to every participant in the three external cohorts for cross-cohort evaluation.

Following TADPOLE convention, for each participant in the ADNI validation and test sets, and for every participant in the three external cohorts, the first half of their visits served as observed input and the second half served as the prediction target. For example, for a participant with 10 visits, the first 5 visits (observed) were used to predict the remaining 5 visits (unobserved). During training, by contrast, the full longitudinal history of each training participant was used so as to maximize data usage.

Hyperparameters for each model were tuned on the validation partition with the Optuna library (Akiba et al., 2019), with a fresh hyperparameter search performed independently for every training/validation/test split. The search space for each algorithm is detailed in the corresponding method section.

### 2.4 MinimalRNN

MinimalRNN is a recurrent neural network (RNN), substantially leaner than LSTM (Long short-term memory). In our previous study tackling the same TADPOLE problem (Nguyen et al., 2020), MinimalRNN outperformed the more complex LSTM, and a simpler linear state space model. As such, the MinimalRNN’s model complexity was well-matched to the data, yielding the best prediction performance among the RNN models we tested.

In RNNs, a participant’s longitudinal data is processed sequentially. At each time step, a repeating computational unit updates an internal “disease state”, which is then used to predict the features of the next timepoint. To handle missing data during inference, MinimalRNN employs a “model-filling” strategy capable of addressing even systemic missingness. At the first timepoint, missing features are imputed with a default value of zero, corresponding to the mean of the z-scored training distribution. Subsequently, the model operates autoregressively: at each timepoint, it uses its hidden state and available input features to predict the next timepoint, using these predictions to fill any corresponding missing values before proceeding. This unified “model-filling” strategy enables MinimalRNN to function effectively, making it a strong baseline for this study.

For our experiments, we utilized the codebase from our previous study (Zhang et al., 2025)^1^, which was itself adapted from the original MinimalRNN implementation (Nguyen et al., 2020). Hyperparameter optimization was performed using Optuna (Akiba et al., 2019).

Table 3 summarizes the hyperparameters tuned by Optuna along with their search ranges. Following the original MinimalRNN design (Nguyen et al., 2020), the model was given only the dynamic features in Table 1 — MRI, cognitive, PET, CSF, and clinical-diagnosis variables — while the static baseline variables (sex, education, marital status, APOE-ε4 count) and age were withheld. Our previous experiments (not shown) found that these additional information did not improve prediction performance.

**Table 3.** Hyperparameter search ranges for MinimalRNN. Hyperparameters were tuned independently on every validation fold via Optuna.

| Hyper-parameter | Range |
| --- | --- |
| Input dropout rate | 0-0.5 |
| Recurrent dropout rate | 0-0.5 |
| L2 weight regularization | $10^{-7}$-$10^{-4}$ |
| Learning rate | $10^{-5}$-$10^{-2}$ |
| Number of hidden layers | 1-3 |
| Size of hidden state | 128, 256, 512 |

### 2.5 AD Course Map (AD-Map)

AD-Map is a Bayesian non-linear mixed-effects framework that jointly models trajectories of cognitive decline and regional brain atrophy. In our previous study (Zhang et al., 2025), AD-Map exhibited strong cross-dataset generalizability for continuous variables, including MMSE and ventricle volume, establishing it as a robust baseline for continuous outcomes. AD-Map models the progression of each biomarker using a population-level logistic trajectory, with different biomarkers having distinct progression rates and ages at the inflection point.

At the individual level, the model estimates individual-specific time-warp parameters (i.e., random effects) that map chronological age into a latent “disease age” shared across all biomarkers, along with individual-specific biomarker offsets. This latent axis reflects an individual’s position along the population disease trajectory and captures personal shifts in onset, progression rate, and the temporal ordering of biomarker changes. By projecting this disease age onto the population-level trajectories, AD-Map generates personalized biomarker curves, enabling prediction of expected biomarker values at any given age.

A key advantage of AD-Map is its native handling of both sporadic missing data and systemic missing biomarkers. During inference, individual parameters are estimated by maximizing the conditional likelihood over available observations, with missing values naturally excluded from the likelihood optimization. When a biomarker is completely absent from the test dataset, it is omitted from the estimation procedure. The model infers a shared latent disease state together with cross-biomarker offsets from the observed biomarkers and uses these to generate predictions for missing ones. This makes AD-Map a robust and interpretable baseline under systemic missingness.

We utilized the AD-Map implementation from our previous study (Zhang et al., 2025), which is based on the Leaspy software^2^. As with the other models, hyperparameters (Table 4) were optimized using Optuna (Akiba et al., 2019). ICV itself was excluded from the feature set because AD-Map operates on time-varying inputs, whereas ICV is essentially stable across adulthood (Courchesne et al., 2000; Jenkins et al., 2000); the remaining MRI volumes were still divided by ICV before being passed to the model, matching the preprocessing used elsewhere in this study.

**Table 4.** Hyperparameter search ranges for AD-Map. Hyperparameters were tuned independently on every validation fold via Optuna.

| Hyper-parameter | Range |
| --- | --- |
| Source dimension | 1-10 |
| # iterations | (1-10) * 500 |

The sigmoidal parameterization that underlies AD-Map applies only to continuous biomarkers, so categorical variables — clinical diagnosis in particular — can neither be supplied as inputs nor read out as predictions (Maheux et al., 2023). Following our previous study (Zhang et al., 2025), we therefore left clinical diagnosis out of the AD-Map model itself, following the original paper, and recovered diagnostic predictions post-hoc by mapping the model’s predicted CDRSB scores onto CN/MCI/DEM probabilities (see below). This preserves AD-Map’s design as a continuous-biomarker model while still allowing diagnostic prediction to be compared against the other algorithms.

The CDRSB-to-diagnosis mapping followed established cutoffs (O’Bryant et al., 2008, 2010): a CDRSB of 0 was assigned entirely to CN, values from 0.5 to 4.5 corresponded to MCI/DEM territory, and values from 4.5 to 18.0 were assigned to DEM. For predicted CDRSB scores falling between cutoffs we linearly interpolated the three class probabilities. As examples: a CDRSB of 0.1 yields probabilities of (CN, MCI, DEM) = (0.80, 0.20, 0.00); a CDRSB of 1.0 yields (0, 0.875, 0.125); a CDRSB at or above 4.5 is fully assigned to DEM.

Static covariates (sex, education, marital status, APOE-ε4 count) are likewise not supported by the AD-Map package. In summary, the AD-Map model was fit on age together with the dynamic biomarkers from Table 1 — MRI features (excluding ICV), cognitive scores, PET, and CSF — without static covariates or clinical diagnosis.

### 2.6 Longitudinal-to-Cross-sectional (L2C) transformation

Standard machine learning models (e.g., gradient-boosted trees, feedforward neural networks) typically require fixed-length inputs, which poses a challenge when modeling variable-length visit histories in longitudinal dementia progression studies. The L2C (Longitudinal-to-Cross-sectional) transformation is a feature engineering technique (Nanopoulos et al., 2001; Deng et al., 2013; Barandas et al., 2020) that addresses this challenge by summarizing an individual’s longitudinal trajectory into a fixed-length feature vector, providing the flexibility to use a wide range of downstream prediction models while preserving key temporal information. L2C was originally introduced as part of the FROG algorithm in the TADPOLE challenge (Marinescu et al., 2019), and was shown in our previous study (Zhang et al., 2025) to be highly effective and adaptable for longitudinal dementia progression prediction.

More specifically, consider an individual with measurements at *m* irregular timepoints *t*_1_, *t*_2_, *t*_3_, *t*_4_,…, *t_m_*, from which we seek to predict a target variable (e.g., clinical diagnosis, MMSE, or hippocampal volume) at a future timepoint *t_f_*. We note that the number of timepoints might be different across individuals. L2C converts each numeric biomarker (i.e., cognitive scores, anatomical ROI volumes, PET and CSF measurements) into seven features (Table 5) and transforms clinical diagnosis into eight features (Table 6), resulting in a total of 22 x 7 + 8 = 162 features.

**Table 5.** L2C feature names and their corresponding meaning for numeric input biomarkers.

| L2C feature names | Meaning |
| --- | --- |
| mr_fname | most recent measurement for feature |
| time_since_mr_fname | time since the most recent measurement |
| mr_change_fname | most recent change rate |
| low_fname | lowest historical measurement |
| time_since_low_fname | time since lowest historical measurement |
| high_fname | highest historical measurement |
| time_since_high_fname | time since highest historical measurement |

**Table 6.** L2C feature names and their corresponding meaning for clinical diagnosis.

| L2C feature names | Meaning |
| --- | --- |
| mr_dx | most recent non-missing diagnosis |
| time_since_mr_dx | time since the most recent non-missing diagnosis |
| best_dx | best historical diagnosis |
| time_since_best_dx | time since best historical diagnosis |
| worst_dx | worst historical diagnosis |
| time_since_worst_dx | time since worst historical diagnosis |
| milder | 1 if a milder diagnosis occurred in history |
| time_since_milder | time since the milder diagnosis and 999 if no milder diagnosis |

These 162 L2C features are further augmented with age at the prediction timepoint *t_f_*, baseline sex, baseline education level, baseline marital status, APOE status and the time interval between the first (baseline) visit and *t_f_* (months since baseline). In total, this yields 162 + 6 = 168 features. Please refer to Table S6 for the complete set of L2C features.

Beyond enabling flexible model selection, the L2C transformation also improves robustness to missing data. By collapsing longitudinal trajectories into summary statistics, L2C features can be computed even when only partial biomarker histories are available, thereby substantially reducing effective input missingness. Specifically, all features derived from a biomarker can be computed with as few as two observed timepoints, while only a single feature (e.g., the most recent change rate) remains undefined when only one observation is available.

In addition, the L2C formulation enables natural data augmentation during training. For a participant with *m* observed timepoints, up to *m* ∗ (*m* − 1)/2 training samples can be generated by varying the input history range and the prediction timepoint. This increases training diversity and encourages the model to generalize across varying history lengths, prediction horizons, and individuals. For a more detailed discussion of these theoretical benefits, please refer to our earlier study (Zhang et al., 2025).

Building upon this framework, we consider four L2C-based model variants, which share the same longitudinal-to-cross-sectional formulation but differ in their downstream predictive models.

#### 2.6.1 L2C eXtreme Gradient Boost (L2C-XGB)

In prior study (Zhang et al., 2025), we evaluated two variants of L2C-based eXtreme Gradient Boosting (T. Chen & Guestrin, 2016): one using separate models for different prediction horizons (L2C-XGBw) and one using a single model across all future timepoints (L2C-XGBnw).

The latter consistently achieved superior performance in both within- and cross-dataset evaluations, suggesting that explicit modeling of forecast horizons may be unnecessary when temporal information is already encoded in L2C features (e.g., time since baseline and time since most recent measurement).

Motivated by these findings, we adopt L2C-XGBnw in this study, hereafter referred to as L2C-XGB for simplicity. To further improve predictive performance, we extend this approach to jointly model correlated regression targets, following the principles of multi-task learning. Specifically, we leverage the native multi-output regression capability in XGBoost (version 2.1.1) to train a single model for the joint prediction of MMSE and hippocampal volume. A separate model is trained for clinical diagnosis, as the current XGBoost package does not support mixed regression and classification tasks.

Four hyperparameters were tuned using Optuna (Akiba et al., 2019). The search space was shared across regression and classification models, except for the number of boosting rounds, which was tuned separately (Table 7).

**Table 7.**
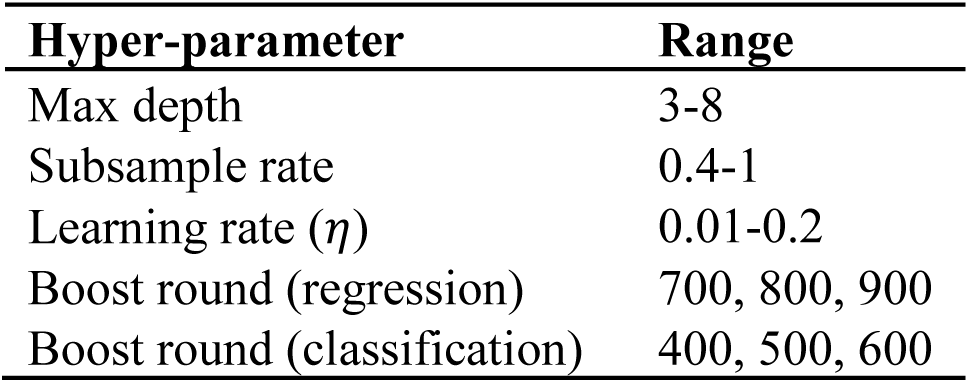
Hyperparameter search ranges for L2C-XGB. Hyperparameters were tuned independently on every validation fold via Optuna.

No additional feature imputation was performed, as XGBoost natively handles missing values by learning a default direction at each split during training, which determines how samples with missing feature values are routed through the tree. Furthermore, feature normalization was unnecessary, as tree-based models are invariant to input scaling. Because of its inherent robustness to sporadic missing data, L2C-XGB serves as a key baseline for evaluating whether such native handling remains effective under the more challenging setting of systemic missingness.

#### 2.6.2 L2C Fully-Connected Feedforward Neural Network (L2C-FNN)

The L2C-FNN model was included as a key neural network baseline, as this L2C-based approach demonstrated superior generalizability in our previous study. Like L2C-XGB, it adopts the L2C transformation and data augmentation framework (Section 2.6). In contrast to L2C-XGB, the FNN architecture enables a unified multi-task model that jointly predicts all three target variables, including both regression and classification tasks.

The model takes the same L2C features as input but requires additional preprocessing to accommodate FNN’s sensitivity to feature scaling and missing values. Specifically, numeric features were imputed using training-set medians and transformed via Gauss Rank normalization implemented using Scikit-learn’s quantile transformer (Pedregosa et al., 2011). Categorical features (e.g., APOE status, sex, most recent diagnosis) were one-hot encoded with an additional “unknown” category to represent missing values. Features with excessive missingness (>60%) were excluded, resulting in a 179-dimensional input vector (Table S7).

Missing data were handled entirely through preprocessing, without the use of advanced imputation techniques. The FNN consists of fully connected layers with LeakyReLU (Maas, 2013) activations and dropout (Srivastava et al., 2014) for enhanced generalizability. The output layer produces a 5-dimensional vector, comprising class probabilities for diagnosis and predictions for MMSE and hippocampal volume. Regression targets were Gauss Rank normalized during preprocessing, and the model was trained to predict values on this transformed scale.

The loss function combined cross-entropy for diagnosis with mean absolute error (MAE) for the regression targets, with equal weighting across tasks. Optimization was performed using stochastic gradient descent with momentum (Qian, 1999; Sutskever et al., 2013), and hyperparameters were tuned using Optuna (Akiba et al., 2019). The learning rate was scheduled via a CosineAnnealingWarmRestarts scheduler (Loshchilov & Hutter, 2017), with *T*_0_ = 20 and *T_mult_* = 2, resulting in a total of 140 training epochs across three restart cycles (20, 40, and 80 epochs). Full hyperparameter details are provided in Table 8.

**Table 8.** Hyperparameter search ranges for L2C-FNN. Hyperparameters were tuned independently on every validation fold via Optuna.

| Hyper-parameter | Range |
| --- | --- |
| Num of hidden layers | 1-6 |
| Size of hidden state | 128-768 |
| Dropout rate | 0-0.5 |
| LeakyReLU slope | 0.01, 0.05, 0.1 |
| L2 weight regularization | $(2^{-4} - 2^2) \times 10^{-3}$ |
| Learning rate | $2^{-9}$-$2^{-4}$ |

#### 2.6.3 L2C-FNN with Multiple Imputation by Chained Equations (L2C-MICE)

L2C-FNN (Section 2.6.2) adopts a simple impute-then-predict strategy, using median imputation for numeric features and an “unknown” category for categorical features. This naïve approach does not account for relationships among features and may be suboptimal under systemic missingness. To assess whether a more sophisticated imputation strategy can improve performance, we implemented L2C-MICE as an advanced baseline.

L2C-MICE uses Multiple Imputation by Chained Equations (MICE), an iterative regression-based approach that models each feature as a function of the others, thereby leveraging multivariate dependencies during imputation (Azur et al., 2011). We used the miceforest Python package^3^, which implements MICE using a LightGBM gradient-boosted tree backend for improved efficiency and scalability. All L2C features, including both numeric and categorical variables, were included in the imputation model.

We fixed the number of boosting iterations to 500. The remaining hyperparameters were tuned within each of the 20 ADNI training/validation/test splits described in Section 2.3. More specifically, for a given training/validation/test split, we ran the miceforest package’s built-in tuning procedure on the corresponding training set: this procedure performs an internal 10-fold cross-validation on the training set to score candidate hyperparameter configurations, and we allocated 50 optimization trials per split. Default search ranges were used for most parameters, except for the learning rate, which we searched over {0.001, 0.005, 0.01, 0.02, 0.05, 0.1}. The MICE model was then refit on the full training set using the best configuration and used to impute missing values in the corresponding training, validation, and test sets. Each imputation pass ran for 10 chained-equations iterations to allow convergence, producing fully imputed L2C feature sets with no remaining missing values.

We applied the same feature preprocessing pipeline as L2C-FNN, including Gauss Rank normalization for numeric features and one-hot encoding for categorical features (Section 2.6.2). A new L2C-FNN model was then trained from scratch on each imputed training fold using the same architecture, optimizer, learning rate scheduler, loss function, and hyperparameter tuning setup. Retraining is necessary because MICE-imputed feature distributions differ substantially from those of the original data, requiring re-optimization of model parameters.

#### 2.6.4 L2C Prior Fitted Network (L2C-PFN)

Tabular Prior Fitted Network (TabPFN) is a foundation model for tabular data that has demonstrated strong performance on small- to moderate-sized datasets (Hollmann et al., 2025). It is pre-trained on millions of synthetic tabular datasets generated from structural causal models (Pearl, 2009), which sample diverse data-generating processes, including varying level of feature interactions, noise, and missingness. Through this large-scale pretraining, TabPFN implicitly learns a rich prior over tabular prediction problems, enabling strong out-of-the-box generalization.

Unlike conventional fit–predict supervised learning approaches, TabPFN adopts an in-context learning paradigm (Brown et al., 2020). During pretraining, the model learns to predict targets for unlabeled samples by conditioning on a labeled “context” within synthetic datasets. At inference, this enables TabPFN to generalize to real-world data by conditioning on labeled training samples and predicting targets for unlabeled test samples in a single forward pass without parameter updates. Its transformer backbone applies two-way attention across both samples and features, preserving the relational structure of tabular data.

Here, we integrate TabPFN with the L2C framework by treating each L2C feature vector as one tabular row. For both within-ADNI and cross-dataset evaluation, the ADNI training partition served as the in-context labeled set, and TabPFN predicted targets for participants in the ADNI test partition (within-ADNI) or in the external cohorts (AIBL, MACC, OASIS). This procedure was repeated for each of the 20 ADNI splits described in Section 2.3, mirroring the evaluation protocol used for all other baselines. The L2C transformation is directly compatible with TabPFN’s input format, so no architectural changes or temporal priors were required (Dooley et al., 2023). Given TabPFN’s pretrained ability to handle missing and uninformative features, this combination serves as a key baseline for testing whether foundation-model-scale priors transfer to the systemic-missingness setting.

We used the open-source implementation^4^, employing “TabPFNClassifier” for clinical diagnosis and “TabPFNRegressor” for MMSE and hippocampal volume prediction. The three target variables were modeled independently. Default settings were used, with the exception of enabling the “ignore_pretraining_limits” option to bypass the default 10,000-sample constraint, as our training sets (with the L2C data augmentation) exceeded this limit. TabPFN internally handles missing values, categorical encoding, and feature normalization, so no additional preprocessing was applied. In contrast to L2C-FNN and L2C-MICE, all L2C features were retained regardless of their missingness ratio, as the model is designed to handle missingness robustly thanks to its pretraining.

As a pretrained foundation model, TabPFN does not involve conventional hyperparameter tuning. Instead, it employs an internal ensemble of predefined inference pipelines (four for classification and eight for regression), each corresponding to a different combination of preprocessing and postprocessing strategies. The predictions from these pipelines are averaged uniformly to produce the final output. We adopted this default ensemble configuration without further modification.

### 2.7 ProFuse-TTA

ProFuse-TTA is a two-stage hierarchical transformer that handles systemic missingness directly through self-attention, without explicit imputation. By operating on incomplete inputs, the model focuses on available and informative biomarkers and avoids error propagation from unreliable data completion. An overview of the full architecture is provided in Figure 2.

**Figure 2.**
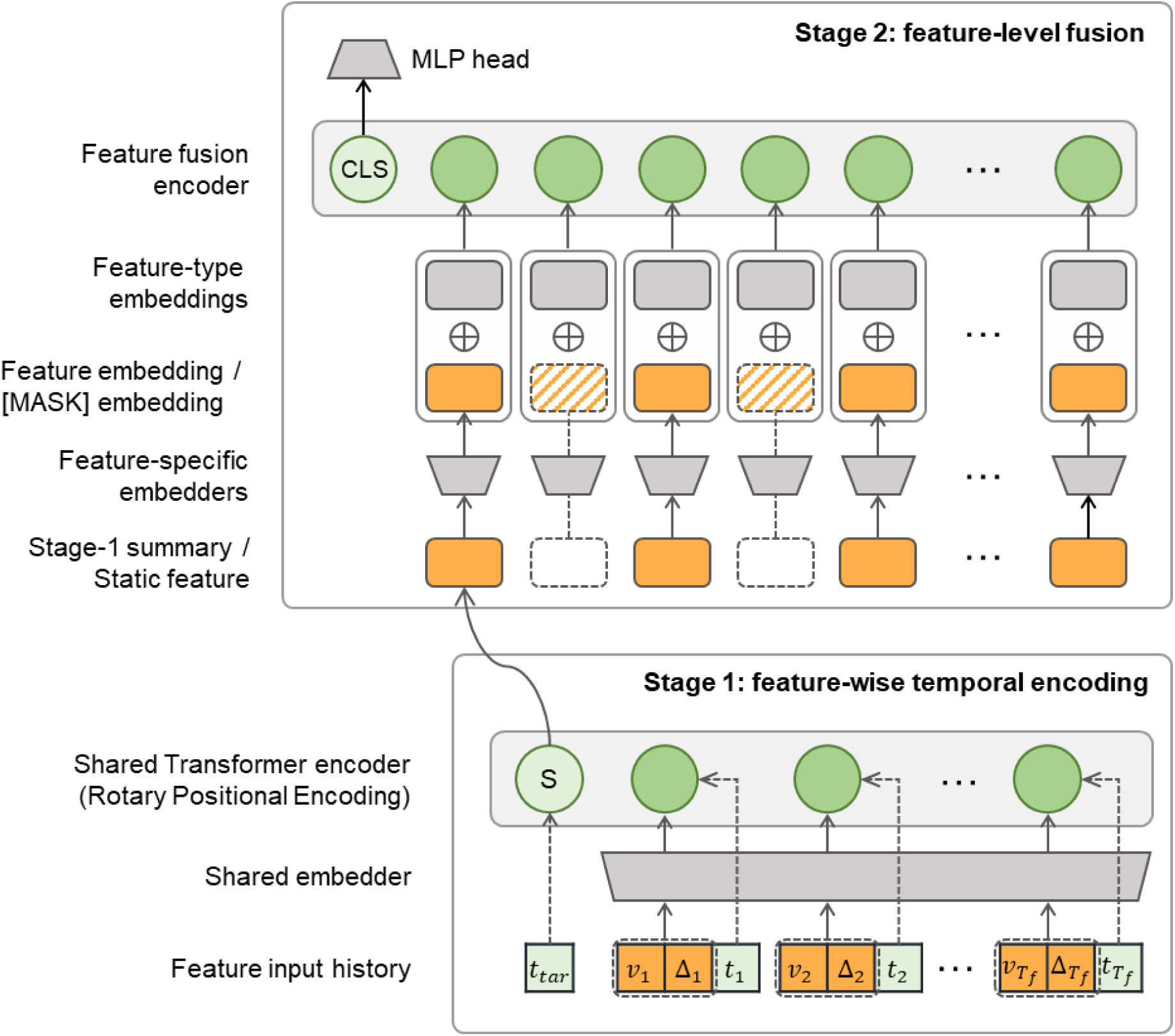
Architecture of ProFuse-TTA. The model consists of two stages. Stage 1 encodes each dynamic feature’s longitudinal history independently into a feature-level representation. Each observed visit becomes a token via an MLP embedder, and a learnable [SUMMARY] token is prepended to the sequence. Rotary positional encoding (RoPE) carries timing information, and a transformer encoder aggregates the visits. The final hidden state of the [SUMMARY] token is the Stage 1 representation for that feature. For clarity, only one dynamic feature is shown. Stage 2 fuses the Stage 1 representations with static features (e.g., demographics) using a second transformer encoder, where each token represents a feature rather than a timepoint. Each token is the sum of an MLP-encoded representation of the feature and a learnable feature-type embedding that identifies which feature it represents. Systemically missing features are represented by a shared learnable [MASK] embedding in place of the MLP output. A learnable [CLS] token is prepended; its final hidden state is passed through a BertPooler and an MLP prediction head, producing the predictions for clinical diagnosis, MMSE, and hippocampal volume at the target timepoint.

Training and inference follow the L2C data augmentation scheme (Section 2.6), where each sample pairs an observed history with a future target timepoint. Unlike the other L2C variants, which operate on engineered L2C feature vectors, ProFuse-TTA models the raw time series directly, enabling end-to-end learning.

The hierarchical architecture decouples temporal modeling from multimodal fusion, allowing it to capture both longitudinal dynamics and cross-modal interactions. Stage 1 (Section 2.7.1) processes each biomarker time series independently with a transformer encoder, treating timepoints as tokens to capture progression of individual biomarkers. Stage 2 (Section 2.7.2) treats the Stage 1 representations and static features as tokens and fuses them with a second transformer encoder to predict the target timepoint.

#### 2.7.1 Stage 1: Feature-wise temporal encoding

In Stage 1, ProFuse-TTA encodes the longitudinal history of each dynamic feature independently into a fixed-dimensional representation (Figure 2). Suppose a participant has *T_f_* observed visits for feature *f*. We denote this history as

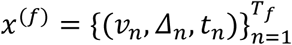

where *v_n_* is the feature value at visit n, Δ*_n_* is the number of months between that visit and the prediction target timestamp, and *t_n_* is the number of months between that visit and baseline.

A key design choice is that each feature is encoded independently, rather than within a feature-by-visit grid. A feature-by-visit grid is problematic because different features are typically missing at different visits, forcing the model to either drop entire visits or impute the missing entries. Encoding each feature independently sidesteps this issue: each feature has its own sequence over the visits at which it was observed, with no need to align across features or fill in missing entries. This stage can therefore be viewed as a learnable replacement for the L2C transformation (Section 2.6), in which the engineered summary statistics of L2C are replaced by a learnable encoder whose parameters are shared across features. The shared encoder keeps the parameter count manageable and lets the model learn temporal patterns common across biomarkers. Feature identity is introduced later, in Stage 2, where the per-feature representations are fused together.

Δ*_n_* tells the model how far each observed visit is from the prediction target. This is important because the same observed history can be used to predict different future timepoints under the L2C data augmentation scheme (Section 2.6), and the model needs to produce different predictions depending on the forecast horizon. For example, if a participant has visits at month 0 and month 12, the model should output different predictions when asked to forecast month 24 versus month 60. Both Δ*_n_* and the feature value *v_n_* are z-normalized using mean and variance computed from the ADNI training set (before L2C-style augmentation).

We use *t_n_* separately as the position index for rotary positional encoding (RoPE; Su et al., 2023). RoPE encodes how far apart any two observations are in time, which the self-attention mechanism uses to model temporal relationships between visits. Unlike Δ*_n_*, we do not z-normalize *t_n_*. The reason is that RoPE was designed for non-negative integer positions, such as token positions in a text sequence, and its preset rotation frequencies assume position indices on that scale. Months-since-baseline naturally fits this mold, but z-normalized values do not: they would all map to small floating-point numbers centered near zero, compressing the rotation angles into a narrow range and making different positions indistinguishable to the attention mechanism. To handle irregular sampling, we round *t_n_* to integer months for use as the RoPE position index.

The pair (*v_n_*, *Δ_n_*) is projected into a token embedding using an MLP that is shared across all timepoints and features. We also prepend a learnable [SUMMARY] token to the sequence and assign it the prediction target timestamp *t_tar_* as its position index. Because RoPE encodes pairwise distances, the attention between [SUMMARY] and each observation is modulated by *t_tar_* − *t_n_*, the number of months between the observation and the prediction target. This allows the model to aggregate the observed history in a way that is conditioned on the prediction target. The final hidden state of the [SUMMARY] token becomes the Stage 1 representation for feature *f*, which is then passed to Stage 2.

#### 2.7.2 Stage 2: Feature-level fusion

In Stage 2, each token represents a single feature. Dynamic features are represented by their Stage 1 encodings, while static demographic features (e.g., sex, marital status) and scalar features such as age at the prediction timepoint are passed directly to Stage 2 without going through Stage 1. This contrasts with most transformer-based time-series models, where each token represents a timepoint. In our study, this results in 28 feature tokens (Table 1).

To improve robustness under systemic missingness, we adopt a three-part embedding strategy that separates feature content, feature identity, and missingness information. First, each feature token is passed through its own MLP encoder, which projects it into a shared hidden dimension and lets the model learn a specialized transformation per feature.

Second, a learnable feature-type embedding is added to each token to signal its feature identity (e.g., MMSE, hippocampal volume). Unlike standard transformers, no positional encoding is used, since the ordering of features is not inherently meaningful.

Third, for an observed feature, the final token embedding is the sum of its MLP-encoded representation and its feature-type embedding. For a missing feature, the MLP output is replaced by a shared learnable [MASK] embedding, which is combined with the corresponding feature-type embedding. This ensures that the model is informed both that a feature is missing and which feature is absent.

Following the BERT framework (Devlin et al., 2019), a learnable [CLS] token is prepended to the sequence of feature tokens. The final hidden state of the [CLS] token serves as an aggregated representation of all features and is passed through a BertPooler module. The pooled representation is then fed into a multi-task MLP prediction head, producing a 5-dimensional output vector: three logits for clinical diagnosis (CN, MCI, DEM) and two regression outputs for MMSE and hippocampal volume.

#### 2.7.3 ProFuse-TTA implementation details

We track which features are systemically missing using a binary mask of shape #samples × #features. A dynamic feature is marked as missing only if its entire longitudinal trajectory is unavailable. This mask is not fed to the transformer as input, but is used internally to decide when a feature representation should be replaced by the shared [MASK] embedding (Section 2.7.2).

To improve robustness to systemic missingness, we randomly mask additional features during training and validation. At each iteration, a random binary mask is combined with the systemic missingness mask described above, so that features already missing remain missing and additional features are masked on top. Features collected together in practice are masked together. For example, the brain ROIs come from a single MRI scan and are masked as a group. Different groups are masked independently. During training, the masking ratio is treated as a hyperparameter (ranging from 0.1 to 0.5 in Table 9). During validation, the masking ratio is uniformly sampled from [0.1, 0.4] per example to expose the model to varying levels of missingness.

**Table 9.** Hyper-parameters and search ranges for ProFuse-TTA, tuned on the validation set using Optuna. The encoder hidden size is the product of the number of attention heads and the head embedding size. The FFN layer hidden size is the product of the encoder hidden size and the FFN multiplier.

| Hyper-parameter | Range |
| --- | --- |
| Number of Stage 1 layers | 1, 2 |
| Number of Stage 1 attention heads | 2, 4 |
| Stage 1 head embedding size | 32, 64 |
| Stage 1 FFN multiplier | 2, 3, 4 |
| Number of Stage 2 layers | 2, 3, 4 |
| Number of Stage 2 attention heads | 4, 6, 8 |
| Stage 2 head embedding size | 32, 64 |
| Stage 2 FFN multiplier | 2, 3, 4 |
| Encoder dropout rate (shared between two stages) | 0-0.5 |
| MLP dropout rate | 0-0.5 |
| MLP hidden state size | 64, 128, 256 |
| L2 weight regularization | $(2^{-4}-2^2) \times 10^{-3}$ |
| Learning rate | $2^{-9}-2^{-4}$ |
| Modality masking ratio | 0.1-0.5 |

Both ProFuse-TTA stages are implemented using HuggingFace: RoFormerEncoder^5^ for Stage 1 and the BertEncoder^6^ for Stage 2. ProFuse-TTA is trained using the same optimizer, learning rate scheduler, and loss functions as L2C-FNN. Regression targets are z-normalized using statistics from the ADNI training set. ProFuse-TTA-specific hyperparameters and search ranges are summarized in Table 9.

#### 2.7.4 Test time adaption with history-aware loss-gating

Test-time adaptation (TTA) has emerged as a way to calibrate models per-instance using the test data itself (D. Wang et al., 2021; J. Liang et al., 2024), but has been little explored in longitudinal disease progression prediction. To improve robustness at test time, we propose a TTA that introduces lightweight per-task adapters on top of the frozen base model. The base model weights are kept fixed so that the population-level disease patterns learned during training are preserved, while the adapters allow individual-level calibration. TTA is applied at evaluation time on both within-ADNI test participants and cross-dataset test participants. Distribution shifts can arise in both settings — across datasets due to differences in cohorts and protocols, and within ADNI due to individual variability — so adaptation is beneficial in either case.

For a test individual with *T* observed timepoints, we construct *T* − 1 adaptation samples by treating each observed timepoint as a held-out prediction target, using all earlier timepoints as context. In other words, for each timepoint *t* ∈ {2,…, *T*}, the model uses observations from 1 to *t* − 1 as context to predict the target variables at timepoint *t*. The adapters are trained on these

*T* − 1 samples to learn an individual-specific adjustment, which is then applied to predictions of the future trajectory. For individuals with fewer than two observed timepoints, no adaptation samples can be constructed, and predictions are obtained directly from the base model.

We use a separate adapter for each prediction target (MMSE, hippocampal volume, clinical diagnosis) to prevent gradient interference between tasks.

##### Regression adapter (MMSE and hippocampal volume)

For regression targets, we use a history-aware adapter that produces a correction δ based on the base prediction *y_base_* and the patient’s historical minimum *y_min_*:

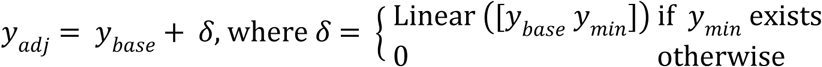

where “Linear” is a linear combination of two scalars *y_base_* and *y_min_*, together with an intercept. Anchoring the correction to the historical minimum is motivated by the fact that MMSE and hippocampal volume both decline monotonically with disease progression. The historical minimum therefore acts as a meaningful floor: a patient’s true future value is unlikely to be much higher than what they have already exhibited. The adapter learns to correct the base prediction toward this floor when the base model over- or under-estimates relative to the patient’s observed trajectory. If no historical observations exist for a given target variable, the adapter outputs zero correction and the base prediction is used directly. The linear layer weights and biases are initialized to zero so that adaptation starts from the base model’s prediction.

The regression loss for each adaptation sample is the MAE between *y_adj_* and the observed target, weighted by a per-sample gate. Since all regression targets are z-normalized using ADNI training set statistics, the raw MAE loss *L_raw_* represents the prediction error in units of standard deviations. The per-sample loss weight is defined as

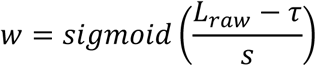

with threshold τ = 1.0 and scale *s* = 0.3. To interpret these hyperparameters, when the prediction error is within 1σ — a range plausibly attributable to measurement variability or label noise — the weight stays small and the adapter remains largely inactive. As the error exceeds 1σ, the weight rises toward 1, and the adapter engages. This prevents the adapter from overfitting to limited patient history on samples where the base model is already well-calibrated, and focuses adaptation on samples that show real distribution shift.

##### Diagnosis adapter (clinical diagnosis)

For the 3-class clinical diagnosis task, we use a Gated Calibration Module (GCM; Kim et al., 2025) to refine the base model’s logits:

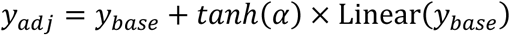

where *α* is a learnable scalar initialized to 0. Since *tanh*(0) = 0, the correction term vanishes at the start of adaptation, and the adapter learns to scale up the correction as needed.

Unlike the regression adapter, the diagnosis loss is an unweighted cross-entropy. No per-sample gating is needed because cross-entropy is self-gating by construction: its gradient with respect to the logits is proportional to (*p* − *y*), which naturally diminishes as the predicted probability *p* approaches the true label *y* (Goodfellow et al., 2016).

##### Optimization

Adaptation is performed per-individual, treating the *T* − 1 samples as a single batch. The final loss is the mean of the weighted (regression) and unweighted (diagnosis) per-sample losses. We use a stochastic gradient descent (SGD) optimizer with an initial learning rate of 5 × 10^−3^ and a cosine annealing scheduler with three warm restart cycles (intervals of 20, 40, and 80 epochs). We apply weight decay (1 × 10^−4^) for regularization. Because all adapters are zero-initialized, weight decay pulls the updates toward identity mappings, so the adapters only provide the minimal correction required.

### 2.8 Evaluating robustness to systemic missingness

A central goal of this study was to evaluate model robustness to systemic missingness, where a biomarker present in the training data is completely absent from a test dataset. We assessed this in two complementary ways.

First, to isolate the effect of systemic missingness from general domain shifts, we performed a controlled, within-cohort ablation study using ADNI. For each of the 20 ADNI test sets, we first identified the subset of participants with at least one observed value for every biomarker across all input timepoints, which we refer to as the “full modality set” baseline. Because only a subset of test participants is included, the results of this analysis are not directly comparable to the main within-ADNI evaluation results (Section 3.1).

We then generated five modified versions of this baseline by ablating one modality group at a time (Table 1): MRI features (MRI), cognitive features (COG), diagnostic features (DX), PET features (PET), or CSF features (CSF). In each version, all biomarkers belonging to the ablated modality group were set to missing (NaN), while the remaining biomarkers were left unaltered. We quantified resilience by the performance drop relative to the full modality set baseline under each ablation. While all algorithms were expected to experience performance decline, a robust algorithm should maintain a reasonable drop under systemic missingness.

Second, our cross-dataset evaluation provided a complementary, real-world test of robustness. The external test sets (AIBL, MACC, and OASIS) lacked numerous biomarkers present in the ADNI training data (Table 1). This evaluation, therefore, assesses model generalizability under the combined, real-world challenge of systemic missingness and domain shift. Together, the two analyses isolate and combine the two sources of distribution shift, respectively.

### 2.9 Further cross-cohort analyses

We performed two additional analyses to compare the effectiveness of ProFuse-TTA against the baseline models – MinimalRNN, AD-Map, and four L2C-based variants.

#### 2.9.1 Impact of number of observed timepoints on cross-cohort prediction performance

Early detection of AD dementia is most useful when a model can produce reliable predictions from limited history. We therefore tested the performance of the seven models on the external datasets when given only 1, 2, 3, or 4 input timepoints per participant. This differed from the main benchmarking analysis (Section 3.3), in which each model was given half of a participant’s timepoints as input.

The set of available test participants differed across the four conditions because participants varied in the number of timepoints they had. In OASIS, we kept only test participants with at least 4 input timepoints, so the same group was used across all four conditions. In AIBL and MACC, the maximum number of input timepoints per participant was 2 and 3 respectively, so the 3- and 4-timepoint conditions were not possible for AIBL, and the 4-timepoint condition was not possible for MACC. The test participant sets in this analysis therefore differ from those of Section 3.3, and the results are not directly comparable.

#### 2.9.2 Breakdown of cross-cohort prediction in yearly intervals

To understand how prediction performance changes with forecast horizons, we broke down the cross-cohort results into yearly intervals up to 6 years into the future. Each participant’s future timepoints were categorized into yearly intervals based on elapsed time from the last input timepoint. For example, given a participant with 10 timepoints, if the last input timepoint (5th timepoint) was at month 60 and the 6th timepoint was at month 70, the 10-month gap places the prediction at the 6th timepoint in the 0-1 year bin.

All models are expected to perform worse as the horizon grows. The question is how gracefully each model’s performance degrades: a robust algorithm should still produce useful predictions several years into the future.

### 2.10 Ablation study of history-aware loss-gated adaptation

To evaluate the contribution of our adaptation strategy, we performed an ablation study comparing the proposed TTA strategy against the non-adapted base model and two global fine-tuning variants (base-heavy and base-light). Unlike the proposed strategy, which only optimizes lightweight task-specific adapters, the global variants update the entire base model.

The base-heavy variant tunes the entire network for 140 epochs using SGD with an initial learning rate of 1×10⁻³ and a cosine annealing scheduler with three warm restart cycles (intervals of 20, 40, and 80 epochs). Because the limited per-individual longitudinal data may cause overfitting, we also tested a more conservative base-light variant, with a reduced initial learning rate of 1×10⁻⁴ over 60 epochs (two warm restart cycles). Unlike our proposed TTA strategy, neither global variant uses weight decay. Our proposed strategy can apply weight decay because its adapters are zero-initialized, so weight decay pulls them toward an identity mapping. The global variants, by contrast, start from the pretrained weights, so weight decay would pull those weights toward zero and erase what the base model had learned.

The global variants also differ from the proposed strategy in two other ways. First, in both base-heavy and base-light variants, the three target variables are predicted by a shared multi-task MLP head with equally weighted losses, rather than by task-specific adapters. This coupling might cause gradient interference, where the training signal is dominated by tasks with larger raw loss magnitudes (e.g., MMSE), potentially degrading performance for targets already well-predicted by the base model (e.g., hippocampal volume). Second, the proposed strategy modulates each sample’s regression loss based on its current prediction error, so the gating shrinks as the adapter improves on a sample. The global variants weight all samples equally throughout adaptation.

### 2.11 Deep neural network implementation

MinimalRNN, L2C-FNN, L2C-MICE, L2C-PFN, and ProFuse-TTA were implemented in Python 3.9.19 using PyTorch 2.3.0 (Paszke et al., 2019) and executed on NVIDIA RTX 3090 GPUs with CUDA 12.1.

### 2.12 Performance evaluation and statistical tests

As a reminder, we used a 20-fold cross-validation procedure to train each model (MinimalRNN, AD-Map, the four L2C variants, and ProFuse-TTA) on ADNI, and applied the trained models to predict clinical diagnosis, MMSE, and hippocampal volume in the ADNI test sets (within-ADNI evaluation) and in three external datasets (cross-dataset evaluation). This section describes the evaluation metrics and statistical procedures used to compare model performance.

Clinical diagnosis was evaluated using the multiclass area under the ROC curve (mAUC; Hand & Till, 2001), following the TADPOLE challenge. mAUC was computed as the average of three two-class AUCs (DEM vs. not DEM, MCI vs. not MCI, and CN vs. not CN). Higher mAUC indicates better performance. Because mAUC is a group-level metric, predictions were pooled across all participants and all future timepoints before computing a single value per test set.

MMSE and hippocampal volume predictions were evaluated using mean absolute error (MAE), following the TADPOLE challenge. Lower MAE indicates better performance. MAE was computed per participant by averaging the absolute errors across all forecast timepoints, yielding one MAE value per participant in the test set.

For the within-ADNI evaluation, the 20-fold cross-validation produced 20 mAUC values, 20 MMSE MAE values, and 20 hippocampal volume MAE values (each MAE averaged across participants within a fold). The per-fold metrics are not independent across folds, so we used the corrected resampled t-test (Nadeau & Bengio, 2003; Zeng et al., 2026) to compare algorithms. Separate tests were performed for clinical diagnosis, MMSE, and hippocampal volume.

For the cross-dataset evaluation, the final performance was computed by averaging the performance metrics across 20 trained models of each algorithm. To compare algorithms, we used paired-sample t-test for MMSE and hippocampal volume (Zeng et al., 2026). For mAUC, which is a group-level metric, we used a paired (“sign-flip”) permutation test (Zeng et al., 2026). All statistical tests were two sided. Figures S3 and S4 illustrate the t-test and permutation test, respectively.

Multiple comparisons were corrected using a false discovery rate (FDR) of q < 0.05 (Benjamini & Hochberg, 1995) for both within-ADNI and cross-cohort evaluations.

## 3 Results

### 3.1 L2C-PFN and ProFuse-TTA perform the best for within-ADNI prediction

Figure 3 and Table 10 compare the prediction performance of MinimalRNN, AD-Map, four L2C variants (L2C-XGB, L2C-FNN, L2C-MICE, L2C-PFN), and ProFuse-TTA for within-cohort (ADNI) prediction of MMSE, hippocampus volume, and clinical diagnosis. Because the focus of this study is robustness under systemic missingness and cross-cohort generalization, we treat the within-ADNI results primarily as a sanity check that ProFuse-TTA remains competitive in the in-domain setting before turning to the harder evaluations in Sections 3.2 and 3.3.

**Figure 3.**
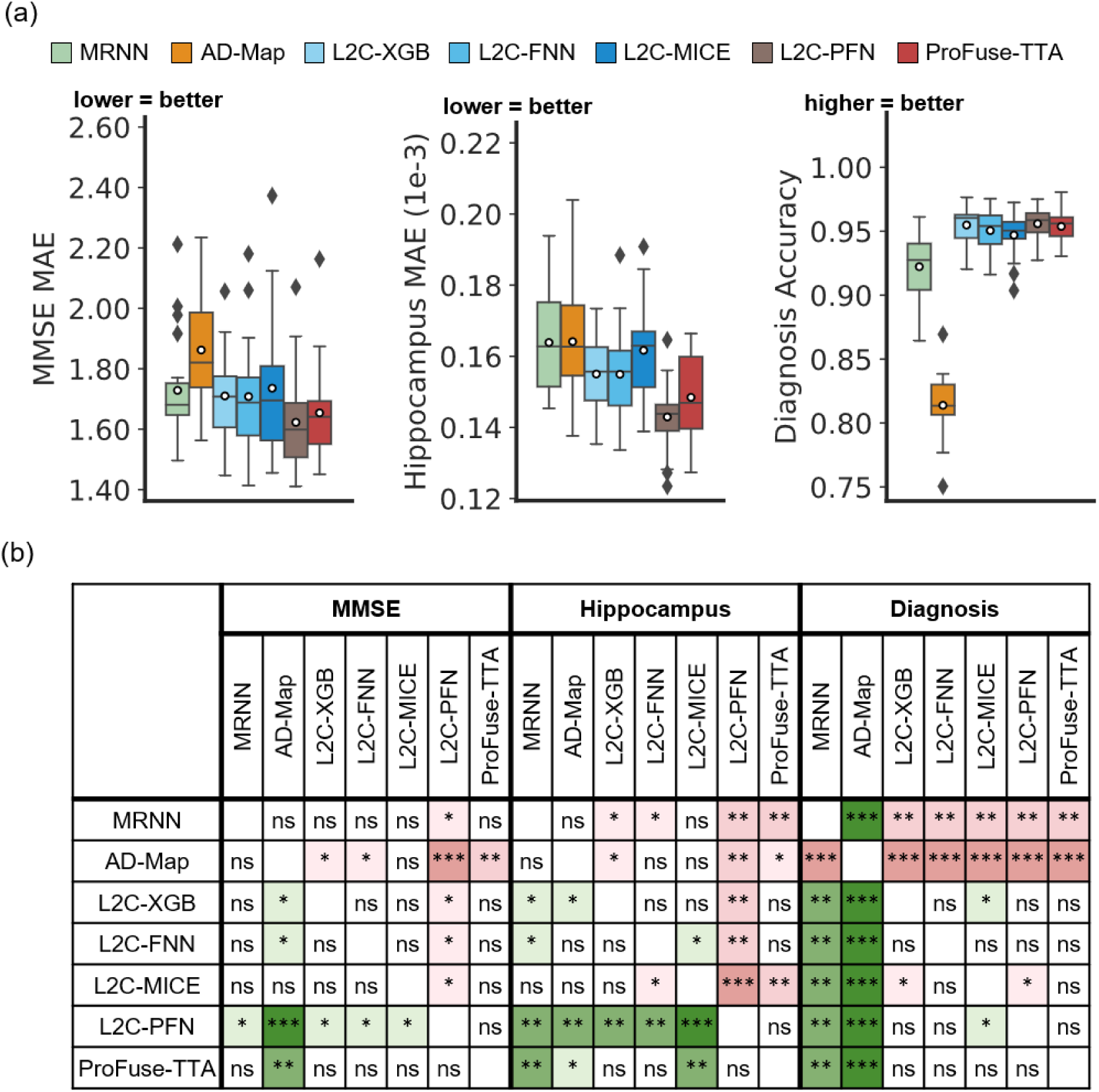
Within-cohort (ADNI) prediction performance. (a) Boxplots represent variability in prediction performance across 20 test folds for (left) MMSE, (middle) hippocampus volume and (right) clinical diagnosis predictions. (b) Statistical difference between all models. “***” indicates p < 0.00001 and statistical significance after multiple comparison correction (FDR q < 0.05). “**” indicates p < 0.001 and statistical significance after multiple comparison correction (FDR q < 0.05). “ns” indicates no statistical significance (p ≥ 0.05) or did not survive FDR correction.

**Table 10.**
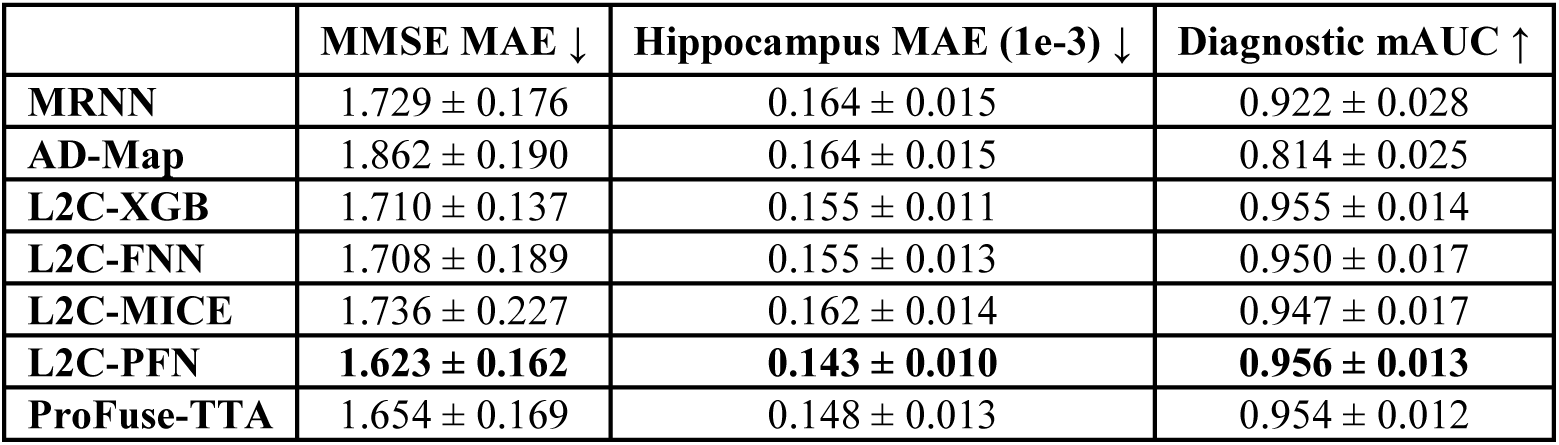
Within-cohort (ADNI) prediction performance averaged across 20 test folds. . For clinical diagnosis, ↑ implies that higher mAUC indicates better performance. For MMSE and hippocampus volume, ↓ implies that lower MAE indicates better performance. The best result for each performance metric was bolded.

|  | MMSE MAE ↓ | Hippocampus MAE (1e-3) ↓ | Diagnostic mAUC ↑ |
| --- | --- | --- | --- |
| **MRNN** | 1.729 ± 0.176 | 0.164 ± 0.015 | 0.922 ± 0.028 |
| **AD-Map** | 1.862 ± 0.190 | 0.164 ± 0.015 | 0.814 ± 0.025 |
| **L2C-XGB** | 1.710 ± 0.137 | 0.155 ± 0.011 | 0.955 ± 0.014 |
| **L2C-FNN** | 1.708 ± 0.189 | 0.155 ± 0.013 | 0.950 ± 0.017 |
| **L2C-MICE** | 1.736 ± 0.227 | 0.162 ± 0.014 | 0.947 ± 0.017 |
| **L2C-PFN** | **1.623 ± 0.162** | **0.143 ± 0.010** | **0.956 ± 0.013** |
| **ProFuse-TTA** | 1.654 ± 0.169 | 0.148 ± 0.013 | 0.954 ± 0.012 |

To simplify the interpretation of the pairwise statistical tests (Figure 3b), Table 11 summarizes the relative ranking of all models on each prediction task. Each model receives a score equal to the number of algorithms it statistically outperforms minus the number that statistically outperform it. For example, on MMSE prediction, L2C-XGB was statistically better than one algorithm and worse than one, giving it a score of 1 − 1 = 0. AD-Map was worse than four and better than none, giving −4. Ranks are then assigned by sorting these scores (1 = best, 7 = worst, ties allowed). The “overall” column in Table 11 averages ranks across the three tasks, so the best possible overall rank is 1 (first place on all three).

**Table 11.** Within-cohort (ADNI) prediction performance rankings of all models. Each row shows the rank of a model for three evaluation metrics: MMSE MAE, hippocampus MAE, and diagnostic mAUC (1 = best, 7 = worst; ties allowed). Rankings were derived by summing wins (+1) and losses (–1) for each model, as indicated by green (win) and red (loss) in the statistical significance tables (Figure 3). The model with the highest total score receives rank 1 for that metric. More details can be found in Section 3.1. The “overall” column in Table 11 shows the overall ranking by averaging the rankings across all three prediction tasks (MMSE, hippocampus, diagnosis). The best possible overall ranking was 1, corresponding to first-place performance in all three tasks. L2C-PFN achieved this optimal score, followed by ProFuse-TTA and L2C-XGB.

|  | MMSE MAE | Hippocampus MAE | Diagnostic mAUC | Overall |
| --- | --- | --- | --- | --- |
| MRNN | 5 | 7 | 6 | 6 |
| AD-Map | 7 | 5 | 7 | 6.3 |
| L2C-XGB | 3 | 3 | 1 | 2.3 |
| L2C-FNN | 3 | 3 | 3 | 3 |
| L2C-MICE | 5 | 5 | 5 | 5 |
| L2C-PFN | 1 | 1 | 1 | 1 |
| ProFuse-TTA | 2 | 2 | 3 | 2.3 |

L2C-PFN achieved this optimal overall rank of 1, ranking first on every task and confirming TabPFN’s strong in-domain capability on tabular data. The next best models were ProFuse-TTA and L2C-XGB with an overall rank of 2.3. Although ProFuse-TTA and L2C-XGB have the same overall ranking, we note that L2C-PFN was statistically better than L2C-XGB for predicting MMSE and hippocampus volume, while being statistically equivalent for clinical diagnosis prediction (Figure 3b). On the other hand, L2C-PFN and ProFuse-TTA were statistically equivalent in all three prediction tasks. Therefore, we conclude that L2C-PFN and ProFuse-TTA performed the best for within-ADNI prediction.

### 3.2 ProFuse-TTA is the most robust to within-ADNI modality ablation

Tables S11 to S13 report the performance drop for each model under each modality ablation, computed relative to the model’s full-modality-set baseline (Section 2.8). For each task, the largest average performance drop across all seven models came from ablating the modality containing the target variable itself: ablating cognition (COG) increased MMSE MAE by 67% on average, ablating MRI increased hippocampus MAE by 256%, and ablating clinical diagnosis (DX) reduced diagnostic mAUC by 6.1% (Tables S11 to S13). We refer to these as the "key modality" for each task.

FIGURE 4 visualizes the performance drop. Figure S5 reports the pairwise statistical tests, and Table 12 summarizes the resulting rankings using the same ranking scheme as Section 3.1. Based on Table 12 (and Figure S5), ProFuse-TTA ranked first in 14 of 15 ablation scenarios, achieving the best overall ranking of 1.1. In particular, ProFuse-TTA was ranked first in every scenario where the key modality for a task was removed (COG for MMSE, MRI for hippocampus, DX for diagnosis), indicating that it degraded the least when the most informative modality was missing.

**Figure 4.**
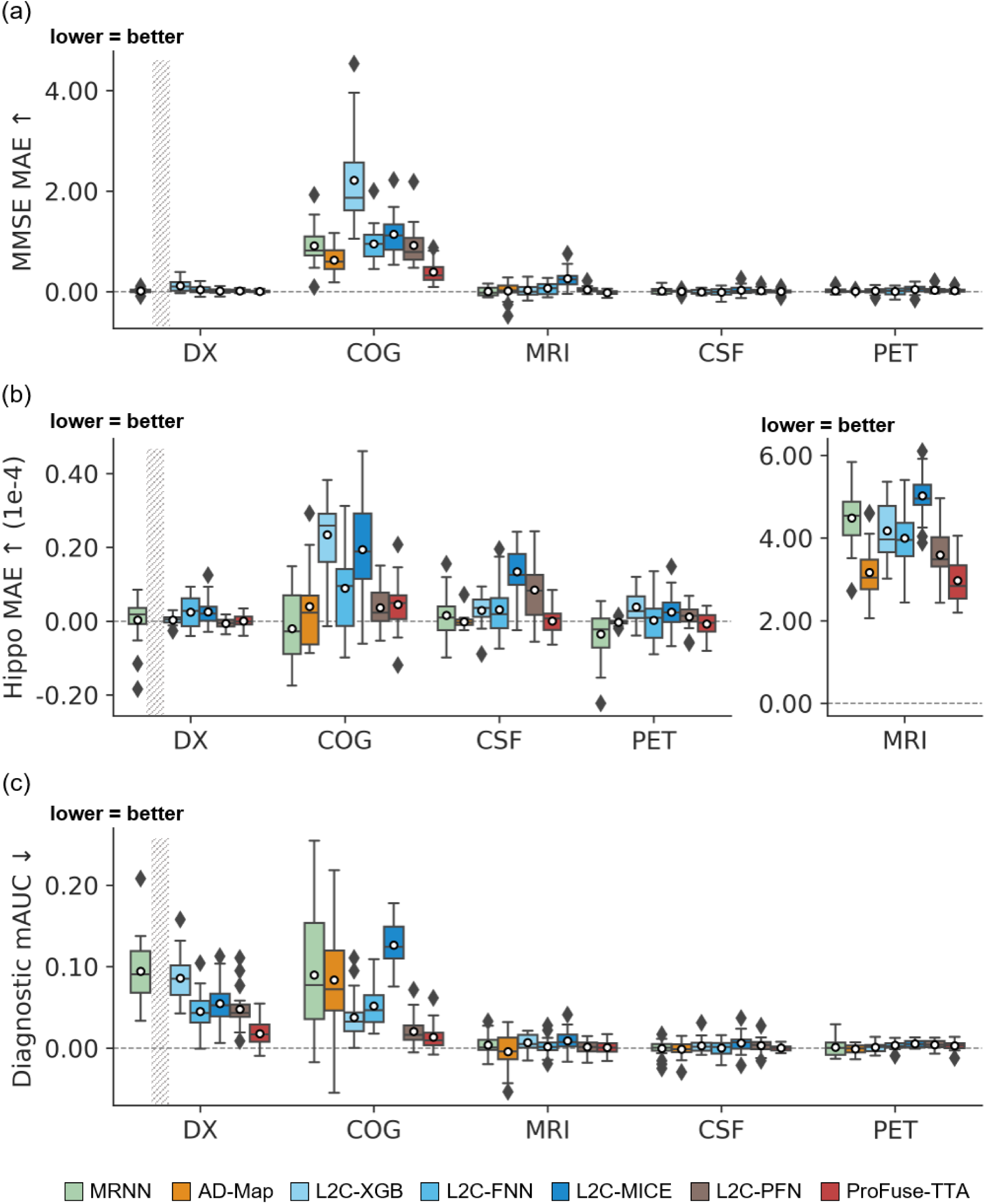
Within-cohort (ADNI) prediction performance drops under different modality ablation scenarios. (a) Boxplots illustrate the variability across 20 trained models (obtained from training with all data in ADNI) for MMSE prediction performance drops under each modality ablation scenario. The x-axis denotes the ablated modality ("DX": ablate diagnosis; "COG": ablate cognition; "MRI": ablate MRI; "CSF": ablate CSF; "PET": ablate PET). The y-axis shows the increase in MAE relative to the "full modality set" baseline (Section 2.8). Due to model design, AD-Map does not utilize diagnostic input features and therefore has no results in the "Ablate DX" condition, which is marked as "N.A." using striped bands. (b) Same as (a) but for hippocampus volume prediction (MAE). MRI ablation drops are shown in a separate panel on the right due to their larger magnitude. (c) Same as (a) but for diagnostic mAUC. The y-axis shows the decrease in mAUC relative to the baseline. Across all three panels, lower values indicate better robustness.

**Table 12.**
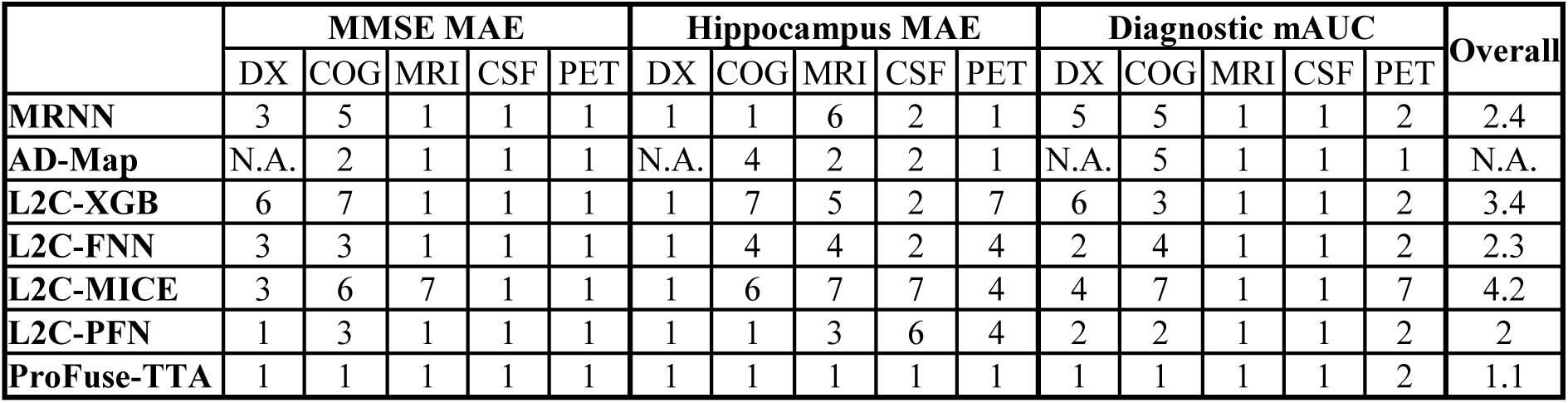
Within-cohort (ADNI) prediction performance drop rankings under modality ablation. Each row shows the rank of a model for MMSE MAE, hippocampus MAE, and diagnostic mAUC under five ablation scenarios (1 = best, 7 = worst; ties allowed). Rankings were derived by summing wins (+1) and losses (–1) for each model, as indicated by green (win) and red (loss) in the statistical significance tables (Figure S5). The model with the highest total score receives rank 1 for that metric under each modality ablation scenario. The “overall” column averages ranks across all scenarios, so the best possible overall ranking is 1 (first place in every scenario). AD-Map does not use clinical diagnosis as input (Section 2.5), so its "DX" ranks and overall average are marked "N.A.". With an overall ranking of 1.1, ProFuse-TTA performed the best.

|  | MMSE MAE |  |  |  |  | Hippocampus MAE |  |  |  |  | Diagnostic mAUC |  |  |  |  | Overall |
| --- | --- | --- | --- | --- | --- | --- | --- | --- | --- | --- | --- | --- | --- | --- | --- | --- |
|  | DX | COG | MRI | CSF | PET | DX | COG | MRI | CSF | PET | DX | COG | MRI | CSF | PET |  |
| MRNN | 3 | 5 | 1 | 1 | 1 | 1 | 1 | 6 | 2 | 1 | 5 | 5 | 1 | 1 | 2 | 2.4 |
| AD-Map | N.A. | 2 | 1 | 1 | 1 | N.A. | 4 | 2 | 2 | 1 | N.A. | 5 | 1 | 1 | 1 | N.A. |
| L2C-XGB | 6 | 7 | 1 | 1 | 1 | 1 | 7 | 5 | 2 | 7 | 6 | 3 | 1 | 1 | 2 | 3.4 |
| L2C-FNN | 3 | 3 | 1 | 1 | 1 | 1 | 4 | 4 | 2 | 4 | 2 | 4 | 1 | 1 | 2 | 2.3 |
| L2C-MICE | 3 | 6 | 7 | 1 | 1 | 1 | 6 | 7 | 7 | 4 | 4 | 7 | 1 | 1 | 7 | 4.2 |
| L2C-PFN | 1 | 3 | 1 | 1 | 1 | 1 | 1 | 3 | 6 | 4 | 2 | 2 | 1 | 1 | 2 | 2 |
| ProFuse-TTA | 1 | 1 | 1 | 1 | 1 | 1 | 1 | 1 | 1 | 1 | 1 | 1 | 1 | 1 | 2 | 1.1 |

### 3.3 ProFuse-TTA performed the best for cross-cohort prediction

Figures 5 and 6 show the prediction performance of the seven models for cross-cohort MMSE, hippocampus volume and clinical diagnosis prediction in three external datasets (AIBL, MACC, and OASIS). Numerical values are reported in Table 13, and Table 14 summarizes the model rankings using the same scoring scheme as Section 3.1.

**Figure 5.**
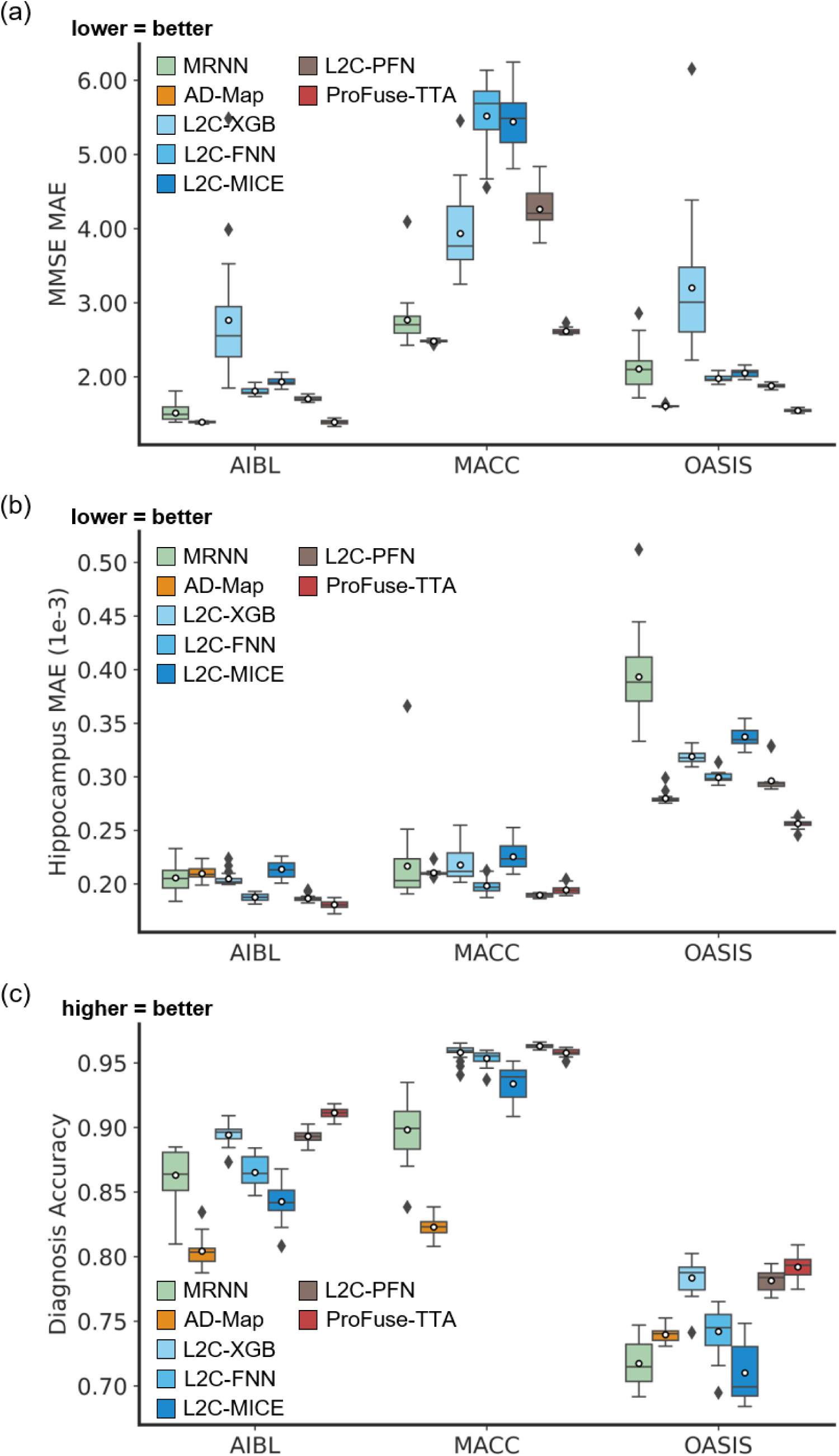
Cross-cohort prediction performance on three external test datasets. (a) Boxplots illustrate the variability across 20 trained models (from ADNI) for MMSE prediction assessed using MAE. The x-axis denotes the test dataset used for evaluation. Lower MAE indicates better performance. (b) Same as (a) but for hippocampus volume prediction (MAE). (c) Same as (a) but for diagnosis accuracy (mAUC). Higher mAUC indicates better performance.

**Figure 6.**
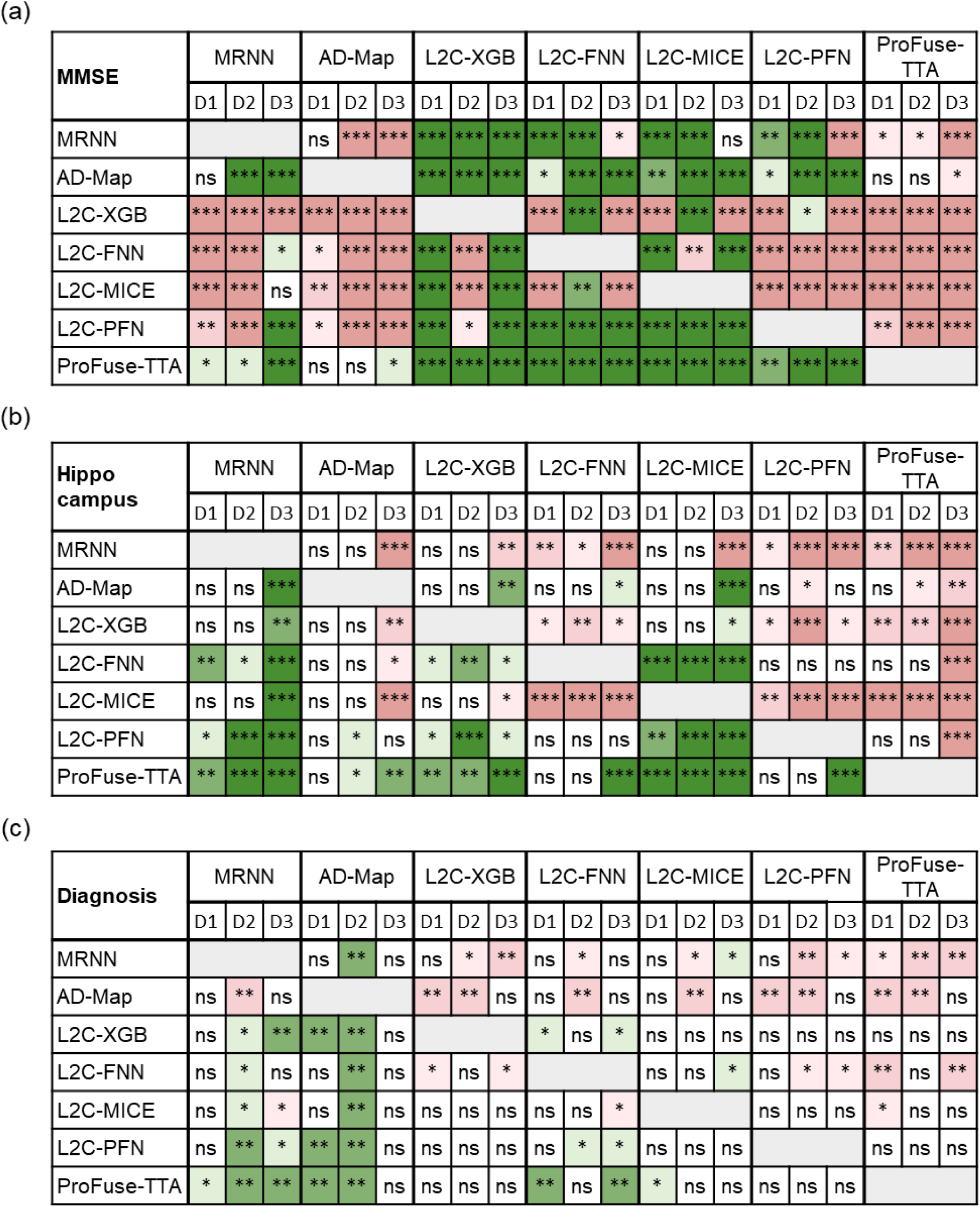
Statistical significance in the prediction performance between all models across the three external datasets. (a) Statistical significance for MMSE prediction. The columns are grouped into seven major columns corresponding to the seven models. Within each major column, the three sub-columns represent results on D1: AIBL, D2: MACC, and D3: OASIS datasets, respectively. Each row shows the statistical difference between one model and all other models. For example, the first row corresponds to the statistical difference between MinimalRNN and the other models – green indicates that MinimalRNN performs better, while red indicates that MinimalRNN performs worse. “*”, “**” and “***” indicate p < 0.05, p < 0.001, and p < 0.00001 respectively, all surviving multiple comparisons correction (FDR q < 0.05). “ns” indicates no statistical significance (p ≥ 0.05) or did not survive FDR correction. (b) Same as (a) but for hippocampus volume prediction. (c) Same as (a) but for clinical diagnosis prediction.

**Table 13.** Cross-cohort prediction performance averaged across 20 trained models (from ADNI). For MMSE and hippocampus volume, lower MAE indicates better performance. For clinical diagnosis, higher mAUC indicates better performance. The best result for each performance metric on each test dataset is bolded.

|  | MMSE MAE ↓ |  |  | Hippocampus MAE (1e-3) ↓ |  |  | Diagnostic mAUC ↑ |  |  |
| --- | --- | --- | --- | --- | --- | --- | --- | --- | --- |
|  | AIBL | MACC | OASIS | AIBL | MACC | OASIS | AIBL | MACC | OASIS |
| MRNN | 1.514 ± 0.103 | 2.765 ± 0.350 | 2.107 ± 0.282 | 0.206 ± 0.013 | 0.217 ± 0.039 | 0.393 ± 0.040 | 0.863 ± 0.020 | 0.898 ± 0.024 | 0.717 ± 0.017 |
| AD-Map | **1.389 ± 0.013** | **2.482 ± 0.018** | 1.606 ± 0.013 | 0.210 ± 0.006 | 0.210 ± 0.003 | 0.280 ± 0.005 | 0.804 ± 0.011 | 0.823 ± 0.007 | 0.740 ± 0.006 |
| L2C-XGB | 2.763 ± 0.835 | 3.933 ± 0.530 | 3.200 ± 0.921 | 0.205 ± 0.006 | 0.218 ± 0.014 | 0.319 ± 0.006 | 0.894 ± 0.008 | 0.958 ± 0.006 | 0.783 ± 0.014 |
| L2C-FNN | 1.809 ± 0.053 | 5.519 ± 0.463 | 1.979 ± 0.053 | 0.188 ± 0.004 | 0.198 ± 0.007 | 0.299 ± 0.005 | 0.865 ± 0.011 | 0.953 ± 0.006 | 0.742 ± 0.018 |
| L2C-MICE | 1.932 ± 0.055 | 5.441 ± 0.392 | 2.051 ± 0.054 | 0.214 ± 0.008 | 0.225 ± 0.012 | 0.337 ± 0.009 | 0.843 ± 0.014 | 0.934 ± 0.013 | 0.710 ± 0.022 |
| L2C-PFN | 1.702 ± 0.030 | 4.261 ± 0.277 | 1.878 ± 0.030 | 0.187 ± 0.003 | **0.190 ± 0.002** | 0.296 ± 0.011 | 0.893 ± 0.005 | **0.963 ± 0.002** | 0.781 ± 0.008 |
| ProFuse-TTA | **1.389 ± 0.029** | 2.617 ± 0.041 | **1.545 ± 0.023** | **0.181 ± 0.004** | 0.194 ± 0.004 | **0.256 ± 0.004** | **0.911 ± 0.004** | 0.958 ± 0.003 | **0.792 ± 0.009** |

**Table 14.** Cross-cohort prediction performance rankings of all models. Each row shows the rank of a model for MMSE MAE, hippocampus volume MAE, and diagnostic mAUC across the AIBL, MACC, and OASIS datasets (1 = best, 7 = worst; ties allowed). Rankings were derived by summing wins (+1) and losses (–1) for each model, as indicated by green (win) and red (loss) in the statistical significance tables (Figure 6). The model with the highest total score receives rank 1 for each metric on each dataset. The "overall" column averages the rankings across cells, so the best possible overall ranking is 1 (first place in every cell). With an overall ranking of 1.1, ProFuse-TTA performed the best.

|  | MMSE MAE |  |  | Hippocampus MAE |  |  | Diagnostic mAUC |  |  | Overall |
| --- | --- | --- | --- | --- | --- | --- | --- | --- | --- | --- |
|  | AIBL | MACC | OASIS | AIBL | MACC | OASIS | AIBL | MACC | OASIS |  |
| MRNN | 3 | 3 | 5 | 5 | 5 | 7 | 4 | 6 | 5 | 4.8 |
| AD-Map | 2 | 1 | 2 | 4 | 4 | 2 | 7 | 7 | 4 | 3.7 |
| L2C-XGB | 7 | 4 | 7 | 5 | 5 | 5 | 2 | 2 | 1 | 4.2 |
| L2C-FNN | 5 | 7 | 4 | 1 | 3 | 4 | 6 | 5 | 5 | 4.4 |
| L2C-MICE | 6 | 6 | 5 | 5 | 5 | 6 | 4 | 2 | 5 | 4.9 |
| L2C-PFN | 4 | 5 | 3 | 1 | 1 | 3 | 3 | 1 | 1 | 2.4 |
| ProFuse-TTA | 1 | 1 | 1 | 1 | 1 | 1 | 1 | 2 | 1 | 1.1 |

In the case of MMSE prediction in the AIBL dataset, ProFuse-TTA was the best, followed by AD-Map. In the MACC dataset, AD-Map and ProFuse-TTA were the best, followed by MinimalRNN. In the OASIS dataset, ProFuse-TTA was the best, followed by AD-Map. Overall, for MMSE prediction, ProFuse-TTA was the best (Table 14).

For hippocampus volume prediction in the AIBL dataset, L2C-FNN, L2C-PFN, and ProFuse-TTA were tied to be the best (Table 14). In the MACC dataset, L2C-PFN and ProFuse-TTA were the best, followed by L2C-FNN. In the OASIS dataset, ProFuse-TTA was the best, followed by AD-Map. Overall, for hippocampal volume prediction, ProFuse-TTA was the best (Table 14).

For clinical diagnosis prediction in the AIBL dataset, ProFuse-TTA was the best, followed by L2C-XGB. In the MACC dataset, L2C-PFN was the best, while L2C-XGB, L2C-MICE and ProFuse-TTA were ranked second. In the OASIS dataset, L2C-XGB, L2C-PFN, and ProFuse-TTA were tied to be the best. Overall, ProFuse-TTA was the best (Table 14).

The “overall” column in Table 14 shows the overall ranking by averaging ranks across all three prediction tasks in the three external datasets. The best possible overall ranking is 1, corresponding to first place in every prediction task in every dataset. ProFuse-TTA achieved an overall ranking of 1.1, ranking first in 8 of the 9 settings, with second place in the remaining setting. The next best model was L2C-PFN with an overall ranking of 2.4.

### 3.4 Further analysis 1: cross-dataset prediction with varying input timepoints

Figures S6 to S8 show the cross-cohort prediction performance of the seven models with varying numbers of observed input timepoints. Numerical values are reported in Tables S14 to S16. Due to the constraints of the datasets, the maximum number of input timepoints for each participant is only 2 for AIBL and 3 for MACC. Therefore, results for AIBL with 3 and 4 timepoints and for MACC with 4 timepoints are marked as "N.A." Figure 7 shows the results of statistical tests comparing ProFuse-TTA with other approaches.

**Figure 7.**
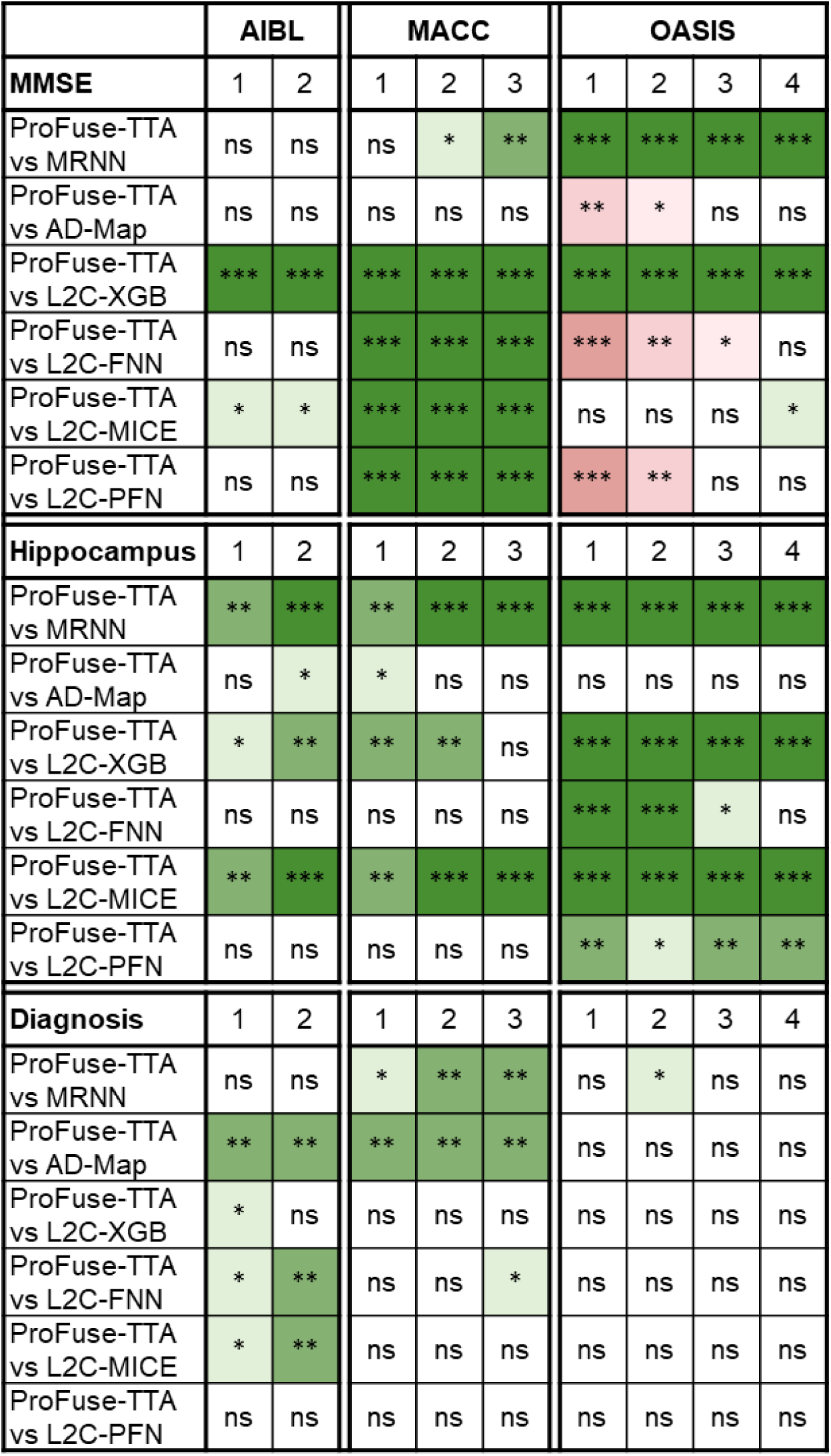
Statistical significance for cross-cohort prediction performance using different numbers of input timepoints. Statistical significance between ProFuse-TTA and other models for cross-cohort MMSE, hippocampus volume, and clinical diagnosis prediction performance (after training on all ADNI timepoints). Green indicates that ProFuse-TTA was statistically better than the model in that column; red indicates that it was statistically worse. “*”, “**”, and “***” indicate p < 0.05, p < 0.001, and p < 0.00001 respectively, all surviving multiple comparisons correction (FDR q < 0.05). "ns" indicates no significance (p ≥ 0.05) or did not survive FDR correction.

As anticipated, prediction performance generally improved with more input timepoints (Figures S6 to S8). Overall, ProFuse-TTA matched or outperformed other approaches across all datasets and timepoint configurations, with the exception of MMSE prediction in the OASIS dataset, where AD-Map, L2C-FNN and L2C-PFN were statistically better than ProFuse-TTA.

### 3.5 Further analysis 2: cross-dataset prediction across varying forecast windows

Figures S9 to S11 show the yearly breakdown of prediction performance from 0 to 6 years into the future (from Figure 5). Numerical values are reported in Tables S17 to S19. Figure 8 shows the results of statistical tests comparing ProFuse-TTA with other approaches across all datasets and forecast horizons. As anticipated, prediction performance for all algorithms declined as the forecast horizon increased. Overall, ProFuse-TTA matched or outperformed other approaches across all forecast horizons and datasets.

**Figure 8.**
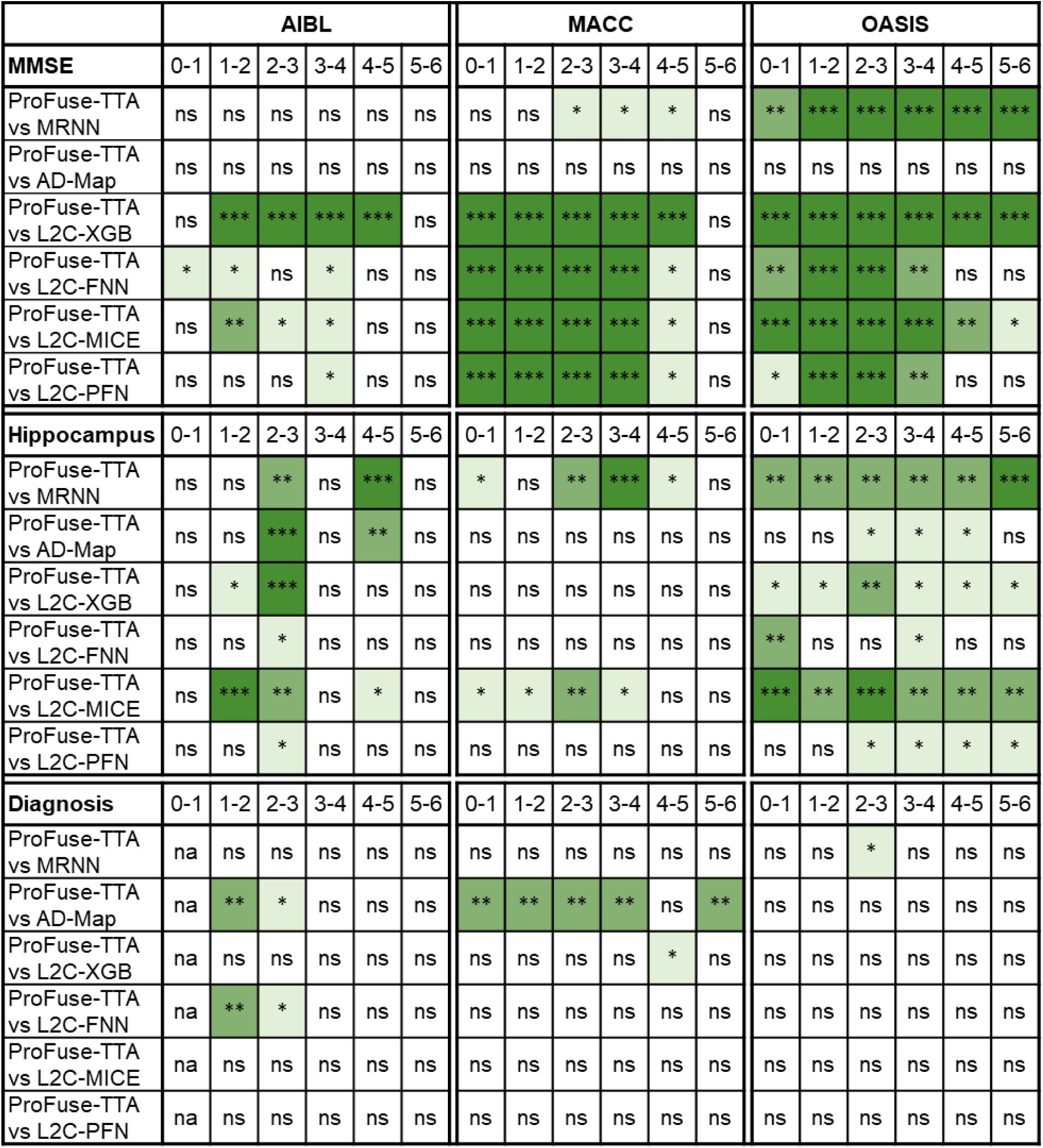
Statistical significance for cross-cohort prediction performance broken down into yearly intervals. Statistical significance between ProFuse-TTA and other models for cross-cohort MMSE, hippocampus volume, and clinical diagnosis prediction (from Figure 5) broken down into yearly intervals up to 6 years into the future. Green indicates that ProFuse-TTA was statistically better than other approaches compared, while red indicates that it was statistically worse. “*”, “**” and “***” indicate p < 0.05, p < 0.001, and p < 0.00001 respectively, all surviving multiple comparisons correction (FDR q < 0.05). “ns” indicates no significance (p ≥ 0.05) or did not survive FDR correction. Due to dataset constraints, AIBL only had one diagnostic class in year 0-1, making mAUC undefined in this case. Therefore, results for AIBL at year 0-1 is marked as “na”.

### 3.6 History-aware loss-gating outperforms global test-time adaptation

Figure 9 shows the performance change of three test-time adaptation strategies (base-light, base-heavy, and our proposed history-aware loss-gating approach) relative to the non-adapted base ProFuse model across the three external datasets. Pairwise statistical comparisons are reported in Figure S12.

**Figure 9.**
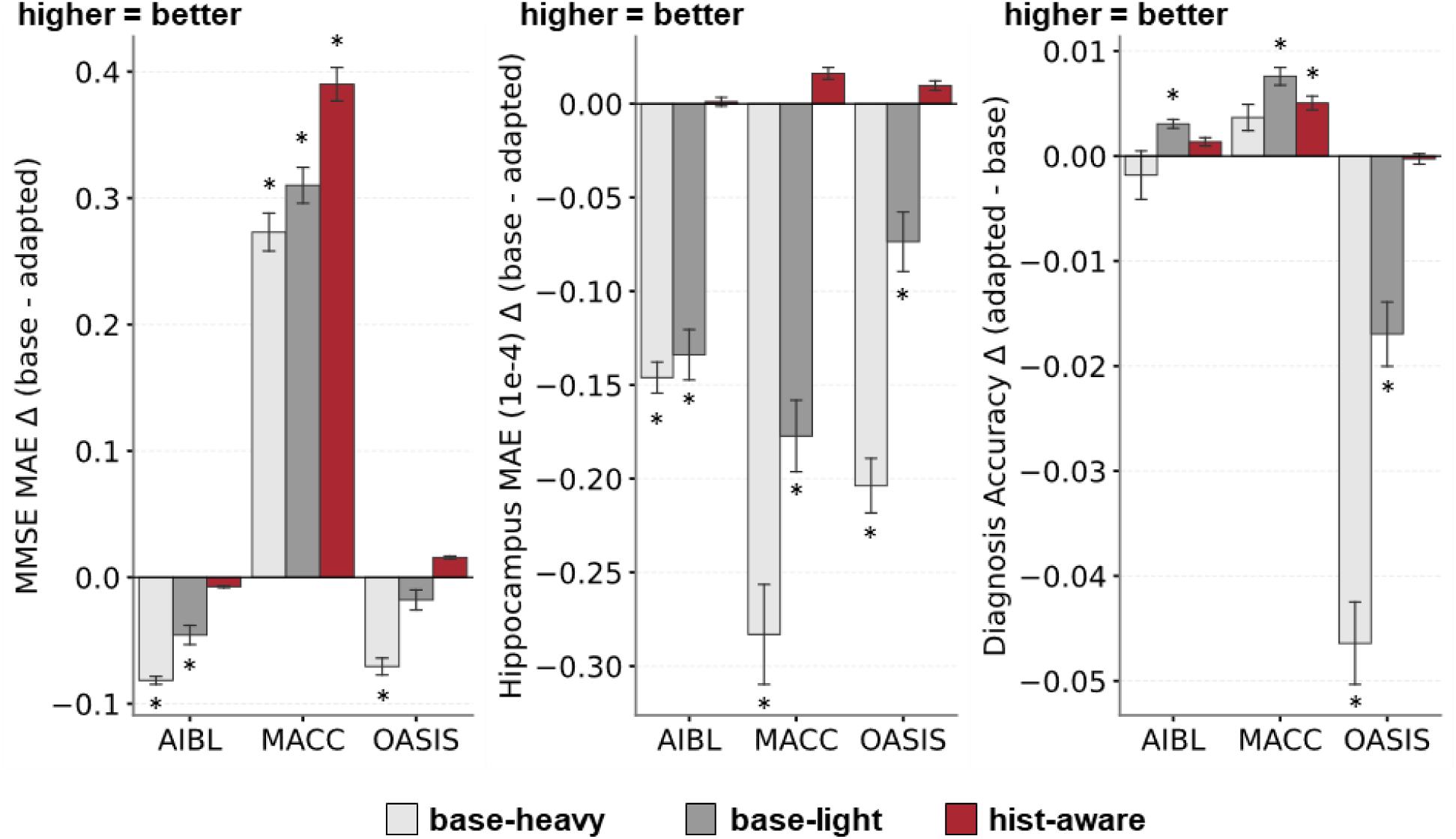
Cross-cohort performance gains of test-time adaptation strategies relative to the non-adapted base model across three external test datasets. Bar plots show the mean performance change across 20 ADNI-trained models for three adaptation strategies: heavily tuned full-model adaptation (base-heavy), lightly tuned full-model adaptation (base-light), and history-aware loss-gating (hist-aware). The performance change is defined as (base − adapted) for MMSE and hippocampal volume MAE, and (adapted − base) for diagnostic mAUC, so that higher values indicate improvement for all three tasks. The x-axis shows the external evaluation dataset. Asterisks denote statistically significant differences from the non-adapted base model (full pairwise comparisons are found in Figure S12). Error bars indicate ±1 standard error.

All three adaptation strategies significantly improved MMSE prediction in the MACC dataset, which has the largest distribution shift from ADNI (Table 2). However, base-light and base-heavy significantly degraded MMSE performance on AIBL and OASIS, while history-aware loss-gating produced only minimal degradation on AIBL and a numerical improvement on OASIS.

For hippocampus volume prediction, both global tuning variants (base-light and base-heavy) degraded performance on all three external datasets, with the most severe degradation on MACC. In contrast, our proposed history-aware loss-gating matched the base model on AIBL and produced numerical improvements on MACC and OASIS.

For clinical diagnosis prediction, base-light significantly improved performance on AIBL and MACC but significantly degraded performance on OASIS. Base-heavy was numerically worse on AIBL, numerically better on MACC, and significantly worse on OASIS. History-aware loss-gating produced numerical improvement on AIBL, statistically significant improvement on MACC, and matched the base model on OASIS.

Overall, our proposed history-aware loss-gating matched or significantly improved on the base model across all external datasets and tasks. By contrast, base-light and base-heavy each yielded large improvements in some scenarios but severe degradation in others.

## 4 Discussion

In this study, we proposed ProFuse-TTA, and compared it with MinimalRNN, AD-Map, and four L2C-based variants (L2C-XGB, L2C-FNN, L2C-MICE, L2C-PFN) on the ADNI dataset and three external cohorts (AIBL, MACC, OASIS). Within ADNI, L2C-PFN ranked first across all three tasks, with ProFuse-TTA closely behind (Table 11), highlighting the strong in-domain capability of TabPFN on tabular data. However, in the controlled within-ADNI modality ablation, ProFuse-TTA ranked first in 14 of 15 scenarios (Table 12), with the smallest performance drop in every scenario where the key modality for a task was removed. Across the three external cohorts, ProFuse-TTA ranked first in 8 of 9 settings and second in the remaining one (Table 14), and maintained this advantage across varying numbers of observed input timepoints (Figure 7) and across prediction horizons from 0 to 6 years (Figure 8).

### 4.1 Algorithmic considerations

Existing strategies for handling sporadic missingness do not necessarily extend to systemic missingness. This was most evident for the two impute-then-predict baselines. L2C-FNN, which was the best performing method in our previous study (Zhang et al., 2025), dropped substantially in ranking once systemic missingness was introduced. L2C-MICE performed even worse despite modeling multivariate dependencies during imputation, consistently underperforming L2C-FNN in both modality ablation (Table 12) and cross-cohort evaluation (Table 14). A key limitation of the impute-then-predict paradigm is that imputation is performed independently of the prediction task, so the resulting values are not aligned with predictive objectives (Wells et al., 2013; Che et al., 2018). Simple methods such as median imputation introduce consistent bias that downstream models can partially accommodate, while MICE produces more complex imputations that introduce structured errors. The issue is compounded under systemic missingness, where the missing-at-random assumption underlying MICE is often violated (White et al., 2011), particularly when high-dimensional modality groups like MRI or cognition are completely absent.

MinimalRNN integrates imputation and prediction within a unified model-filling framework, but its performance under systemic missingness remains limited. The ablation experiments show that MinimalRNN is highly sensitive to the removal of the key modality for each prediction task (e.g., COG for MMSE, MRI for hippocampal volume). We hypothesize that this stems from its autoregressive inductive bias: although the model is trained to predict all biomarkers jointly, the objective favors within-modality extrapolation over learning strong cross-modal dependencies, limiting its ability to infer a missing modality from the available ones.

L2C-XGB showed one of the steepest performance declines under systemic missingness despite ranking on par with ProFuse-TTA within ADNI (Table 11). This fragility reflects the nature of XGBoost’s native missingness handling, which relies on fixed default split directions learned only from sporadic missingness during training. When an entire modality is removed at test time, all splits involving its features fall back to the default splits, which were not meant for handling systemic missingness. There is no mechanism to reweight or re-route decisions based on the remaining modalities. The COG ablation illustrates this: MMSE prediction error increases by 2.2, nearly double that of the next-worst model (Table S11).

L2C-PFN achieves exceptional within-ADNI performance, substantially outperforming all other methods. However, its performance drops noticeably under systemic missingness, where it is consistently outperformed by ProFuse-TTA, especially when key modalities are removed (Tables 12 and 14). This likely reflects a mismatch between the missingness patterns at evaluation and those seen during pretraining. TabPFN is pretrained on synthetic datasets with sporadic missingness under a missing-completely-at-random assumption (Hollmann et al., 2025), whereas our experiments introduce structured, modality-level missingness that falls outside its learned prior. Adapting the pretraining process to incorporate more realistic missingness patterns may help mitigate this limitation.

Unlike the L2C variants that decouple feature extraction from prediction, ProFuse-TTA learns end-to-end from raw longitudinal data and handles missingness through architectural design rather than imputation. In Stage 1, each biomarker is encoded using only its observed timepoints, naturally bypassing sporadic missingness. In Stage 2, a structured embedding design encodes both feature identity and missingness, allowing the attention mechanism to dynamically integrate information across available biomarkers while explicitly distinguishing absent ones. Combined with simulated modality dropout during training, this enables ProFuse-TTA to learn robust strategies for structured, modality-level missingness that are not captured by MinimalRNN’s within-modality extrapolation, L2C-XGB’s static split routing, or TabPFN’s pretrained prior.

### 4.2 Distribution shifts and test-time adaptation

Cross-dataset evaluation also introduces substantial distribution shifts between training and test cohorts. The average baseline MMSE score was 27.4 in ADNI, 28.0 in AIBL, 28.3 in OASIS, but only 21.6 in MACC (Table 2), and prediction error on MACC was correspondingly higher across all methods (Table 13). The proposed history-aware loss-gating significantly improved MMSE prediction on MACC, demonstrating that test-time adaptation can be effective for severe distribution shifts (Figure 9). However, the two global tuning variants (base-light and base-heavy) also improved MACC but at the expense of degraded performance for other datasets and target variables.

In contrast, our adaptation strategy matched or improved on the base model across all datasets and tasks. We attribute this robustness to four design choices. First, lightweight task-specific adapters preserve the population-level patterns learned by the base model. Second, anchoring the regression adapter to the observed historical trajectory addresses an identifiability issue: identical prediction errors can correspond to very different underlying trajectories, and the historical anchor lets the model distinguish such cases. Third, per-task optimization avoids cross-task gradient interference. Fourth, loss-gating automatically reduces adaptation strength when the base model is already well-calibrated, which is supported by the empirical observation that base-heavy consistently underperforms base-light.

### 4.3 Limitations

A limitation of the current study is that ProFuse-TTA is trained on a single dataset (ADNI) and a feature set constrained by what was available at training time. Training on a more diverse collection of large-scale cohorts such as the National Alzheimer’s Coordinating Center (NACC) and European Prevention of Alzheimer’s Dementia (EPAD) datasets (Besser et al., 2018; Lorenzini et al., 2022) would expose the model to broader heterogeneity in population characteristics, acquisition protocols, and clinical practices, potentially yielding representations that are inherently more robust to cross-dataset variation (Dou et al., 2019; Q. Liu et al., 2020; P. Chen et al., 2024). Incorporating emerging biomarkers such as plasma-based assays and retinal imaging (Cheung et al., 2021; Chong et al., 2021; Ashton et al., 2024; An et al., 2026) would also extend clinical applicability and allow evaluation of their predictive value relative to established markers. The proposed architecture is well-suited for such extensions.

A further consideration concerns the conditions under which test-time adaptation can be applied. The proposed strategy operates in a fully unsupervised manner by constructing one-step-ahead prediction pairs from the observed input trajectory, and therefore requires at least two observed timepoints, with the target variable observed in the historical trajectory for the history-aware adjustment to engage. When these conditions are not met, the adaptation module is bypassed, and the base model prediction is used directly. While this design ensures stable performance and avoids degradation in sparse-data scenarios, it implies that the benefits of adaptation may be limited when historical observations are extremely scarce. Extending the framework to handle such cases is an important direction for future work.

## 5 Conclusion

We introduced ProFuse-TTA, a two-stage hierarchical transformer that handles systemic missingness through architectural design rather than imputation, paired with a history-aware test-time adaptation module that calibrates predictions to individual trajectories. Across controlled modality ablations and three external cohorts spanning 2,316 participants and 13,205 timepoints, ProFuse-TTA outperformed impute-then-predict baselines, autoregressive recurrent models, tree-based learners, and a tabular foundation model. These results position ProFuse-TTA as a practical and extensible framework for real-world clinical deployment, where incomplete data and heterogeneous cohorts are the rule rather than the exception.

## 6 Acknowledgment

Our research is supported by the NUS Yong Loo Lin School of Medicine (NUHSRO/2020/124/TMR/LOA), the Singapore National Medical Research Council (NMRC) LCG (OFLCG19May-0035), NMRC CTG-IIT (CTGIIT23jan-0001), NMRC OF-IRG (OFIRG24jan-0006; OFIRG24jul-0049), NMRC STaR (STaR20nov-0003), Singapore Ministry of Health (MOH) Centre Grant (CG21APR1009), the United States National Institutes of Health (R01MH133334 & 2R01MH120080) and the Singapore National Research Foundation (NRF) Investigatorship (NRFI10-2024-0014). Any opinions, findings and conclusions or recommendations expressed in this material are those of the authors and do not reflect the views of the funders.

Data collection and sharing for the Alzheimer’s Disease Neuroimaging Initiative (ADNI) is funded by the National Institute on Aging (National Institutes of Health Grant U19 AG024904). The grantee organization is the Northern California Institute for Research and Education. In the past, ADNI has also received funding from the National Institute of Biomedical Imaging and Bioengineering, the Canadian Institutes of Health Research, and private sector contributions through the Foundation for the National Institutes of Health (FNIH) including generous contributions from the following: AbbVie, Alzheimer’s Association; Alzheimer’s Drug Discovery Foundation; Araclon Biotech; BioClinica, Inc.; Biogen; Bristol-Myers Squibb Company; CereSpir, Inc.; Cogstate; Eisai Inc.; Elan Pharmaceuticals, Inc.; Eli Lilly and Company; EuroImmun; F. Hoffmann-La Roche Ltd and its affiliated company Genentech, Inc.; Fujirebio; GE Healthcare; IXICO Ltd.; Janssen Alzheimer Immunotherapy Research & Development, LLC.; Johnson & Johnson Pharmaceutical Research &Development LLC.; Lumosity; Lundbeck; Merck & Co., Inc.; Meso Scale Diagnostics, LLC.; NeuroRx Research; Neurotrack Technologies; Novartis Pharmaceuticals Corporation; Pfizer Inc.; Piramal Imaging; Servier; Takeda Pharmaceutical Company; and Transition Therapeutics.

We thank all the participants and their families who took part in the AIBL Study, as well as the clinicians who may have referred participants. The AIBL Study (https://aibl.org.au/) is a consortium comprising Austin Health, CSIRO, Edith Cowan University, the Florey Institute for Neuroscience and Mental Health (University of Melbourne), and the National Ageing Research Institute. Numerous commercial interactions have supported data collection and analyses. In-kind support has been provided by the consortium institutions, as well as Alzheimer’s Research Australia, Sir Charles Gairdner Hospital, Hollywood Private Hospital, and St Vincent’s Hospital. The study has also received financial support at various stages from the Alzheimer’s Drug Discovery Foundation, the Victorian Government’s Operational Infrastructure Support program, Alzheimer’s Research Australia, and the National Health and Medical Research Council (NHMRC).

Data were provided in part by OASIS-3: Longitudinal Multimodal Neuroimaging: Principal Investigators: T. Benzinger, D. Marcus, J. Morris; NIH P30 AG066444, P50 AG00561, P30 NS09857781, P01 AG026276, P01 AG003991, R01 AG043434, UL1 TR000448, R01 EB009352. AV-45 doses were provided by Avid Radiopharmaceuticals, a wholly owned subsidiary of Eli Lilly.

## 7 Data availability statement

The ADNI and the AIBL datasets can be accessed via the Image & Data Archive (https://ida.loni.usc.edu/). The MACC dataset can be obtained via a data-transfer agreement with the MACC (http://www.macc.sg/). The OASIS dataset can be requested from (https://www.oasis-brains.org/).

## 8 Code availability statement

Code for all five models can be found here (GITHUB_LINK). Two co-authors (H.L. and F.T.) reviewed the code before merging it into the GitHub repository to reduce the chance of coding errors.

## Footnotes

1 https://github.com/ThomasYeoLab/CBIG/tree/master/stable_projects/predict_phenotypes/Zhang2025_L2CFNN

2 https://gitlab.com/icm-institute/aramislab/leaspy

3 https://anaconda.org/conda-forge/miceforest

4 https://github.com/PriorLabs/TabPFN

5 https://huggingface.co/docs/transformers/model_doc/roformer

6 https://huggingface.co/docs/transformers/model_doc/bert

