## Supplemental materials for "Robust Longitudinal Dementia Prediction under Systemic Missingness via Hierarchical Fusion and Test-Time Adaptation"

### Supplementary Materials

Table S1. Scanner information for 9671 scans in ADNI dataset.

| Vendor | Scanner Model | Field Strength | Number of Scans |
| --- | --- | --- | --- |
| GE | Discovery MR750 | 3.0T | 886 |
|  | Discovery MR750w | 3.0T | 123 |
|  | Genesis Signa | 1.5T | 248 |
|  |  | 3.0T | 6 |
|  | Signa Excite | 1.5T | 892 |
|  |  | 3.0T | 6 |
|  | Signa HDx | 1.5T | 463 |
|  |  | 3.0T | 36 |
|  | Signa HDxt | 1.5T | 208 |
|  |  | 3.0T | 419 |
|  | Signa Premier | 3.0T | 34 |
|  | Signa UHP | 3.0T | 1 |
| Philips | Achieva dStream | 3.0T | 88 |
|  | Ingenia | 3.0T | 180 |
|  | Ingenia Elition X | 3.0T | 8 |
|  | Achieva | 1.5T | 73 |
|  |  | 3.0T | 541 |
|  | Gemini | 3.0T | 32 |
|  | Gyroscan Intera | 1.5T | 12 |
|  | Gyroscan NT | 1.5T | 2 |
|  | Ingenuity | 3.0T | 18 |
|  | Intera | 1.5T | 333 |
|  |  | 3.0T | 217 |
|  | Intera Achieva | 1.5T | 6 |
| SIEMENS | Allegra | 3.0T | 32 |
|  | Avanto | 1.5T | 391 |
|  | Biograph_mMR | 3.0T | 19 |
|  | Espree | 1.5T | 24 |
|  | Numaris4 | 1.5T | 2 |
|  | Prisma | 3.0T | 191 |
|  | Prisma_fit/Magnetom Prisma_fit | 3.0T | 466 |
|  | Skyra | 3.0T | 406 |
|  | Skyra_fit | 3.0T | 17 |
|  | Sonata | 1.5T | 379 |
|  | SonataVision | 1.5T | 31 |
|  | Symphony/SymphonyTim | 1.5T | 671 |
|  | Trio/TrioTim | 3.0T | 1489 |
|  | Verio | 3.0T | 718 |
|  | Skyra\|DicomCleaner | 3.0T | 3 |

Table S2. Scanner information for 942 scans in AIBL dataset.

| Vendor | Scanner Model | Field Strength | Number of Scans |
| --- | --- | --- | --- |
| SIEMENS | Avanto | 1.5T | 245 |
|  | Trio/TrioTim | 3.0T | 624 |
|  | Verio | 3.0T | 73 |

Table S3. Scanner information for 1456 scans in MACC dataset.

| Vendor | Scanner Model | Field Strength | Number of Scans |
| --- | --- | --- | --- |
| SIEMENS | Prisma | 3.0T | 86 |
|  | Trio/TrioTim | 3.0T | 1370 |

Table S4. Scanner information for 2519 scans in OASIS dataset.

| Vendor | Scanner Model | Field Strength | Number of Scans |
| --- | --- | --- | --- |
| SIEMENS | Avanto | 1.5T | 2 |
|  | Biograph_mMR | 3.0T | 812 |
|  | Magnetom Vida | 3.0T | 305 |
|  | Prisma_fit/Magnetom Prisma_fit | 3.0T | 1 |
|  | Sonata | 1.5T | 40 |
|  | Trio/TrioTim | 3.0T | 1359 |

Table S5. Six regional brain volumes used in our study

(Hippocampus, Entorhinal, Fusiform, MidTemp, Ventricles, WholeBrain), computed by summing the volumes of various FreeSurfer regions of interest. Note that the 7th anatomical volume (intracranial volume) was directly provided by FreeSurfer.

| Regional volumetric feature | Brain regions used to compute regional volumes |
| --- | --- |
| Hippocampus | Left-Hippocampus, Right-Hippocampus |
| Entorhinal | lh_entorhinal_volume, rh_entorhinal_volume |
| Fusiform | lh_fusiform_volume, rh_fusiform_volume |
| MidTemp | lh_middletemporal_volume, rh_middletemporal_volume |
| Ventricles | Left-Inf-Lat-Vent, Left-Lateral-Ventricle,  Right-Inf-Lat-Vent, Right-Lateral-Ventricle |
| WholeBrain | WM-hypointensities, Left-Cerebellum-Cortex,  Left-Cerebellum-White-Matter, Left-Thalamus-Proper,  Left-Caudate, Left-Putamen,  Left-Pallidum, Left-Hippocampus,  Left-Amygdala, Left-Accumbens-area,  Left-VentralDC, Right-Cerebellum-Cortex,  Right-Cerebellum-White-Matter, Right-Thalamus-Proper, Right-Caudate, Right-Putamen,  Right-Pallidum, Right-Hippocampus,  Right-Amygdala, Right-Accumbens-area,  Right-VentralDC, lhCortexVol,  rhCortexVol, lhCerebralWhiteMatterVol, rhCerebralWhiteMatterVol |

Table S6. Complete set of L2C features and their corresponding original features.

| Original feature | L2C features |
| --- | --- |
| Demographic & Static Features | |
| Number of APOE-ε4 alleles (categorical: 0, 1, 2) | APOE4 |
| Sex (categorical: female, male) | is_male |
| Years of education received (numeric, ordinal) | educ |
| Marital status (categorical: married, not married) | marital_status |
| Age at target timepoint (numeric, continuous) | current_age |
| Number of months since baseline visit (numeric, continuous) | month_since_baseline |
| Longitudinal Features | |
| Clinical diagnosis (categorical: CN, MCI, DEM) | mr_dx, time_since_mr_dx, best_dx, time_since_best_dx, worst_dx, time_since_worst_dx, milder, time_since_milder |
| MMSE score (numeric, ordinal: 0-30) | mr_MMSE, time_since_mr_MMSE, mr_change_MMSE, low_MMSE, time_since_low_MMSE, high_MMSE, time_since_high_MMSE |
| MoCA score (numeric, ordinal: 0-30) | mr_MoCA, time_since_mr_MoCA, mr_change_MoCA, low_MoCA, time_since_low_MoCA, high_MoCA, time_since_high_MoCA |
| FAQ score (numeric, ordinal: 0-30) | mr_FAQ, time_since_mr_FAQ, mr_change_FAQ, low_FAQ, time_since_low_FAQ, high_FAQ, time_since_high_FAQ |
| CDRSB score (numeric, ordinal: 0-18) | mr_CDRSB, time_since_mr_CDRSB, mr_change_CDRSB, low_CDRSB, time_since_low_CDRSB, high_CDRSB, time_since_high_CDRSB |
| ADAS-11 score (numeric, ordinal: 0-70) | mr_ADAS11, time_since_mr_ADAS11, mr_change_ADAS11, low_ADAS11, time_since_low_ADAS11, high_ADAS11, time_since_high_ADAS11 |
| ADAS-13 score (numeric, ordinal: 0-85) | mr_ADAS13, time_since_mr_ADAS13, mr_change_ADAS13, low_ADAS13, time_since_low_ADAS13, high_ADAS13, time_since_high_ADAS13 |
| RAVLT immediate score (numeric, ordinal: 0-75) | mr_RAVLT_immediate, time_since_mr_RAVLT_immediate, mr_change_RAVLT_immediate, low_RAVLT_immediate, time_since_low_RAVLT_immediate, high_RAVLT_immediate, time_since_high_RAVLT_immediate |
| RAVLT learning score (numeric, ordinal: 0-15) | mr_RAVLT_learning, time_since_mr_RAVLT_learning, mr_change_RAVLT_learning, low_RAVLT_learning, time_since_low_RAVLT_learning, high_RAVLT_learning, time_since_high_RAVLT_learning |
| RAVLT forgetting score (numeric, ordinal: 0-15) | mr_RAVLT_forgetting, time_since_mr_RAVLT_forgetting, mr_change_RAVLT_forgetting, low_RAVLT_forgetting, time_since_low_RAVLT_forgetting, high_RAVLT_forgetting, time_since_high_RAVL_forgetting |
| RAVLT percent forgetting score (numeric, continuous: 0-100) | mr_RAVLT_perc_forgetting, time_since_mr_RAVLT_perc_forgetting, mr_change_RAVLT_perc_forgetting, low_RAVLT_perc_forgetting, time_since_low_RAVLT_perc_forgetting, high_RAVLT_perc_forgetting, time_since_high_RAVL_perc_forgetting |
| Entorhinal volume (numeric, continuous) | mr_Entorhinal, time_since_mr_Entorhinal, mr_change_Entorhinal, low_Entorhinal, time_since_low_Entorhinal, high_Entorhinal, time_since_high_Entorhinal |
| Ventricle volume (numeric, continuous) | mr_Ventricles, time_since_mr_Ventricles, mr_change_Ventricles, low_Ventricles, time_since_low_Ventricles, high_Ventricles, time_since_high_Ventricles |
| Fusiform volume (numeric, continuous) | mr_Fusiform, time_since_mr_Fusiform, mr_change_Fusiform, low_Fusiform, time_since_low_Fusiform, high_Fusiform, time_since_high_Fusiform |
| WholeBrain volume (numeric, continuous) | mr_WholeBrain, time_since_mr_WholeBrain, mr_change_WholeBrain, low_WholeBrain, time_since_low_WholeBrain, high_WholeBrain, time_since_high_WholeBrain |
| Hippocampus volume (numeric, continuous) | mr_Hippocampus, time_since_mr_Hippocampus, mr_change_Hippocampus, low_Hippocampus, time_since_low_Hippocampus, high_Hippocampus, time_since_high_Hippocampus |
| Middle temporal volume (numeric, continuous) | mr_MidTemp, time_since_mr_MidTemp, mr_change_MidTemp, low_MidTemp, time_since_low_MidTemp, high_MidTemp, time_since_high_MidTemp |
| Intracranial volume (numeric, continuous) | mr_ICV, time_since_mr_ICV, mr_change_ICV, low_ICV, time_since_low_ICV, high_ICV, time_since_high_ICV |
| AV45 SUVR (numeric, continuous) | mr_AV45, time_since_mr_AV45, mr_change_AV45, low_AV45, time_since_low_AV45, high_AV45, time_since_high_AV45 |
| FDG SUVR (numeric, continuous) | mr_FDG, time_since_mr_FDG, mr_change_FDG, low_FDG, time_since_low_FDG, high_FDG, time_since_high_FDG |
| ABETA concentration (numeric, continuous) | mr_ABETA, time_since_mr_ABETA, mr_change_ABETA, low_ABETA, time_since_low_ABETA, high_ABETA, time_since_high_ABETA |
| TAU concentration (numeric, continuous) | mr_TAU, time_since_mr_TAU, mr_change_TAU, low_TAU, time_since_low_TAU, high_TAU, time_since_high_TAU |
| PTAU concentration (numeric, continuous) | mr_PTAU, time_since_mr_PTAU, mr_change_PTAU, low_PTAU, time_since_low_PTAU, high_PTAU, time_since_high_PTAU |

Table S7. Preprocessed input vector dimensionality (for L2C-FNN and L2C-MICE).

For discrete features, the calculation is performed after one-hot encoding, including an additional class for missing data or unknown.

| Category | L2C features | # features |
| --- | --- | --- |
| Discrete features (dimensions are after one-hot encoding, including “unknown” class) | apoe | 4 |
|  | is_male | 3 |
|  | marital_status | 3 |
|  | mr_dx | 4 |
|  | best_dx | 4 |
|  | worst_dx | 4 |
|  | milder | 3 |
| MRI features | mr_Entorhinal, time_since_mr_Entorhinal, mr_change_Entorhinal, low_Entorhinal, time_since_low_Entorhinal, high_Entorhinal, time_since_high_Entorhinal,  mr_Ventricles, time_since_mr_Ventricles, mr_change_Ventricles, low_Ventricles, time_since_low_Ventricles, high_Ventricles, time_since_high_Ventricles,  mr_Fusiform, time_since_mr_Fusiform, mr_change_Fusiform, low_Fusiform, time_since_low_Fusiform, high_Fusiform, time_since_high_Fusiform,  mr_WholeBrain, time_since_mr_WholeBrain, mr_change_WholeBrain, low_WholeBrain, time_since_low_WholeBrain, high_WholeBrain, time_since_high_WholeBrain,  mr_Hippocampus, time_since_mr_Hippocampus, mr_change_Hippocampus, low_Hippocampus, time_since_low_Hippocampus, high_Hippocampus, time_since_high_Hippocampus,  mr_MidTemp, time_since_mr_MidTemp, mr_change_MidTemp, low_MidTemp, time_since_low_MidTemp, high_MidTemp, time_since_high_MidTemp,  mr_ICV, time_since_mr_ICV, mr_change_ICV, low_ICV, time_since_low_ICV, high_ICV, time_since_high_ICV | 49 |
| Cognitive features | mr_MMSE, time_since_mr_MMSE, mr_change_MMSE, low_MMSE, time_since_low_MMSE, high_MMSE, time_since_high_MMSE,  mr_MoCA, time_since_mr_MoCA, low_MoCA, time_since_low_MoCA, high_MoCA, time_since_high_MoCA,  mr_FAQ, time_since_mr_FAQ, mr_change_FAQ, low_FAQ, time_since_low_FAQ, high_FAQ, time_since_high_FAQ,  mr_CDRSB, time_since_mr_CDRSB, mr_change_CDRSB, low_CDRSB, time_since_low_CDRSB, high_CDRSB, time_since_high_CDRSB,  mr_ADAS11, time_since_mr_ADAS11, mr_change_ADAS11, low_ADAS11, time_since_low_ADAS11, high_ADAS11, time_since_high_ADAS11,  mr_ADAS13, time_since_mr_ADAS13, mr_change_ADAS13, low_ADAS13, time_since_low_ADAS13, high_ADAS13, time_since_high_ADAS13,  mr_RAVLT_immediate, time_since_mr_RAVLT_immediate, mr_change_RAVLT_immediate, low_RAVLT_immediate, time_since_low_RAVLT_immediate, high_RAVLT_immediate, time_since_high_RAVLT_immediate,  mr_RAVLT_learning, time_since_mr_RAVLT_learning, mr_change_RAVLT_learning, low_RAVLT_learning, time_since_low_RAVLT_learning, high_RAVLT_learning, time_since_high_RAVL_learning,  mr_RAVLT_forgetting, time_since_mr_RAVLT_forgetting, mr_change_RAVLT_forgetting, low_RAVLT_forgetting, time_since_low_RAVLT_forgetting, high_RAVLT_forgetting, time_since_high_RAVL_forgetting,  mr_RAVLT_perc_forgetting, time_since_mr_RAVLT_perc_forgetting, mr_change_RAVLT_perc_forgetting, low_RAVLT_perc_forgetting, time_since_low_RAVLT_perc_forgetting, high_RAVLT_perc_forgetting, time_since_high_RAVL_perc_forgetting | 69 |
| PET features | mr_AV45, time_since_mr_AV45, low_AV45, time_since_low_AV45, high_AV45, time_since_high_AV45  mr_FDG, time_since_mr_FDG, low_FDG, time_since_low_FDG, high_FDG, time_since_high_FDG | 12 |
| CSF features | mr_ABETA, time_since_mr_ABETA, low_ABETA, time_since_low_ABETA, high_ABETA, time_since_high_ABETA  mr_TAU, time_since_mr_TAU, low_TAU, time_since_low_TAU, high_TAU, time_since_high_TAU  mr_PTAU, time_since_mr_PTAU, low_PTAU, time_since_low_PTAU, high_PTAU, time_since_high_PTAU | 18 |
| Diagnostic features | time_since_mr_dx, time_since_best_dx, time_since_worst_dx | 3 |
| Demographic and time-dependent features | baseline education level, current_age, month_since_baseline | 3 |

Table S8. Within-cohort (ADNI) MMSE prediction performance under different modality ablation scenarios.

Results were averaged across 20 trained models (obtained from training with all data in ADNI). Lower MAE indicates better performance. The best result for each ablation scenario is bolded. Due to model design, AD-Map does not utilize diagnostic input features and therefore has no results in the “Ablate DX” condition, which is marked as “N.A.”

| MMSE MAE ↓ | No ablation | Ablate DX | Ablate COG | Ablate MRI | Ablate CSF | Ablate PET |
| --- | --- | --- | --- | --- | --- | --- |
| MRNN | 1.561 ± 0.270 | 1.577 ± 0.261 | 2.474 ± 0.421 | 1.566 ± 0.275 | 1.577 ± 0.280 | 1.582 ± 0.262 |
| AD-Map | 1.701 ± 0.251 | N.A. | 2.329 ± 0.353 | 1.717 ± 0.251 | 1.705 ± 0.265 | 1.704 ± 0.249 |
| L2C-XGB | 1.540 ± 0.220 | 1.659 ± 0.237 | 3.758 ± 0.982 | 1.569 ± 0.267 | 1.533 ± 0.234 | 1.555 ± 0.229 |
| L2C-FNN | 1.511 ± 0.239 | 1.551 ± 0.277 | 2.460 ± 0.453 | 1.577 ± 0.264 | 1.499 ± 0.232 | 1.515 ± 0.218 |
| L2C-MICE | 1.492 ± 0.238 | 1.508 ± 0.247 | 2.632 ± 0.480 | 1.750 ± 0.353 | 1.522 ± 0.198 | 1.533 ± 0.220 |
| L2C-PFN | **1.425 ± 0.195** | **1.441 ± 0.207** | 2.348 ± 0.455 | 1.465 ± 0.228 | **1.446 ± 0.215** | **1.455 ± 0.202** |
| ProTuse-TTA | 1.462 ± 0.230 | 1.467 ± 0.226 | **1.853 ± 0.306** | **1.441 ± 0.221** | 1.464 ± 0.237 | 1.483 ± 0.251 |

Table S9. Within-cohort (ADNI) hippocampus volume prediction performance under different modality ablation scenarios.

Results were averaged across 20 trained models (obtained from training with all data in ADNI). Lower MAE indicates better performance. The best result for each ablation scenario is bolded. Due to model design, AD-Map does not utilize diagnostic input features and therefore has no results in the “Ablate DX” condition, which is marked as “N.A.”

| Hippocampus MAE (1e-3) ↓ | No ablation | Ablate DX | Ablate COG | Ablate MRI | Ablate CSF | Ablate PET |
| --- | --- | --- | --- | --- | --- | --- |
| MRNN | 0.164 ± 0.022 | 0.165 ± 0.023 | 0.162 ± 0.025 | 0.613 ± 0.073 | 0.166 ± 0.023 | 0.161 ± 0.019 |
| AD-Map | 0.161 ± 0.023 | N.A. | 0.165 ± 0.024 | 0.478 ± 0.080 | 0.161 ± 0.023 | 0.161 ± 0.023 |
| L2C-XGB | 0.151 ± 0.020 | 0.151 ± 0.020 | 0.174 ± 0.018 | 0.569 ± 0.072 | 0.154 ± 0.020 | 0.155 ± 0.019 |
| L2C-FNN | 0.154 ± 0.026 | 0.156 ± 0.026 | 0.163 ± 0.022 | 0.553 ± 0.060 | 0.157 ± 0.025 | 0.154 ± 0.026 |
| L2C-MICE | 0.153 ± 0.022 | 0.156 ± 0.022 | 0.173 ± 0.018 | 0.655 ± 0.056 | 0.166 ± 0.025 | 0.156 ± 0.025 |
| L2C-PFN | **0.142 ± 0.019** | **0.142 ± 0.019** | **0.146 ± 0.019** | 0.501 ± 0.068 | 0.151 ± 0.022 | **0.143 ± 0.019** |
| ProTuse-TTA | 0.147 ± 0.020 | 0.147 ± 0.021 | 0.151 ± 0.019 | **0.444 ± 0.057** | **0.147 ± 0.019** | 0.146 ± 0.020 |

Table S10. Within-cohort (ADNI) clinical diagnosis prediction performance under different modality ablation scenarios.

Results were averaged across 20 trained models (obtained from training with all data in ADNI). Higher mAUC indicates better performance. The best result for each ablation scenario is bolded. Due to model design, AD-Map does not utilize diagnostic input features and therefore has no results in the “Ablate DX” condition, which is marked as “N.A.”

| Diagnostic mAUC ↑ | No ablation | Ablate DX | Ablate COG | Ablate MRI | Ablate CSF | Ablate PET |
| --- | --- | --- | --- | --- | --- | --- |
| MRNN | 0.926 ± 0.028 | 0.831 ± 0.045 | 0.836 ± 0.085 | 0.922 ± 0.031 | 0.926 ± 0.031 | 0.925 ± 0.030 |
| AD-Map | 0.815 ± 0.028 | N.A. | 0.731 ± 0.068 | 0.819 ± 0.031 | 0.816 ± 0.029 | 0.816 ± 0.027 |
| L2C-XGB | 0.954 ± 0.023 | 0.868 ± 0.036 | 0.916 ± 0.040 | 0.947 ± 0.023 | 0.951 ± 0.025 | **0.953 ± 0.024** |
| L2C-FNN | 0.945 ± 0.025 | 0.900 ± 0.033 | 0.894 ± 0.036 | 0.943 ± 0.029 | 0.945 ± 0.024 | 0.942 ± 0.025 |
| L2C-MICE | 0.944 ± 0.027 | 0.890 ± 0.034 | 0.818 ± 0.036 | 0.935 ± 0.029 | 0.938 ± 0.026 | 0.939 ± 0.028 |
| L2C-PFN | **0.955 ± 0.024** | 0.907 ± 0.037 | 0.934 ± 0.033 | **0.953 ± 0.025** | 0.951 ± 0.023 | 0.950 ± 0.025 |
| ProTuse-TTA | 0.954 ± 0.020 | **0.936 ± 0.027** | **0.940 ± 0.028** | **0.953 ± 0.022** | **0.954 ± 0.021** | 0.951 ± 0.021 |

Table S11. Within-cohort (ADNI) MMSE MAE increase compared with “full-set” baseline under different modality ablation scenarios.

Results were averaged across 20 trained models (obtained from training with all data in ADNI). Lower MAE increase indicates a smaller performance drop. The smallest drop for each ablation scenario is bolded. Due to model design, AD-Map does not utilize diagnostic input features and therefore has no results in the “Ablate DX” condition, which is marked as “N.A.”

| MMSE MAE Increase ↓ | Ablate DX | Ablate COG | Ablate MRI | Ablate CSF | Ablate PET |
| --- | --- | --- | --- | --- | --- |
| MRNN | 0.016 ± 0.041 | 0.914 ± 0.422 | 0.005 ± 0.089 | 0.016 ± 0.066 | 0.021 ± 0.049 |
| AD-Map | N.A. | 0.628 ± 0.287 | 0.016 ± 0.175 | 0.004 ± 0.031 | **0.003 ± 0.012** |
| L2C-XGB | 0.119 ± 0.114 | 2.218 ± 0.974 | 0.029 ± 0.127 | -0.007 ± 0.049 | 0.015 ± 0.065 |
| L2C-FNN | 0.041 ± 0.075 | 0.950 ± 0.379 | 0.066 ± 0.113 | **-0.012 ± 0.090** | 0.004 ± 0.077 |
| L2C-MICE | 0.016 ± 0.060 | 1.140 ± 0.400 | 0.258 ± 0.196 | 0.030 ± 0.090 | 0.042 ± 0.080 |
| L2C-PFN | 0.017 ± 0.030 | 0.923 ± 0.408 | 0.041 ± 0.062 | 0.021 ± 0.049 | 0.030 ± 0.057 |
| ProTuse-TTA | **0.005 ± 0.023** | **0.391 ± 0.214** | **-0.021 ± 0.048** | 0.002 ± 0.048 | 0.022 ± 0.038 |

Table S12. Within-cohort (ADNI) hippocampus volume MAE increase compared with “full-set” baseline under different modality ablation scenarios.

Results were averaged across 20 trained models (obtained from training with all data in ADNI). Lower MAE increase indicates a smaller performance drop. The smallest drop for each ablation scenario is bolded. Due to model design, AD-Map does not utilize diagnostic input features and therefore has no results in the “Ablate DX” condition, which is marked as “N.A.”

| Hippocampus MAE (1e-4) Increase ↓ | Ablate DX | Ablate COG | Ablate MRI | Ablate CSF | Ablate PET |
| --- | --- | --- | --- | --- | --- |
| MRNN | 0.003 ± 0.062 | **-0.020 ± 0.101** | 4.483 ± 0.743 | 0.016 ± 0.060 | **-0.035 ± 0.070** |
| AD-Map | N.A. | 0.040 ± 0.107 | 3.166 ± 0.721 | **-0.001 ± 0.020** | -0.003 ± 0.008 |
| L2C-XGB | 0.003 ± 0.012 | 0.235 ± 0.107 | 4.177 ± 0.716 | 0.029 ± 0.043 | 0.039 ± 0.043 |
| L2C-FNN | 0.025 ± 0.042 | 0.090 ± 0.120 | 3.994 ± 0.714 | 0.031 ± 0.076 | 0.003 ± 0.059 |
| L2C-MICE | 0.025 ± 0.039 | 0.194 ± 0.134 | 5.021 ± 0.601 | 0.134 ± 0.066 | 0.025 ± 0.050 |
| L2C-PFN | **-0.006 ± 0.017** | 0.037 ± 0.051 | 3.592 ± 0.670 | 0.085 ± 0.088 | 0.012 ± 0.034 |
| ProTuse-TTA | 0.000 ± 0.019 | 0.045 ± 0.073 | **2.970 ± 0.522** | 0.001 ± 0.037 | -0.008 ± 0.031 |

Table S13. Within-cohort (ADNI) clinical diagnosis mAUC reduction compared with “full-set” baseline under different modality ablation scenarios.

Results were averaged across 20 trained models (obtained from training with all data in ADNI). Lower mAUC reduction indicates a smaller performance drop. The smallest drop for each ablation scenario is bolded. Due to model design, AD-Map does not utilize diagnostic input features and therefore has no results in the “Ablate DX” condition, which is marked as “N.A.”

| Diagnostic mAUC Drop ↓ | Ablate DX | Ablate COG | Ablate MRI | Ablate CSF | Ablate PET |
| --- | --- | --- | --- | --- | --- |
| MRNN | 0.094 ± 0.041 | 0.090 ± 0.079 | 0.004 ± 0.014 | -0.000 ± 0.010 | 0.001 ± 0.010 |
| AD-Map | N.A. | 0.084 ± 0.065 | **-0.004 ± 0.023** | **-0.001 ± 0.010** | **-0.001 ± 0.006** |
| L2C-XGB | 0.086 ± 0.029 | 0.038 ± 0.028 | 0.007 ± 0.010 | 0.003 ± 0.009 | 0.001 ± 0.006 |
| L2C-FNN | 0.045 ± 0.025 | 0.052 ± 0.025 | 0.002 ± 0.011 | 0.000 ± 0.009 | 0.004 ± 0.005 |
| L2C-MICE | 0.055 ± 0.026 | 0.126 ± 0.030 | 0.009 ± 0.014 | 0.006 ± 0.012 | 0.005 ± 0.005 |
| L2C-PFN | 0.048 ± 0.024 | 0.020 ± 0.018 | 0.001 ± 0.009 | 0.003 ± 0.009 | 0.004 ± 0.005 |
| ProTuse-TTA | **0.018 ± 0.017** | **0.014 ± 0.016** | 0.001 ± 0.008 | 0.000 ± 0.004 | 0.003 ± 0.005 |

Table S14. Cross-cohort MMSE prediction performance using different numbers of input timepoints

averaged across 20 trained models (training with all timepoints in ADNI). Lower MAE indicates better performance. The best result for each input horizon (i.e., number of input timepoints) on each test dataset is bolded. Due to dataset constraints, the maximum number of input timepoints for each participant is only 2 for AIBL and 3 for MACC. Therefore, results for AIBL with 3 and 4 timepoints and MACC with 4 timepoints are marked as “N.A.”

| MMSE MAE ↓ | 1 timepoints | 2 timepoints | 3 timepoints | 4 timepoints |
| --- | --- | --- | --- | --- |
| AIBL | | | | |
| MRNN | 1.571 ± 0.079 | 1.435 ± 0.067 | N.A. | N.A. |
| AD-Map | 1.524 ± 0.008 | 1.488 ± 0.048 | N.A. | N.A. |
| L2C-XGB | 3.325 ± 1.043 | 3.079 ± 0.981 | N.A. | N.A. |
| L2C-FNN | 1.592 ± 0.020 | 1.511 ± 0.026 | N.A. | N.A. |
| L2C-MICE | 1.713 ± 0.050 | 1.634 ± 0.067 | N.A. | N.A. |
| L2C-PFN | 1.594 ± 0.008 | 1.489 ± 0.014 | N.A. | N.A. |
| ProTuse-TTA | **1.462 ± 0.036** | **1.374 ± 0.033** | N.A. | N.A. |
| MACC | | | | |
| MRNN | 3.310 ± 0.343 | 2.931 ± 0.314 | 2.519 ± 0.272 | N.A. |
| AD-Map | **2.975 ± 0.050** | **2.599 ± 0.015** | **2.246 ± 0.031** | N.A. |
| L2C-XGB | 4.217 ± 0.624 | 4.008 ± 0.641 | 3.767 ± 0.646 | N.A. |
| L2C-FNN | 5.062 ± 0.299 | 4.897 ± 0.344 | 4.828 ± 0.348 | N.A. |
| L2C-MICE | 4.781 ± 0.239 | 4.648 ± 0.279 | 4.653 ± 0.308 | N.A. |
| L2C-PFN | 4.118 ± 0.200 | 3.858 ± 0.224 | 3.703 ± 0.231 | N.A. |
| ProTuse-TTA | 3.070 ± 0.077 | 2.611 ± 0.080 | 2.272 ± 0.057 | N.A. |
| OASIS | | | | |
| MRNN | 2.660 ± 1.031 | 2.664 ± 1.019 | 2.543 ± 0.918 | 2.508 ± 0.878 |
| AD-Map | 1.372 ± 0.013 | 1.323 ± 0.013 | 1.321 ± 0.018 | 1.347 ± 0.017 |
| L2C-XGB | 4.739 ± 1.415 | 4.713 ± 1.470 | 4.603 ± 1.430 | 4.442 ± 1.346 |
| L2C-FNN | **1.333 ± 0.028** | **1.292 ± 0.030** | **1.278 ± 0.025** | **1.248 ± 0.020** |
| L2C-MICE | 1.495 ± 0.035 | 1.439 ± 0.041 | 1.424 ± 0.047 | 1.395 ± 0.039 |
| L2C-PFN | 1.346 ± 0.011 | 1.300 ± 0.009 | 1.287 ± 0.012 | 1.268 ± 0.013 |
| ProTuse-TTA | 1.565 ± 0.062 | 1.421 ± 0.060 | 1.360 ± 0.051 | 1.300 ± 0.039 |

Table S15. Cross-cohort hippocampus volume prediction performance using different numbers of input timepoints

averaged across 20 trained models (training with all timepoints in ADNI). Lower MAE indicates better performance. The best result for each input horizon (i.e., number of input timepoints) on each test dataset is bolded. Due to dataset constraints, the maximum number of input timepoints for each participant is only 2 for AIBL and 3 for MACC. Therefore, results for AIBL with 3 and 4 timepoints and MACC with 4 timepoints are marked as “N.A.”

| Hippocampus MAE (1e-3) ↓ | 1 timepoints | 2 timepoints | 3 timepoints | 4 timepoints |
| --- | --- | --- | --- | --- |
| AIBL | | | | |
| MRNN | 0.301 ± 0.023 | 0.207 ± 0.017 | N.A. | N.A. |
| AD-Map | 0.247 ± 0.004 | 0.229 ± 0.011 | N.A. | N.A. |
| L2C-XGB | 0.262 ± 0.010 | 0.194 ± 0.008 | N.A. | N.A. |
| L2C-FNN | 0.244 ± 0.003 | 0.181 ± 0.005 | N.A. | N.A. |
| L2C-MICE | 0.271 ± 0.010 | 0.202 ± 0.007 | N.A. | N.A. |
| L2C-PFN | 0.253 ± 0.008 | 0.184 ± 0.003 | N.A. | N.A. |
| ProTuse-TTA | **0.226 ± 0.004** | **0.175 ± 0.005** | N.A. | N.A. |
| MACC | | | | |
| MRNN | 0.258 ± 0.053 | 0.262 ± 0.058 | 0.216 ± 0.040 | N.A. |
| AD-Map | 0.248 ± 0.007 | 0.241 ± 0.003 | 0.207 ± 0.002 | N.A. |
| L2C-XGB | 0.263 ± 0.029 | 0.267 ± 0.029 | 0.211 ± 0.013 | N.A. |
| L2C-FNN | 0.243 ± 0.009 | 0.247 ± 0.009 | 0.202 ± 0.008 | N.A. |
| L2C-MICE | 0.272 ± 0.013 | 0.274 ± 0.014 | 0.227 ± 0.011 | N.A. |
| L2C-PFN | **0.225 ± 0.003** | **0.224 ± 0.003** | **0.189 ± 0.002** | N.A. |
| ProTuse-TTA | 0.227 ± 0.005 | 0.225 ± 0.005 | 0.195 ± 0.004 | N.A. |
| OASIS | | | | |
| MRNN | 0.767 ± 0.097 | 0.631 ± 0.081 | 0.528 ± 0.069 | 0.484 ± 0.064 |
| AD-Map | 0.428 ± 0.007 | **0.340 ± 0.004** | 0.317 ± 0.003 | 0.297 ± 0.005 |
| L2C-XGB | 0.503 ± 0.012 | 0.408 ± 0.013 | 0.372 ± 0.014 | 0.352 ± 0.015 |
| L2C-FNN | 0.495 ± 0.010 | 0.391 ± 0.006 | 0.334 ± 0.004 | 0.311 ± 0.003 |
| L2C-MICE | 0.551 ± 0.012 | 0.435 ± 0.013 | 0.376 ± 0.010 | 0.351 ± 0.013 |
| L2C-PFN | 0.479 ± 0.022 | 0.390 ± 0.020 | 0.363 ± 0.019 | 0.342 ± 0.018 |
| ProTuse-TTA | **0.418 ± 0.006** | 0.341 ± 0.006 | **0.309 ± 0.004** | **0.292 ± 0.004** |

Table S16. Cross-cohort diagnosis prediction performance using different numbers of input timepoints

averaged across 20 trained models (training with all timepoints in ADNI). Higher mAUC indicates better performance. The best result for each input horizon (i.e., number of input timepoints) on each test dataset is bolded. Due to dataset constraints, the maximum number of input timepoints for each participant is only 2 for AIBL and 3 for MACC. Therefore, results for AIBL with 3 and 4 timepoints and MACC with 4 timepoints are marked as “N.A.”

| mAUC ↑ | 1 timepoints | 2 timepoints | 3 timepoints | 4 timepoints |
| --- | --- | --- | --- | --- |
| AIBL | | | | |
| MRNN | 0.797 ± 0.021 | 0.874 ± 0.019 | N.A. | N.A. |
| AD-Map | 0.723 ± 0.010 | 0.796 ± 0.011 | N.A. | N.A. |
| L2C-XGB | 0.800 ± 0.018 | 0.893 ± 0.011 | N.A. | N.A. |
| L2C-FNN | 0.801 ± 0.013 | 0.860 ± 0.013 | N.A. | N.A. |
| L2C-MICE | 0.764 ± 0.016 | 0.832 ± 0.020 | N.A. | N.A. |
| L2C-PFN | 0.815 ± 0.010 | 0.901 ± 0.006 | N.A. | N.A. |
| ProTuse-TTA | **0.846 ± 0.011** | **0.913 ± 0.006** | N.A. | N.A. |
| MACC | | | | |
| MRNN | 0.839 ± 0.024 | 0.856 ± 0.026 | 0.882 ± 0.030 | N.A. |
| AD-Map | 0.795 ± 0.007 | 0.802 ± 0.007 | 0.810 ± 0.007 | N.A. |
| L2C-XGB | 0.879 ± 0.007 | 0.901 ± 0.007 | 0.957 ± 0.006 | N.A. |
| L2C-FNN | 0.881 ± 0.010 | 0.904 ± 0.008 | 0.946 ± 0.008 | N.A. |
| L2C-MICE | 0.856 ± 0.014 | 0.881 ± 0.016 | 0.926 ± 0.018 | N.A. |
| L2C-PFN | **0.886 ± 0.004** | **0.905 ± 0.003** | **0.963 ± 0.002** | N.A. |
| ProTuse-TTA | 0.878 ± 0.005 | 0.900 ± 0.005 | 0.955 ± 0.005 | N.A. |
| OASIS | | | | |
| MRNN | 0.590 ± 0.015 | 0.612 ± 0.015 | 0.636 ± 0.016 | 0.651 ± 0.016 |
| AD-Map | **0.659 ± 0.004** | **0.681 ± 0.003** | **0.688 ± 0.005** | 0.704 ± 0.005 |
| L2C-XGB | 0.642 ± 0.009 | 0.664 ± 0.009 | 0.674 ± 0.014 | **0.709 ± 0.012** |
| L2C-FNN | 0.624 ± 0.012 | 0.637 ± 0.022 | 0.644 ± 0.025 | 0.667 ± 0.027 |
| L2C-MICE | 0.584 ± 0.019 | 0.594 ± 0.028 | 0.603 ± 0.028 | 0.624 ± 0.028 |
| L2C-PFN | 0.627 ± 0.008 | 0.645 ± 0.013 | 0.656 ± 0.013 | 0.702 ± 0.011 |
| ProTuse-TTA | 0.617 ± 0.017 | 0.659 ± 0.017 | 0.676 ± 0.016 | 0.699 ± 0.014 |

Table S17. Cross-cohort MMSE prediction performance broken down into yearly intervals up to 6 years into the future.

Results were averaged across 20 trained models (from ADNI). Lower MAE indicates better performance. The best result for each yearly interval (i.e., prediction horizon) on each test dataset is bolded.

| MMSE MAE ↓ | 0-1 | 1-2 | 2-3 | 3-4 | 4-5 | 5-6 |
| --- | --- | --- | --- | --- | --- | --- |
| AIBL | | | | | | |
| MRNN | 1.179 ± 0.204 | 1.333 ± 0.081 | 1.634 ± 0.118 | 2.351 ± 0.221 | 1.487 ± 0.099 | **2.826 ± 0.267** |
| AD-Map | **0.300 ± 0.029** | **1.237 ± 0.010** | 1.556 ± 0.039 | 2.215 ± 0.047 | 1.574 ± 0.078 | 2.841 ± 0.211 |
| L2C-XGB | 3.199 ± 0.914 | 2.519 ± 0.834 | 3.015 ± 0.874 | 3.524 ± 0.784 | 3.569 ± 1.188 | 3.710 ± 0.817 |
| L2C-FNN | 1.193 ± 0.297 | 1.544 ± 0.045 | 1.959 ± 0.059 | 2.979 ± 0.091 | 1.511 ± 0.049 | 2.864 ± 0.098 |
| L2C-MICE | 1.275 ± 0.413 | 1.666 ± 0.054 | 2.055 ± 0.081 | 3.110 ± 0.089 | 1.649 ± 0.082 | 2.900 ± 0.322 |
| L2C-PFN | 1.113 ± 0.061 | 1.448 ± 0.023 | 1.875 ± 0.032 | 2.847 ± 0.046 | **1.445 ± 0.014** | 2.831 ± 0.041 |
| ProTuse-TTA | 1.082 ± 0.129 | 1.308 ± 0.029 | **1.498 ± 0.046** | **2.016 ± 0.067** | 1.447 ± 0.054 | 2.912 ± 0.209 |
| MACC | | | | | | |
| MRNN | 2.192 ± 0.298 | 2.644 ± 0.363 | 2.907 ± 0.337 | 3.447 ± 0.456 | 3.797 ± 0.466 | 2.786 ± 1.382 |
| AD-Map | **2.057 ± 0.018** | **2.329 ± 0.015** | **2.616 ± 0.039** | 2.966 ± 0.087 | 2.817 ± 0.105 | **1.915 ± 0.256** |
| L2C-XGB | 3.401 ± 0.558 | 3.763 ± 0.509 | 4.148 ± 0.623 | 4.766 ± 0.593 | 5.599 ± 0.810 | 3.541 ± 0.743 |
| L2C-FNN | 4.899 ± 0.504 | 5.283 ± 0.449 | 5.276 ± 0.377 | 6.358 ± 0.426 | 5.972 ± 0.343 | 4.575 ± 0.405 |
| L2C-MICE | 5.025 ± 0.424 | 5.208 ± 0.389 | 5.034 ± 0.316 | 5.882 ± 0.411 | 5.572 ± 0.410 | 4.056 ± 0.770 |
| L2C-PFN | 3.768 ± 0.276 | 4.083 ± 0.261 | 4.026 ± 0.248 | 4.878 ± 0.283 | 4.767 ± 0.264 | 3.616 ± 0.291 |
| ProTuse-TTA | 2.228 ± 0.043 | 2.481 ± 0.043 | 2.655 ± 0.049 | **2.940 ± 0.094** | **2.563 ± 0.143** | 2.000 ± 0.262 |
| OASIS | | | | | | |
| MRNN | 1.380 ± 0.102 | 1.602 ± 0.134 | 1.901 ± 0.186 | 2.073 ± 0.279 | 2.563 ± 0.673 | 3.429 ± 1.241 |
| AD-Map | 1.246 ± 0.012 | 1.402 ± 0.019 | 1.577 ± 0.015 | 1.565 ± 0.021 | 1.690 ± 0.037 | 1.803 ± 0.031 |
| L2C-XGB | 3.015 ± 0.877 | 3.019 ± 0.855 | 3.250 ± 0.934 | 3.417 ± 1.106 | 3.736 ± 1.229 | 3.461 ± 1.218 |
| L2C-FNN | 1.518 ± 0.055 | 1.727 ± 0.061 | 1.927 ± 0.054 | 1.840 ± 0.046 | 1.714 ± 0.048 | 1.798 ± 0.045 |
| L2C-MICE | 1.624 ± 0.077 | 1.808 ± 0.040 | 2.007 ± 0.055 | 1.938 ± 0.064 | 1.825 ± 0.053 | 1.861 ± 0.068 |
| L2C-PFN | 1.413 ± 0.033 | 1.640 ± 0.028 | 1.844 ± 0.032 | 1.792 ± 0.024 | 1.720 ± 0.020 | 1.754 ± 0.012 |
| ProTuse-TTA | **1.189 ± 0.029** | **1.377 ± 0.024** | **1.524 ± 0.019** | **1.482 ± 0.026** | **1.574 ± 0.040** | **1.709 ± 0.070** |

Table S18. Cross-cohort hippocampus volume prediction performance broken down into yearly intervals up to 6 years into the future.

Results were averaged across 20 trained models (from ADNI). Lower MAE indicates better performance. The best result for each yearly interval (i.e., prediction horizon) on each test dataset is bolded.

| Hippocampus  MAE ↓ | 0-1 | 1-2 | 2-3 | 3-4 | 4-5 | 5-6 |
| --- | --- | --- | --- | --- | --- | --- |
| AIBL | | | | | | |
| MRNN | 0.124 ± 0.023 | 0.141 ± 0.008 | 0.311 ± 0.021 | 0.197 ± 0.011 | 0.271 ± 0.035 | 0.260 ± 0.023 |
| AD-Map | 0.094 ± 0.008 | 0.141 ± 0.004 | 0.336 ± 0.011 | 0.227 ± 0.008 | 0.286 ± 0.011 | 0.229 ± 0.025 |
| L2C-XGB | **0.064 ± 0.033** | 0.152 ± 0.006 | 0.310 ± 0.010 | 0.219 ± 0.013 | 0.220 ± 0.010 | 0.237 ± 0.025 |
| L2C-FNN | 0.122 ± 0.042 | 0.137 ± 0.003 | 0.290 ± 0.009 | 0.178 ± 0.008 | **0.202 ± 0.006** | **0.198 ± 0.012** |
| L2C-MICE | 0.105 ± 0.054 | 0.166 ± 0.009 | 0.303 ± 0.011 | 0.212 ± 0.011 | 0.228 ± 0.010 | 0.226 ± 0.030 |
| L2C-PFN | 0.135 ± 0.010 | 0.140 ± 0.002 | 0.293 ± 0.003 | **0.177 ± 0.003** | 0.212 ± 0.008 | 0.220 ± 0.011 |
| ProTuse-TTA | 0.146 ± 0.034 | **0.134 ± 0.004** | **0.257 ± 0.006** | 0.199 ± 0.008 | 0.203 ± 0.008 | 0.249 ± 0.025 |
| MACC | | | | | | |
| MRNN | 0.192 ± 0.036 | 0.199 ± 0.026 | 0.230 ± 0.063 | 0.255 ± 0.062 | 0.285 ± 0.070 | 0.538 ± 0.089 |
| AD-Map | 0.181 ± 0.006 | 0.209 ± 0.002 | 0.224 ± 0.002 | 0.208 ± 0.010 | 0.213 ± 0.008 | 0.389 ± 0.020 |
| L2C-XGB | 0.187 ± 0.011 | 0.222 ± 0.014 | 0.229 ± 0.018 | 0.213 ± 0.022 | 0.253 ± 0.024 | **0.191 ± 0.024** |
| L2C-FNN | 0.174 ± 0.006 | 0.196 ± 0.008 | 0.217 ± 0.009 | 0.191 ± 0.010 | 0.213 ± 0.014 | 0.262 ± 0.015 |
| L2C-MICE | 0.200 ± 0.014 | 0.224 ± 0.014 | 0.238 ± 0.015 | 0.224 ± 0.018 | 0.232 ± 0.017 | 0.327 ± 0.119 |
| L2C-PFN | **0.165 ± 0.004** | **0.187 ± 0.002** | **0.194 ± 0.003** | 0.205 ± 0.009 | **0.203 ± 0.011** | 0.212 ± 0.032 |
| ProTuse-TTA | 0.168 ± 0.007 | 0.201 ± 0.004 | 0.195 ± 0.005 | **0.183 ± 0.014** | 0.212 ± 0.020 | 0.265 ± 0.042 |
| OASIS | | | | | | |
| MRNN | 0.282 ± 0.014 | 0.298 ± 0.021 | 0.314 ± 0.039 | 0.412 ± 0.045 | 0.475 ± 0.077 | 0.632 ± 0.110 |
| AD-Map | 0.211 ± 0.004 | 0.230 ± 0.005 | 0.249 ± 0.006 | 0.309 ± 0.004 | 0.335 ± 0.008 | 0.402 ± 0.006 |
| L2C-XGB | 0.241 ± 0.006 | 0.272 ± 0.007 | 0.279 ± 0.006 | 0.330 ± 0.007 | 0.337 ± 0.010 | 0.439 ± 0.012 |
| L2C-FNN | 0.249 ± 0.006 | 0.248 ± 0.006 | 0.248 ± 0.007 | 0.312 ± 0.005 | 0.309 ± 0.006 | 0.408 ± 0.005 |
| L2C-MICE | 0.286 ± 0.014 | 0.282 ± 0.015 | 0.280 ± 0.011 | 0.344 ± 0.020 | 0.352 ± 0.022 | 0.444 ± 0.016 |
| L2C-PFN | 0.207 ± 0.006 | 0.251 ± 0.012 | 0.259 ± 0.011 | 0.321 ± 0.008 | 0.355 ± 0.015 | 0.439 ± 0.018 |
| ProTuse-TTA | **0.189 ± 0.004** | **0.216 ± 0.004** | **0.223 ± 0.005** | **0.274 ± 0.004** | **0.272 ± 0.008** | **0.378 ± 0.010** |

Table S19. Cross-cohort clinical diagnosis prediction performance broken down into yearly intervals up to 6 years into the future.

Results were averaged across 20 trained models (from ADNI). Higher mAUC indicates better performance. The best result for each yearly interval (i.e., prediction horizon) on each test dataset is bolded. Due to dataset constraints, AIBL only had one diagnostic class in year 0-1, making mAUC undefined in this case. Therefore, results for AIBL at year 0-1 is marked as “N.A.”

| mAUC ↑ | 0-1 | 1-2 | 2-3 | 3-4 | 4-5 | 5-6 |
| --- | --- | --- | --- | --- | --- | --- |
| AIBL | | | | | | |
| MRNN | N.A. | 0.877 ± 0.019 | 0.919 ± 0.024 | 0.845 ± 0.023 | 0.819 ± 0.029 | 0.665 ± 0.068 |
| AD-Map | N.A. | 0.826 ± 0.010 | 0.811 ± 0.022 | 0.766 ± 0.012 | 0.791 ± 0.016 | **0.802 ± 0.046** |
| L2C-XGB | N.A. | 0.915 ± 0.005 | 0.919 ± 0.015 | 0.867 ± 0.013 | 0.825 ± 0.021 | 0.730 ± 0.053 |
| L2C-FNN | N.A. | 0.878 ± 0.010 | 0.908 ± 0.015 | 0.821 ± 0.022 | 0.791 ± 0.020 | 0.666 ± 0.041 |
| L2C-MICE | N.A. | 0.870 ± 0.012 | 0.876 ± 0.029 | 0.805 ± 0.028 | 0.762 ± 0.036 | 0.687 ± 0.093 |
| L2C-PFN | N.A. | 0.907 ± 0.008 | 0.902 ± 0.010 | 0.851 ± 0.009 | **0.849 ± 0.016** | 0.774 ± 0.057 |
| ProTuse-TTA | N.A. | **0.926 ± 0.006** | **0.950 ± 0.008** | **0.876 ± 0.012** | 0.836 ± 0.012 | 0.688 ± 0.084 |
| MACC | | | | | | |
| MRNN | 0.934 ± 0.022 | 0.899 ± 0.026 | 0.884 ± 0.028 | 0.866 ± 0.034 | 0.862 ± 0.046 | 0.819 ± 0.136 |
| AD-Map | 0.859 ± 0.008 | 0.823 ± 0.008 | 0.829 ± 0.006 | 0.773 ± 0.011 | 0.782 ± 0.027 | 0.696 ± 0.043 |
| L2C-XGB | 0.983 ± 0.003 | **0.958 ± 0.007** | 0.934 ± 0.006 | 0.933 ± 0.012 | 0.899 ± 0.017 | 0.965 ± 0.015 |
| L2C-FNN | 0.979 ± 0.004 | 0.952 ± 0.006 | 0.932 ± 0.006 | **0.948 ± 0.007** | 0.922 ± 0.016 | **1.000 ± 0.000** |
| L2C-MICE | 0.960 ± 0.014 | 0.934 ± 0.011 | 0.910 ± 0.016 | 0.917 ± 0.020 | 0.896 ± 0.032 | 0.982 ± 0.028 |
| L2C-PFN | **0.985 ± 0.001** | **0.958 ± 0.002** | **0.943 ± 0.002** | **0.948 ± 0.003** | 0.896 ± 0.008 | 0.996 ± 0.010 |
| ProTuse-TTA | 0.983 ± 0.003 | 0.954 ± 0.003 | 0.935 ± 0.005 | 0.928 ± 0.008 | **0.938 ± 0.012** | 0.935 ± 0.024 |
| OASIS | | | | | | |
| MRNN | 0.846 ± 0.031 | 0.792 ± 0.020 | 0.777 ± 0.034 | 0.735 ± 0.031 | 0.647 ± 0.030 | 0.627 ± 0.028 |
| AD-Map | 0.833 ± 0.018 | 0.816 ± 0.013 | 0.811 ± 0.011 | 0.770 ± 0.011 | 0.687 ± 0.010 | 0.583 ± 0.014 |
| L2C-XGB | 0.832 ± 0.025 | **0.871 ± 0.022** | 0.855 ± 0.016 | 0.780 ± 0.019 | 0.726 ± 0.023 | 0.664 ± 0.024 |
| L2C-FNN | 0.860 ± 0.019 | 0.806 ± 0.022 | 0.822 ± 0.013 | 0.745 ± 0.024 | 0.680 ± 0.032 | 0.622 ± 0.038 |
| L2C-MICE | 0.807 ± 0.043 | 0.785 ± 0.024 | 0.764 ± 0.029 | 0.703 ± 0.028 | 0.658 ± 0.034 | 0.614 ± 0.036 |
| L2C-PFN | 0.895 ± 0.014 | 0.858 ± 0.011 | 0.849 ± 0.008 | 0.750 ± 0.018 | 0.729 ± 0.009 | 0.651 ± 0.014 |
| ProTuse-TTA | **0.897 ± 0.009** | 0.851 ± 0.014 | **0.865 ± 0.012** | **0.799 ± 0.017** | **0.739 ± 0.012** | **0.657 ± 0.027** |

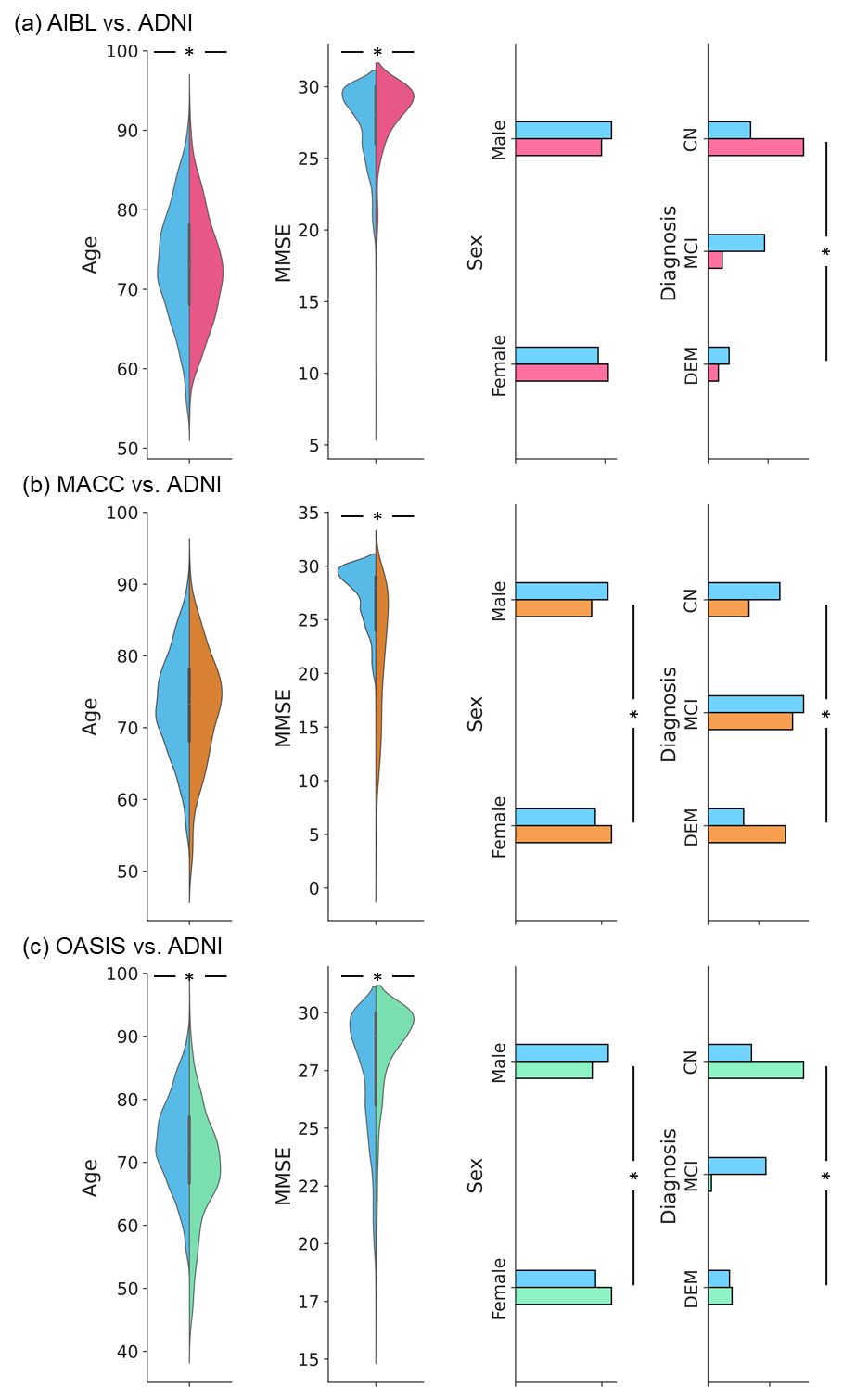

F**igure S1.** Baseline age, MMSE, clinical diagnosis, and sex distribution differences between ADNI and external test set.

(a) Distributions of age, sex, MMSE and clinical diagnosis for ADNI (blue) and AIBL (pink). (b) Distributions of age, sex, MMSE and clinical diagnosis for ADNI (blue) and MACC (yellow). (c) Distributions of age, sex, MMSE and clinical diagnosis for ADNI (blue) and OASIS (green). To test for differences in mean age and mean MMSE between ADNI and the external dataset, a permutation test was used. To test for differences in distributions of sex and diagnosis between ADNI and the external dataset, the chi square test was used. * indicates statistical significance after correcting for multiple comparisons with false discovery rate (FDR) q < 0.05. See Table 2 in the main text for the full set of statistical tests.

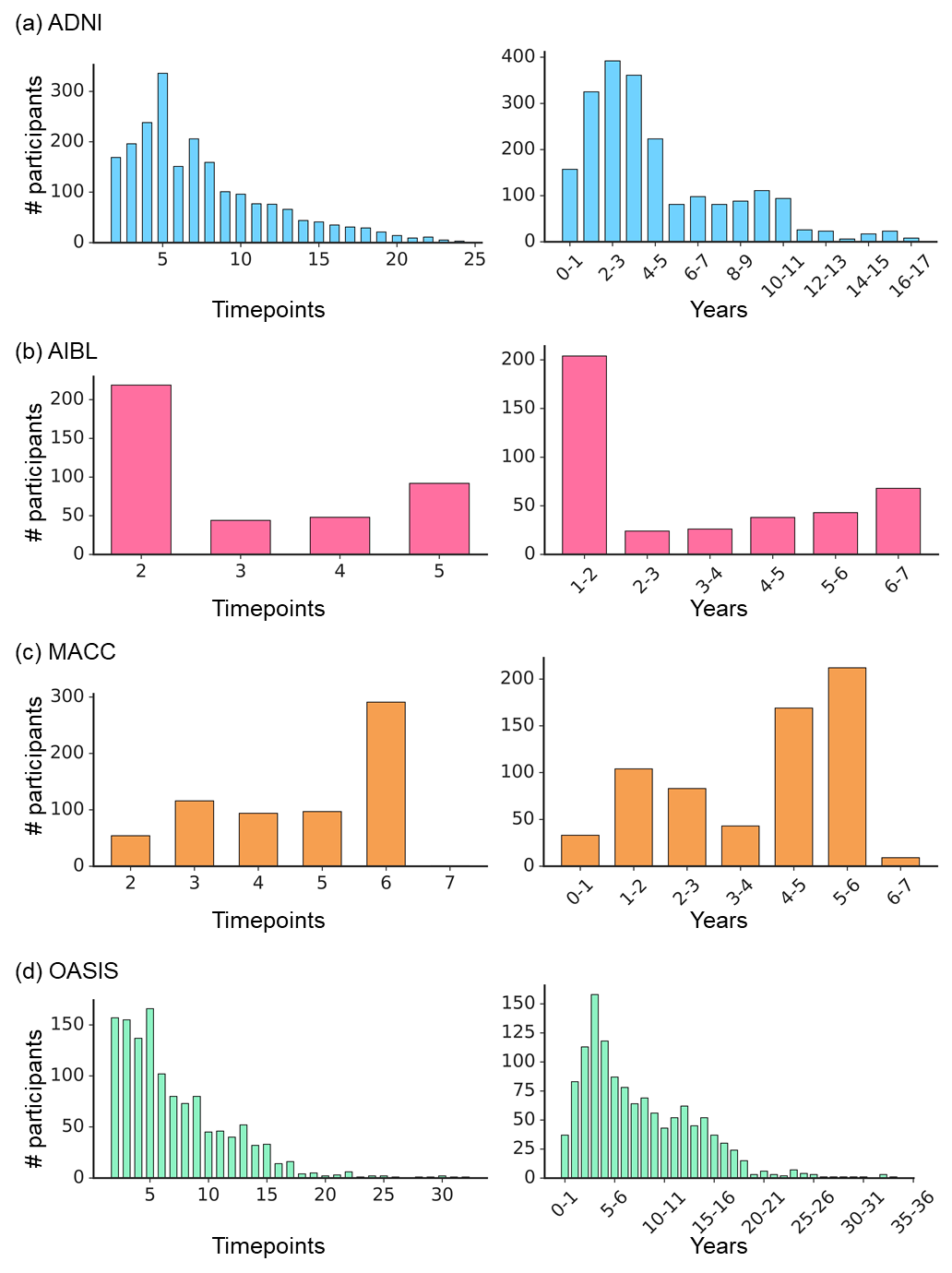

Figure S2. Participants longitudinal follow-up distributions.

Left: Distribution of the number of timepoints for all participants in each dataset. Right: Distribution of the number of years between baseline and the last observation for all participants in each dataset. Note that year 0-1 means 0<t≤12, where t is the interval between baseline and the last observed timepoint; year 1-2 means 12<t≤24, and so on. (a) ADNI. (b) AIBL. (c) MACC. (d) OASIS.

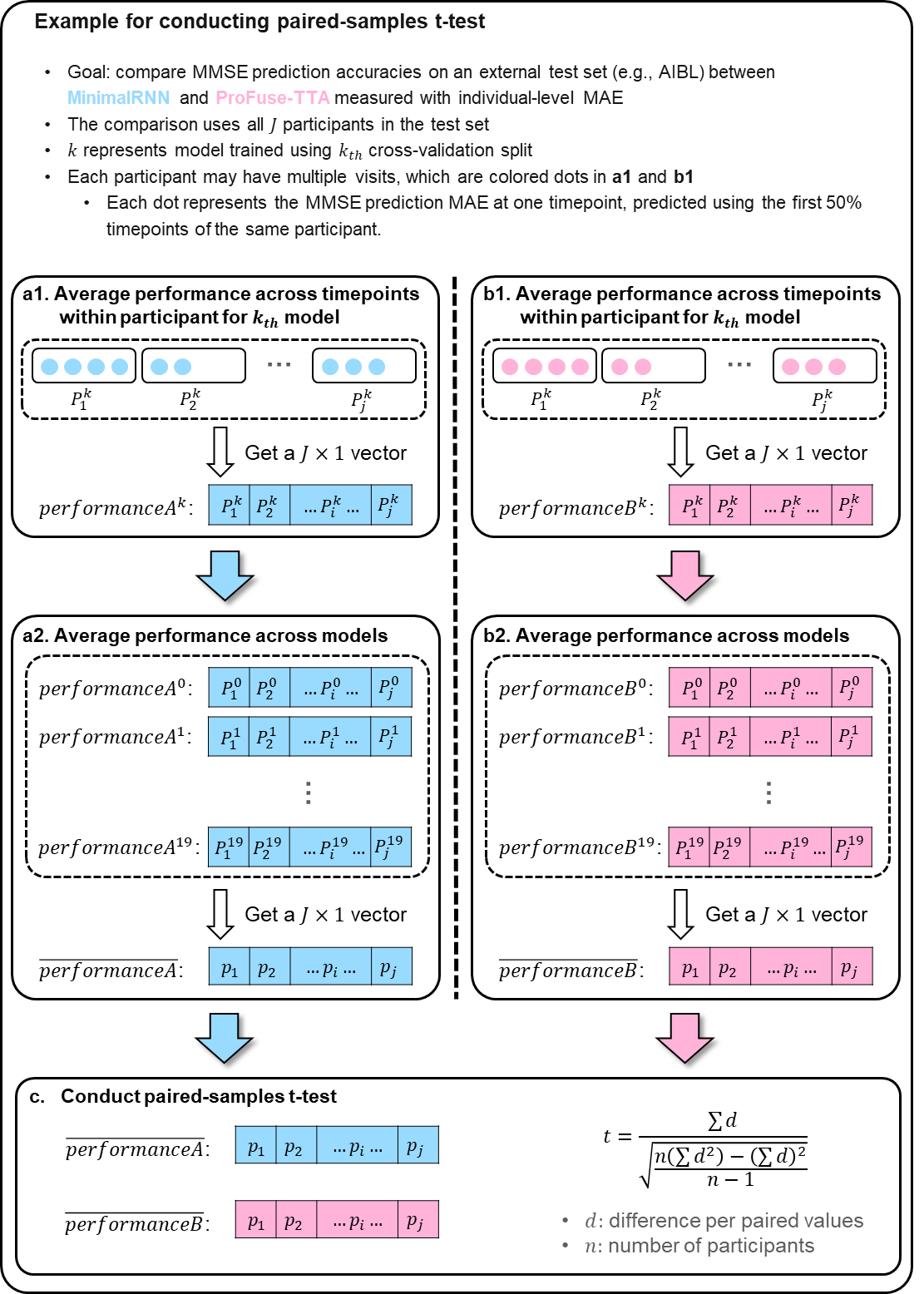

Figure S3. Illustration of paired-samples t-test for comparing performance measured with individual-level metrics

(e.g., MMSE MAE) of MinimalRNN and ProFuse-TTA on an external test set. (a1) For a given model, we averaged the MMSE MAE within each participant across all timepoints for MinimalRNN. (a2) Averaging the participant-level MMSE MAE obtained in (a1) across the 20 models. (b1 & b2) Same as a1 and a2 but for ProFuse-TTA. (c) Conduct paired-samples t-test to obtain p value.

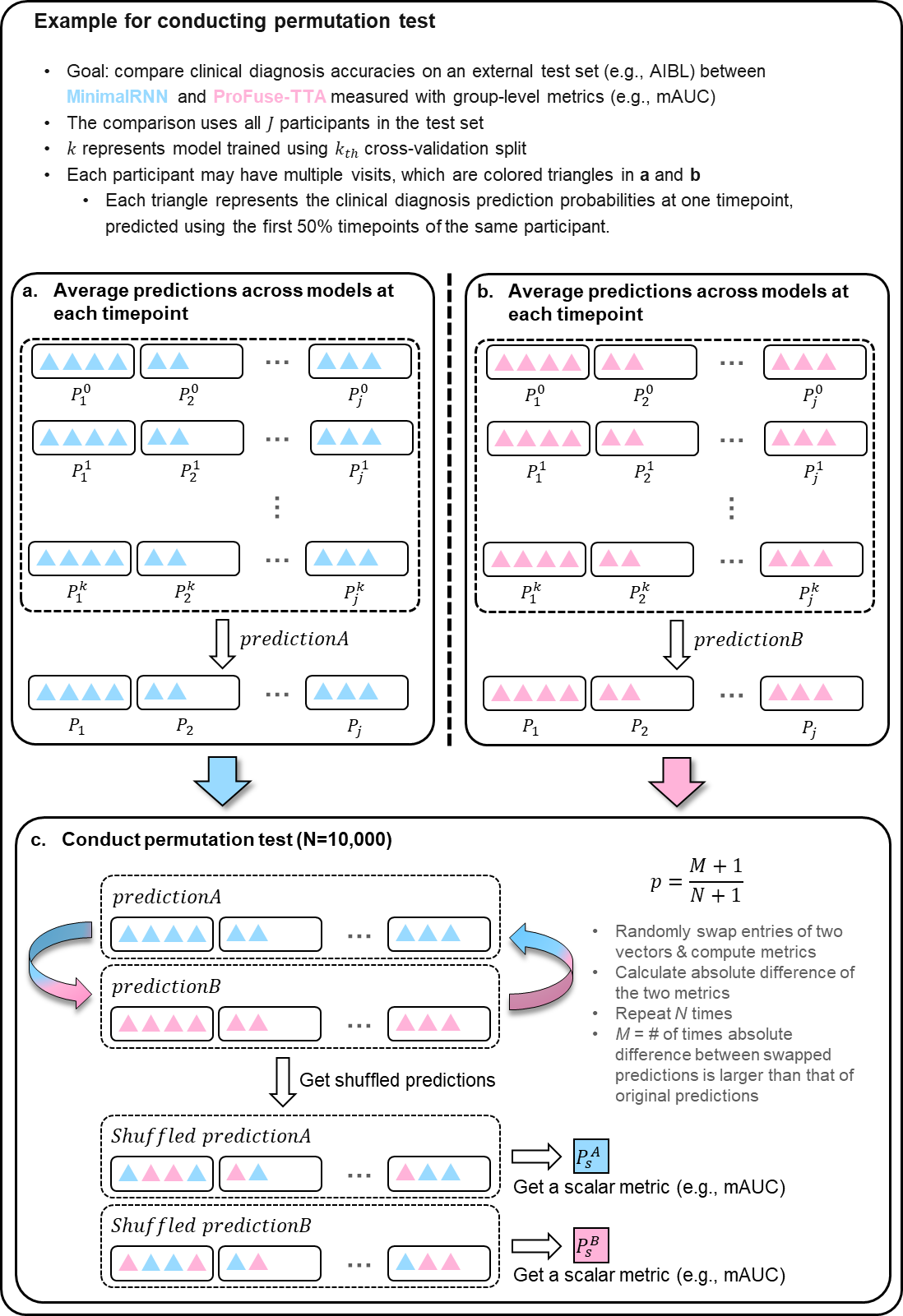

Figure S4. Illustration of permutation test for comparing performance measured with group-level metric

(e.g., diagnostic prediction mAUC) of MinimalRNN and ProFuse-TTA on external test set. (a) For each participant, we averaged the predictions at each timepoint across 20 models for MinimalRNN. (b) Same as A but for ProFuse-TTA. (c) Permute 10,000 times and compute group-level metric (e.g., mAUC) on permuted predictions to obtain p value**.**

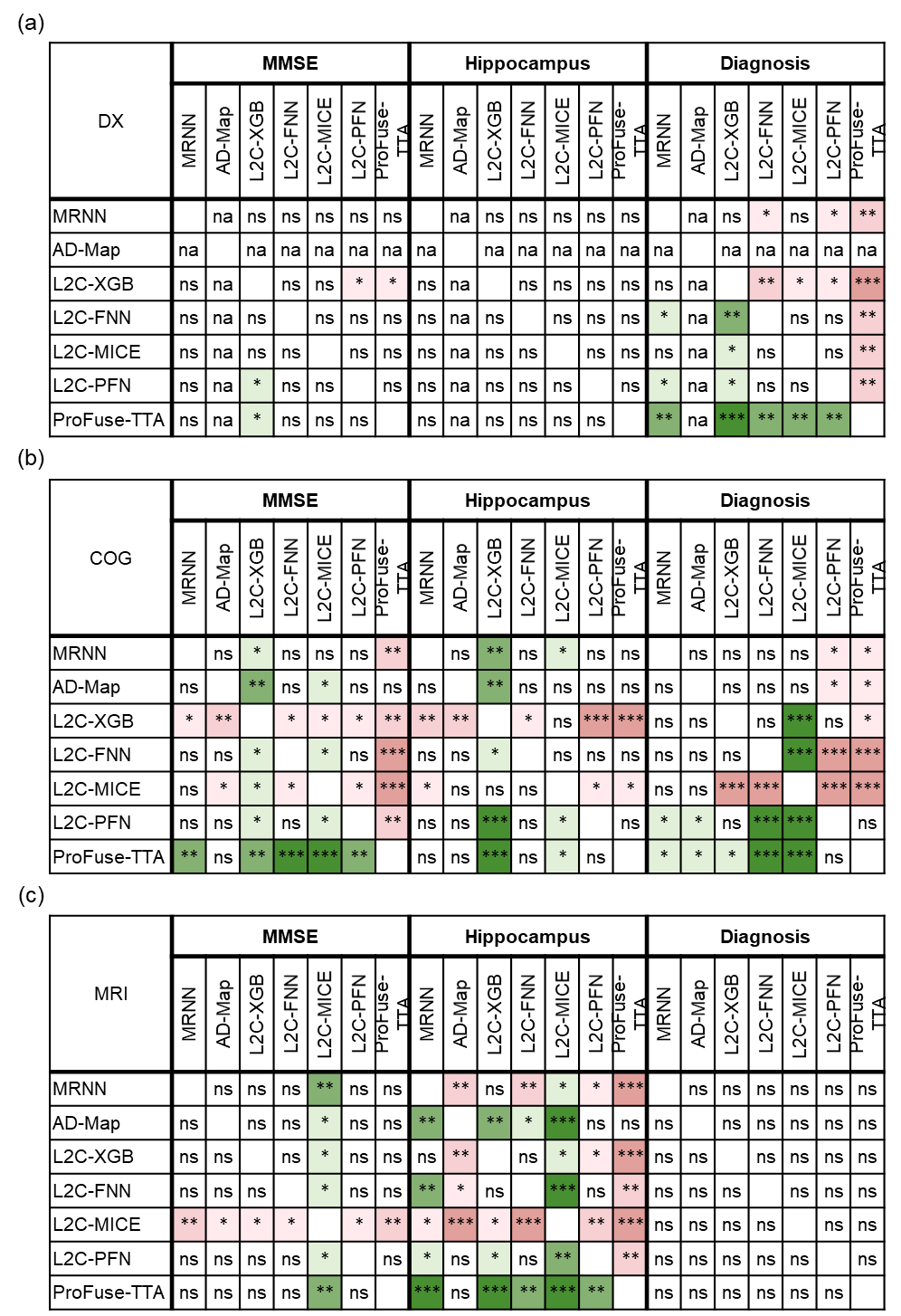

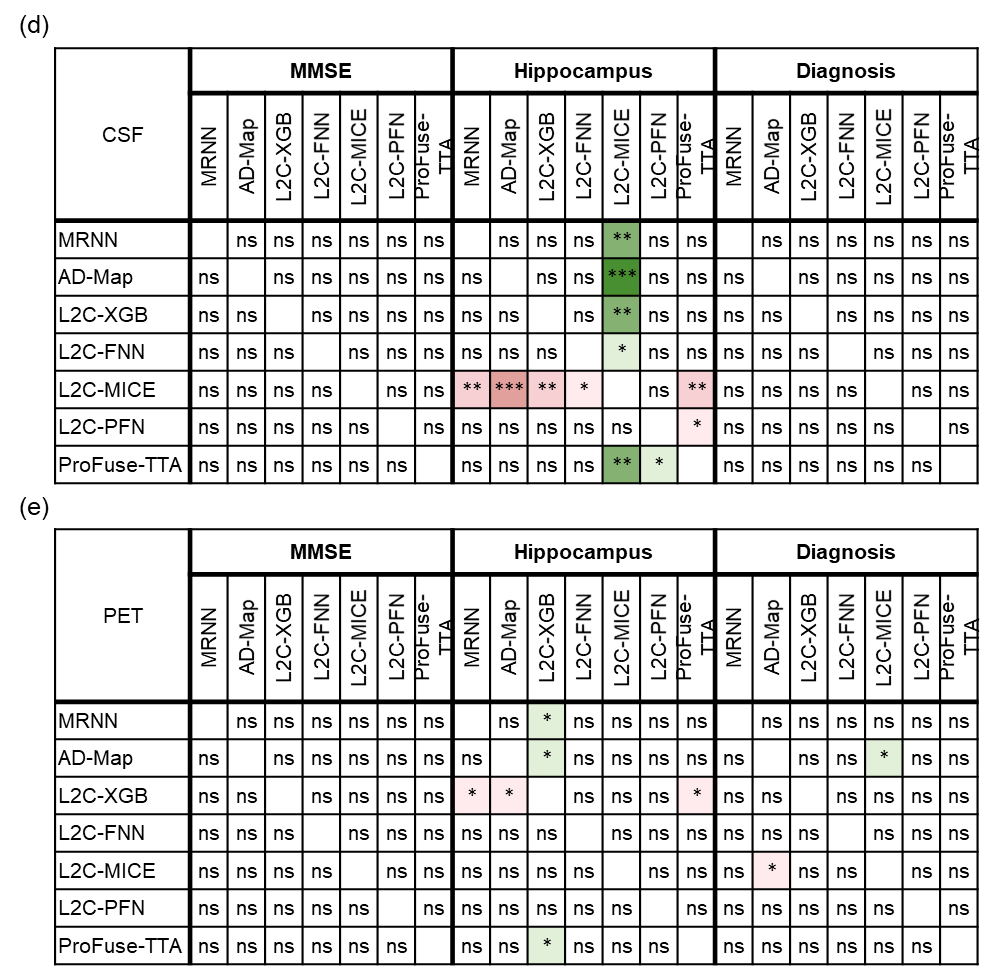

Figure S5. Statistical significance for within-cohort modality ablation.

Statistical significance between all models for within-cohort (ADNI) MMSE, hippocampus volume, and clinical diagnosis prediction performance drops under different modality ablation scenarios (“DX”: ablate diagnosis; “COG”: ablate cognition; “MRI”: ablate MRI, “CSF”: ablate CSF, “PET”: ablate PET). (a) Ablation of “DX”. Columns are grouped into three major columns corresponding to the three prediction tasks. Within each major column, the seven sub-columns represent the seven evaluated models. Each row shows the statistical difference between one model and all other models. For example, the first row corresponds to the statistical difference between MinimalRNN and the other models – green indicates that MinimalRNN performs better, while red indicates that MinimalRNN performs worse. “*****”, “******” and “*******” indicate p < 0.05, p < 0.001, and p < 0.00001 respectively, all surviving multiple comparisons correction (FDR q < 0.05). “ns” indicates no statistical significance (p ≥ 0.05) or did not survive FDR correction. Due to model design, AD-Map did not utilize clinical diagnosis as input features (Section 2.5) and thus did not have results under the “DX” condition. These cases are marked as “N.A.” (not applicable). (b–e) Same as (a), but for ablation of “COG”, “MRI”, “CSF”, and “PET”, respectively.

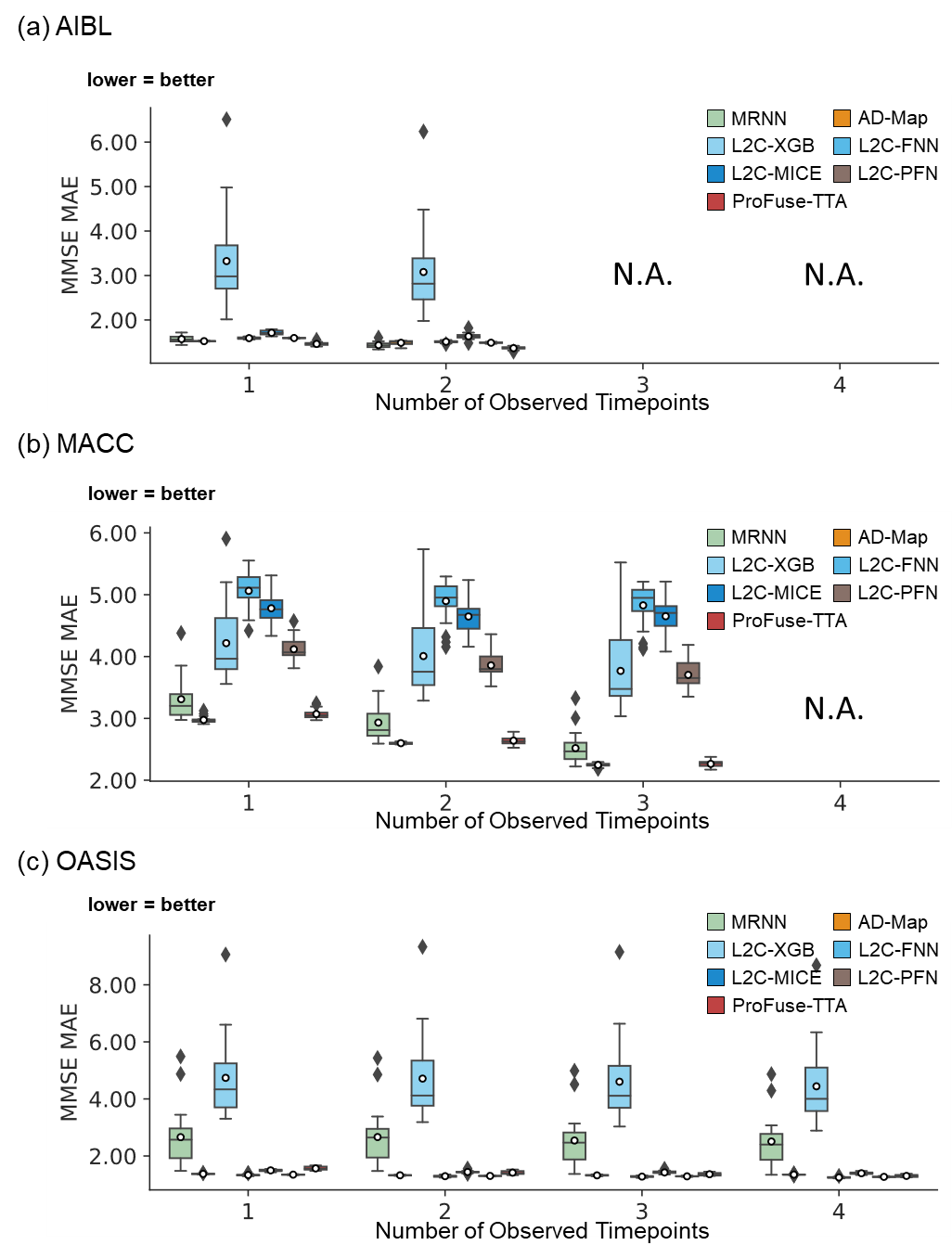

Figure S6. Cross-cohort MMSE prediction performance using different numbers of input timepoints

(after training with all timepoints in ADNI). ProFuse-TTA’s performance was generally competitive, though its relative performance to other models varied depending on the specific dataset and the number of available input timepoints. Results of statistical comparisons between ProFuse-TTA and other approaches are reported in Figure 7.

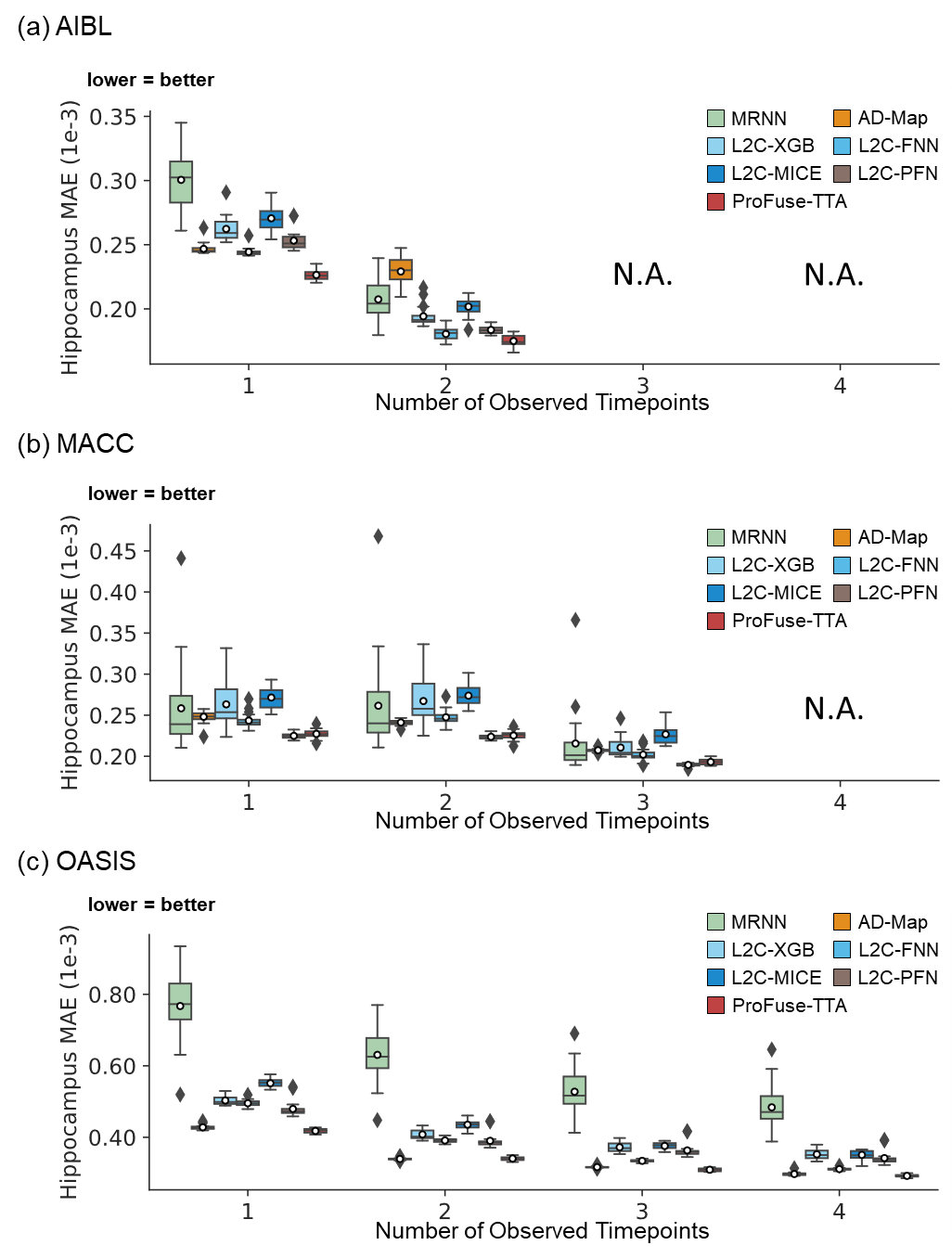

Figure S7. Cross-cohort hippocampus volume prediction performance using different numbers of input timepoints

(after training with all timepoints in ADNI). ProFuse-TTA compared favorably with respect to other approaches across three external test datasets. Results of statistical tests between ProFuse-TTA and other approaches are reported in Figure 7.

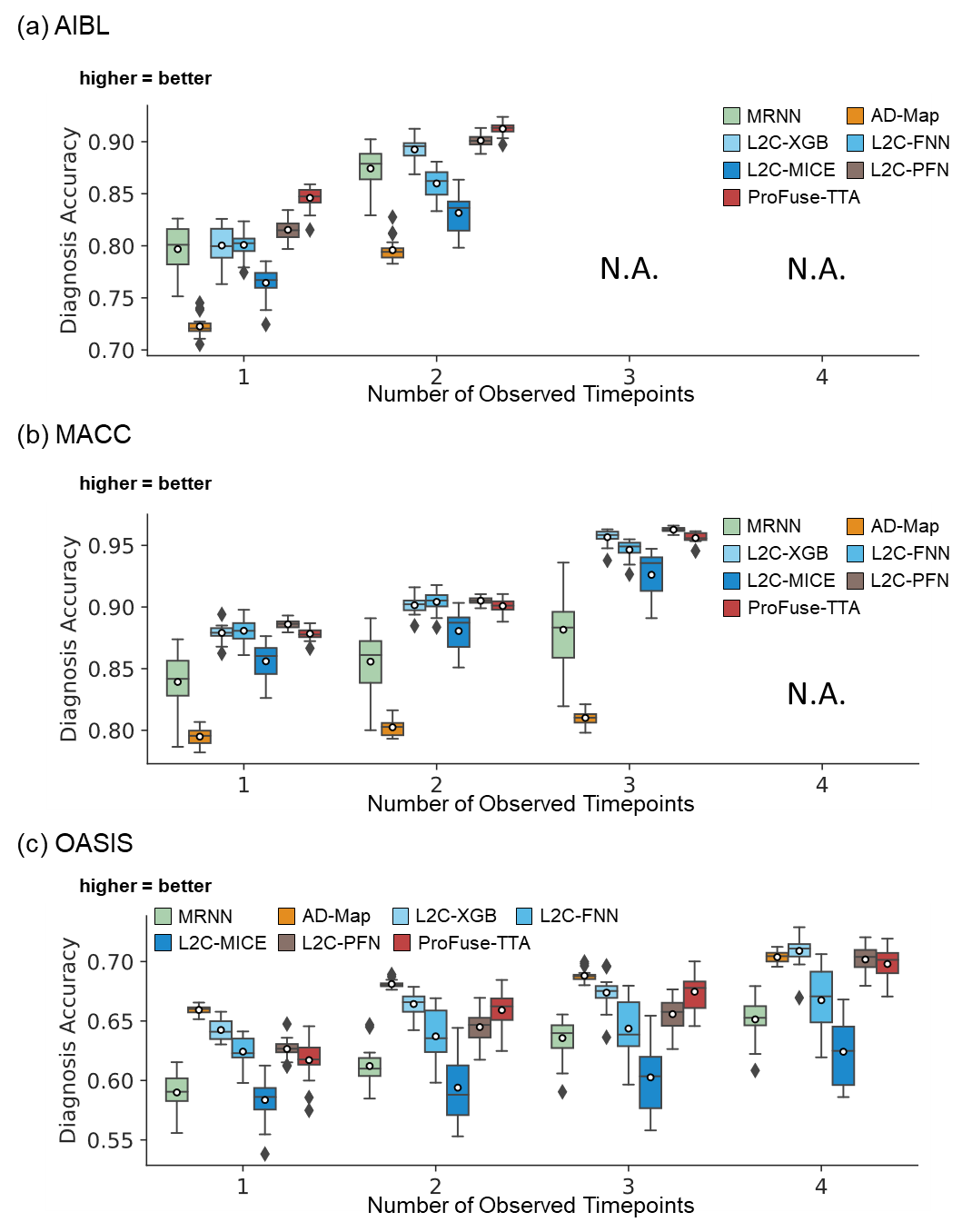

Figure S8. Cross-cohort clinical diagnosis prediction performance using different numbers of input timepoints

(after training with all timepoints in ADNI). ProFuse-TTA compared favorably with respect to other approaches across three external test datasets. Results of statistical tests between ProFuse-TTA and other approaches are reported in Figure 7.

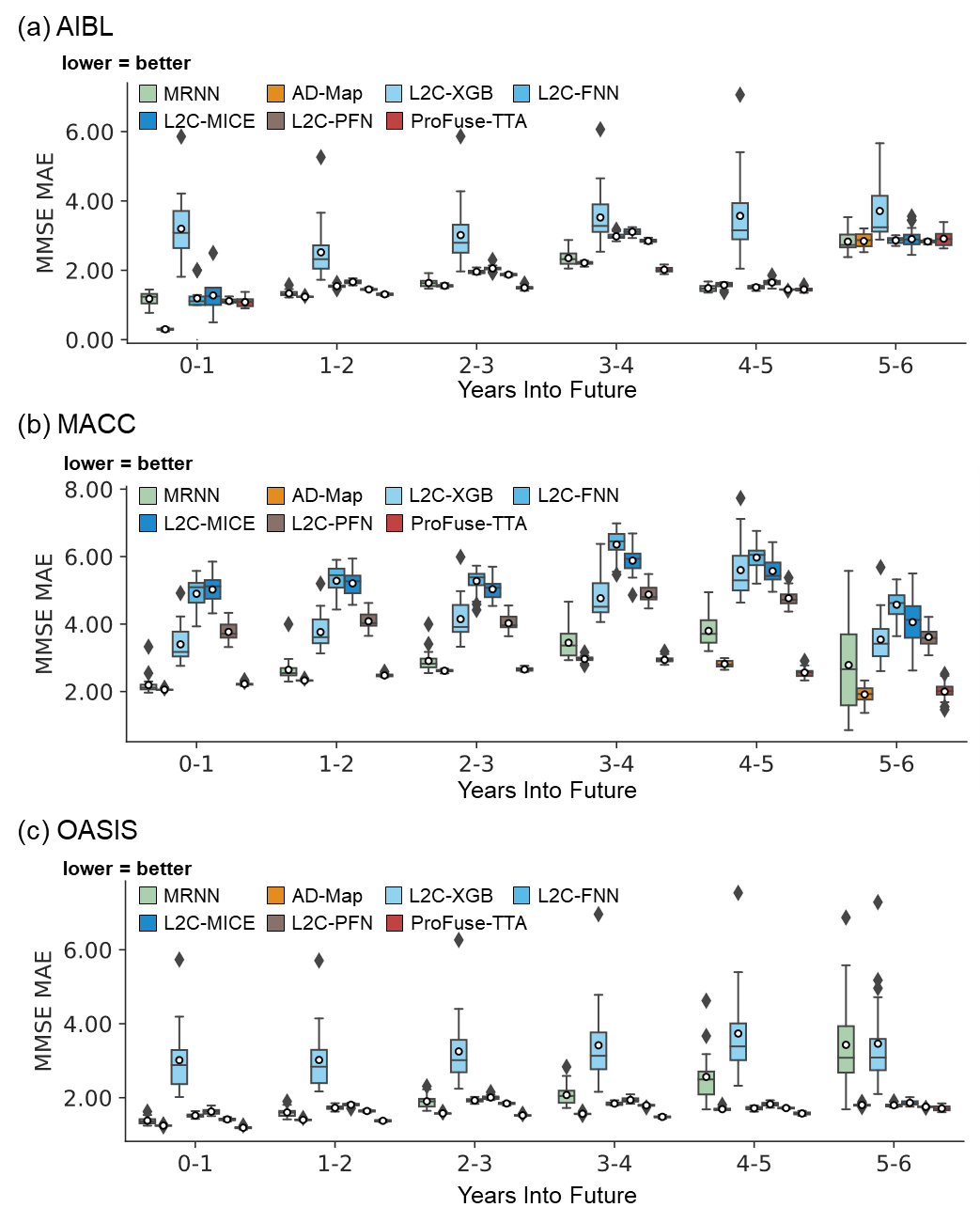

Figure S9. Cross-cohort MMSE prediction performance broken down into yearly intervals up to 6 years into the future.

Note that the last observed time point is at month 0, so year 0-1 means that the prediction was for a future observation at 0 < month ≤ 12, year 1-2 means that the prediction was for a future observation at 12 < month ≤ 24, etc. All algorithms became worse further into the future. ProFuse-TTA was comparable to or better than all models across all years in three external test datasets. Results of statistical tests between ProFuse-TTA and other approaches are reported in Figure 8.

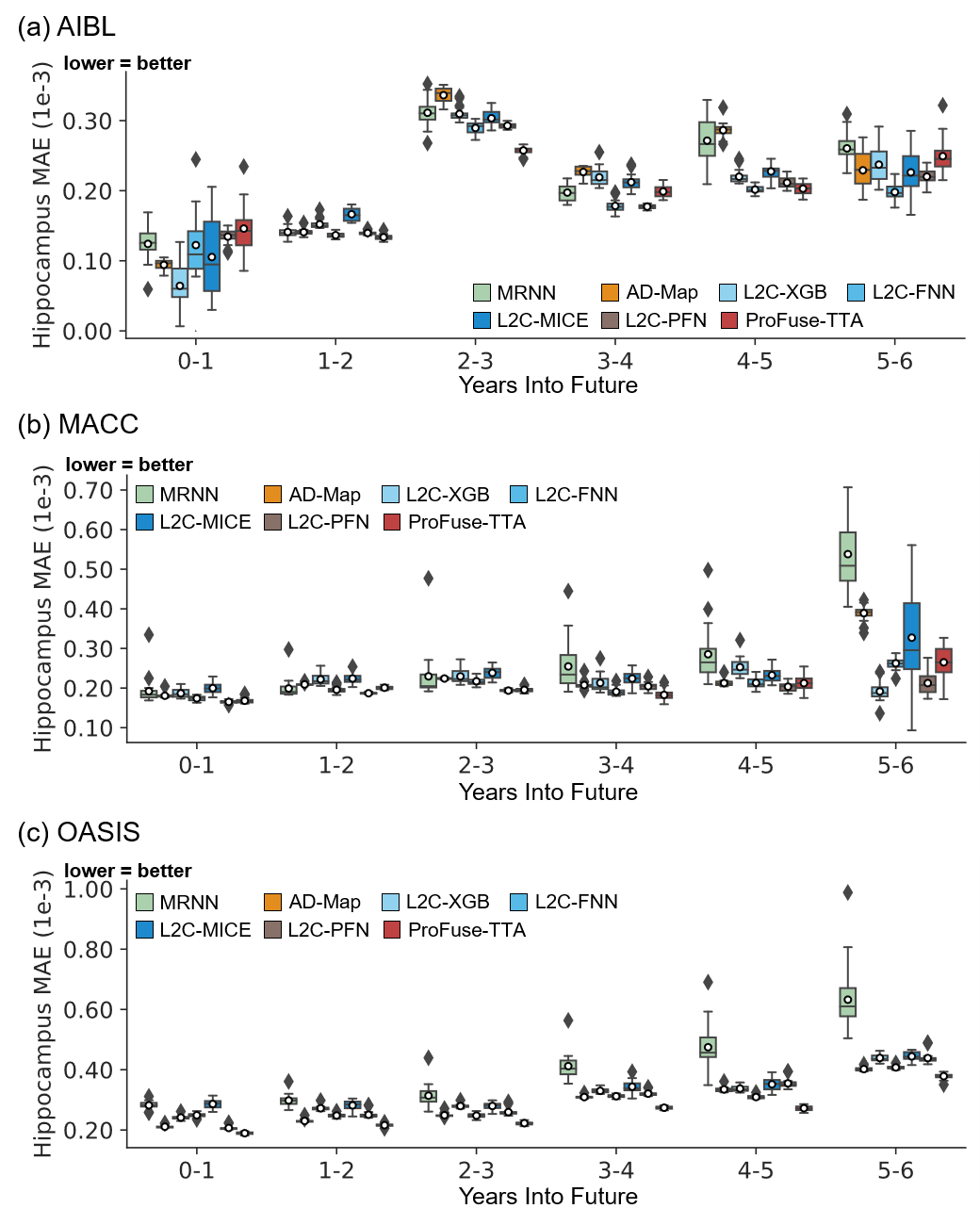

Figure S10. Cross-cohort hippocampus volume prediction performance broken down into yearly intervals up to 6 years into the future.

Note that the last observed time point is at month 0, so year 0-1 means that the prediction was for a future observation at 0 < month ≤ 12, year 1-2 means that the prediction was for a future observation at 12 < month ≤ 24, etc. All algorithms became worse further into the future. ProFuse-TTA was comparable to or better than all models across all years in three external test datasets. Results of statistical tests between ProFuse-TTA and other approaches are reported in Figure 8.

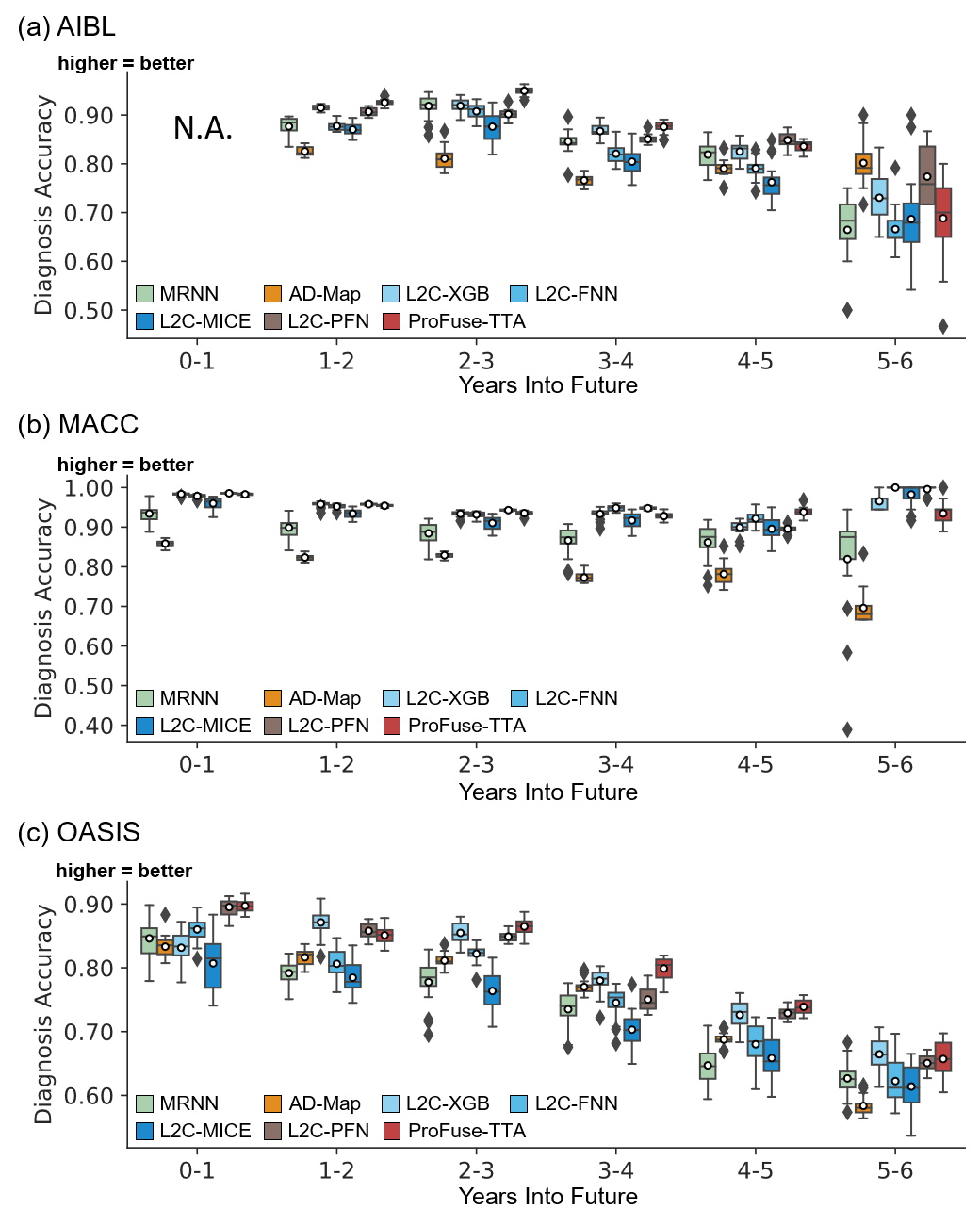

Figure S11. Cross-cohort clinical diagnosis prediction performance broken down into yearly intervals up to 6 years into the future.

Note that the last observed time point is at month 0, so year 0-1 means that the prediction was for a future observation at 0 < month ≤ 12, year 1-2 means that the prediction was for a future observation at 12 < month ≤ 24, etc. All algorithms became worse further into the future. ProFuse-TTA was comparable to or better than all models across all years in three external test datasets. Due to dataset constraints, AIBL includes only one diagnostic class in year 0-1, making mAUC undefined in this case. Therefore, results for AIBL at year 0-1 is marked as “N.A.” Results of statistical tests between ProFuse-TTA and other approaches are reported in Figure 8.

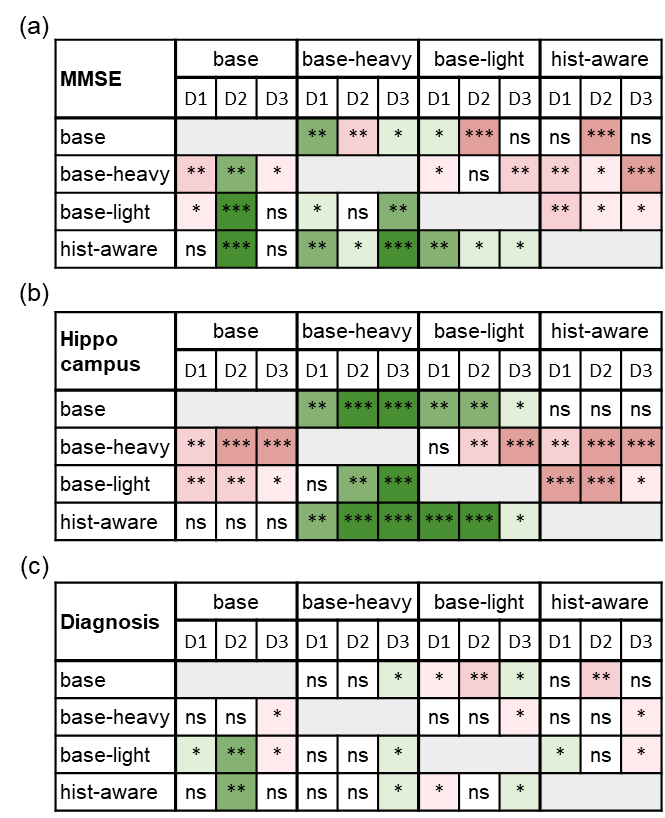

Figure S12. Statistical significance of cross-cohort prediction performance for the ablation study on test-time adaptation strategies,

including heavily tuned full-model adaptation (base-heavy), lightly tuned full-model adaptation (base-light), the proposed history-aware loss-gated strategy (hist-aware), and the non-adapted base model (base), evaluated across three external datasets. (a) Statistical significance for MMSE prediction. The columns are grouped into four major columns corresponding to the four strategies. Within each major column, the three sub-columns represent results on D1: AIBL, D2: MACC, and D3: OASIS datasets, respectively. Each row represents the statistical difference between one strategy and all other strategies. For example, the first row corresponds to the statistical difference between the non-adapted base model with the other strategies – green indicates that base model performs better, while red indicates that base model performs worse. “*”, “**” and “***” indicate p < 0.05, p < 0.001, and p < 0.00001 respectively, all surviving multiple comparisons correction (FDR q < 0.05). “ns” indicates no statistical significance (p ≥ 0.05) or did not survive FDR correction. (b) Same as (a) but for hippocampus volume prediction. (c) Same as (a) but for clinical diagnosis prediction.
